# Evaluating Multi-Ancestry Genome-Wide Association Methods: Statistical Power, Population Structure, and Practical Implications

**DOI:** 10.1101/2025.03.11.25323772

**Authors:** Julie-Alexia Dias, Tony Chen, Hua Xing, Xiaoyu Wang, Alex A. Rodriguez, Ravi K. Madduri, Peter Kraft, Haoyu Zhang

## Abstract

The increasing availability of diverse biobanks has enabled multi-ancestry genome-wide association studies (GWAS), enhancing the discovery of genetic variants across traits and diseases. However, the choice of an optimal method remains debated due to challenges in statistical power differences across ancestral groups and approaches to account for population structure. Two primary strategies exist: (1) Pooled analysis, which combines individuals from all genetic backgrounds into a single dataset while adjusting for population stratification using principal components, increasing the sample size and statistical power but requiring careful control of population stratification. (2) Meta-analysis, which performs ancestry-group-specific GWAS and subsequently combines summary statistics, potentially capturing fine-scale population structure, but facing limitations in handling admixed individuals. Using large-scale simulations with varying sample sizes and ancestry compositions, we compare these methods alongside real data analyses of eight continuous and five binary traits from the UK Biobank (N≈324,000) and All of Us Research Program (N≈207,000). Our results demonstrate that pooled analysis generally exhibits better statistical power while effectively adjusting for population stratification. We further present a theoretical framework linking power differences to allele frequency variations across populations. These findings, validated across both biobanks, highlight pooled analysis as a robust and scalable strategy for multi-ancestry GWAS, improving genetic discovery while maintaining rigorous population structure control.

## Introduction

Genome-wide association studies (GWAS) have played a crucial role in identifying genetic variants associated with various traits and diseases^1,2^. However, these studies have historically been dominated by individuals of European ancestry, who comprised approximately 94.5% of study participants as of 2025^3^, limiting the broader applicability of genetic discoveries. This imbalance poses challenges for the generalizability of genetic findings across populations, since allele frequencies, linkage disequilibrium (LD) patterns, and genetic architectures vary across ancestries^4–6^. To address this gap, researchers have increasingly incorporated participants from diverse genetic backgrounds into multi-ancestry GWAS^7–11^. These studies leverage genetic diversity to identify novel variants and refine polygenic risk scores (PRS), with recent multi-ancestry PRS methods demonstrating improved cross-population risk prediction^12–17^.

Despite these advances, questions remain regarding the optimal methodology for multi-ancestry GWAS. Two primary strategies are commonly used to analyze multi-ancestry GWAS: pooled analysis and meta-analysis and. pooled analysis combines all participants, regardless of ancestry, into a single analysis, adjusting for population stratification using principal components (PCs) in the pooled sample. This method increases the total sample size, enables the inclusion of admixed individuals, and often leads to improved statistical power. However, it raises concerns about inflated false positives if population stratification is not fully accounted for, and may be less effective at capturing subtle local ancestry effects compared to ancestry-group-specific analyses.

Meta-analysis, in contrast, conducts ancestry-group-specific GWAS and then combines the summary statistics^18^. This method better accounts for fine-scale population structure, facilitates data sharing when individual-level data are restricted, and may better account for heterogenous effect sizes across populations. An extension, MR- MEGA^6^, leverages allele frequency differences among contributing studies to boost power and handle admixed individuals. However, this method introduces additional parameters that can reduce power, especially when dealing with complex admixture.

Both strategies can be implemented using fixed-effect or mixed-effect models. Fixed-effect modelling (e.g., PLINK2^19^), assumes genetic effects are constant across individuals, providing computational efficiency, but limited ability to handle cryptic relatedness. In contrast, mixed-effect modelling^20–24^ includes both fixed and random effects to account for population structure and relatedness, enhancing robustness at the cost of increased computational demands. This approach is particularly useful in large biobank studies, where cryptic relatedness is common, and case-control imbalances may introduce biases if not properly accounted for.

In this study, we systematically compare three methods: pooled analysis, fixed-effect meta-analysis, and MR-MEGA^6^, using both fixed-effect and mixed-effect frameworks. Our primary goal is to identify which approach optimally balances statistical power and population stratification control in multi-ancestry contexts. We conduct large-scale simulations with individuals from five ancestry groups, varying sample sizes, ancestry-group proportions, and outcomes (continuous and binary). To further assess the impact of varying levels of admixture, we simulate admixed individuals using the Admix-kit pipeline^25^. Finally, we validate our findings in real-world data by analyzing eight continuous and five binary traits from two large biobanks, the All of Us Research Program^26^ (AoU, N≈207,000) and the UK Biobank^27^ (UKB, N≈324,000).

Our results demonstrate that pooled analysis achieves higher statistical power than meta-analysis and MR-MEGA across a range of study designs while maintaining well-controlled type I error in realistic scenarios. We further propose a theoretical framework linking these power gains to allele frequency differences across ancestry groups. These findings support pooled analysis as a robust and scalable approach for multi-ancestry GWAS, improving genetic discovery and enhancing the generalizability of GWAS findings across populations.

## Material and Methods

### Theoretical Motivation for Method Discrepancies

Consider a multi-ancestry cohort comprising 𝐽 distinct subcohorts (ancestry groups). Let 𝑛_j_ denote the number of subjects in subcohort 𝑗, and let 𝑓_j_ be the allele frequency of a causal variant in subcohort 𝑗. We assume that the allelic effect of this variant (𝛽) is constant across ancestry groups. For individual 𝑖 in population 𝑗, the genotype 𝑔_𝑖j_ follows a binomial distribution 𝑔_𝑖j_ ∼ 𝐵𝑖𝑛(2, 𝑓_j_). The phenotype is defined as

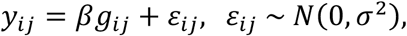

where 𝑖 = 1, … , 𝑛_j_ and 𝑗 = 1, … , 𝐽, and 𝜎^2^ represents the residual variance.

### Within-Population GWAS

If we conduct a separate linear regression in each subcohort (with intercept), we obtain

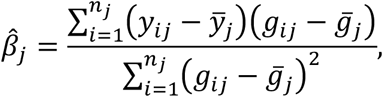

and

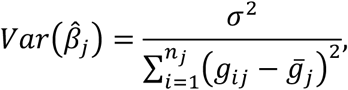

where 𝑦̅_j_ and 𝑔̅_j_ are the phenotypic and genotype means, respectively, in subcohort 𝑗.

### Pooled Analysis GWAS

When pooling all individuals across the 𝐽 subcohorts into a single GWAS, we have:

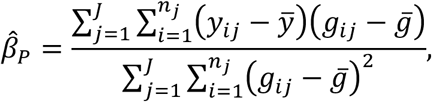

and

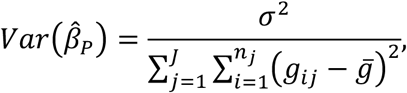

where 𝑦̅ and 𝑔̅ are the global phenotype and genotype means, respectively.

### Fixed-effect Meta-analysis

Using inverse-variance weighting, the fixed-effect meta-analysis estimate is

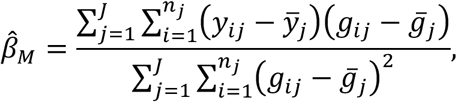

and

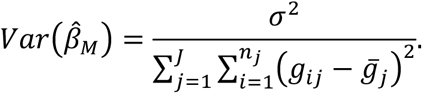

As 𝑛_j_ → ∞, for all 𝑗, both *β̂_M_* and *β̂_P_* converge to 𝛽.

### Asymptotic Relative Efficiency

We define the asymptomatic relative efficiency (ARE) of meta-analysis vs. pooled-analysis as:

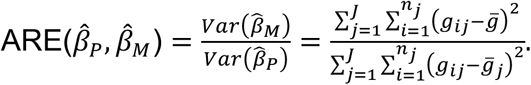

It follows that

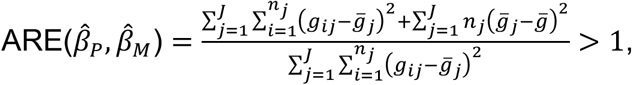

because of the standard ANOVA decomposition:

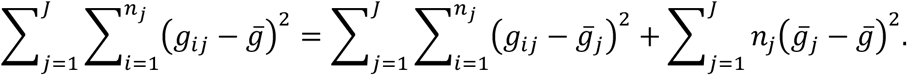

The term 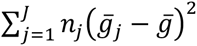 represents the sample-size-weighted variance in allele frequencies across populations. When allele frequencies differ substantially across subcohorts, the pooled analysis benefits from leveraging higher variance in carrier counts across populations, improving power. If one population is disproportionately large (e.g. 𝑛_2_ ≪ 𝑛_1_ in a two-population settings), the advantage of pooled analysis diminishes because the dominant population drives the association signal, and the sample-size-weighted variance in allele frequencies will be small.

## Implementation of Methods

We applied the following approaches in both simulations and real data analyses using AoU and UKB datasets.

### Pooled and Meta-analysis

For pooled analysis, GWAS was performed on the entire dataset by combining all ancestry groups into a single analysis. Population structure was adjusted using top ten cross-ancestry-group PCs derived from the full pooled dataset, along with covariate adjustments for age and sex. Variants were included in the pooled analysis if they had MAF >1% in at least one ancestry group. For meta-analysis, GWAS was conducted separately within each ancestry group. Top ten ancestry-group-specific PCs were used for population structure adjustment, along with age and sex as covariates. Each ancestry-specific GWAS included only variants with MAF >1% within that ancestry group. Summary statistics from individual ancestry analyses were then combined using a fixed-effect meta-analysis approach with inverse-variance weighting. As a result, variants were included in the final meta-analysis if they met the MAF >1% threshold in at least one ancestry group, ensuring comparability with pooled analysis.

Both approaches used REGENIE^20^ (--qt for continuous traits and --bt for binary traits) for the mixed-effect modelling and PLINK2^19^ (--linear for continuous traits) for the fixed-effect modelling. REGENIE was selected as a scalable approach for mixed-model association testing, as it uses a local ridge regression framework to adjust for population structure and relatedness. While REGENIE does not explicitly introduce a random effect term, its regularization-based approach functionally approximates mixed-effect modelling by accounting for cryptic relatedness and polygenic effects, distinguishing it from standard fixed-effect models implemented in PLINK2.

### MR-MEGA

MR-MEGA extends meta-analysis by incorporating ancestry-group-specific summary statistics while explicitly accounting for allele frequency differences across ancestry groups. MR-MEGA models ancestry-group-specific genetic effects as a function of population structure, using PCs derived from allele frequency variation across groups. The input for MR-MEGA is similar to standard meta-analysis, using summary statistics from either mixed-effect or fixed-effect GWAS results. The number of axes of genetic variation used in MR-MEGA (--pc) was set to the maximum of the number of distinct genetic ancestry groups minus three. Specifically, for UKB samples, --pc was set to 2, while for AoU samples, --pc was set to 3. MR-MEGA was applied only when combining data from at least four ancestry groups. For example, MR-MEGA was not used for prostate cancer in UKB, as only three ancestry groups had a sufficient number of cases. It is important to note that: MR-MEGA restricts its output to variants with MAF > 1% in all genetic ancestry groups, while meta-analyses include variants with MAF > 1% in at least one ancestry group.

### Type I Error Simulations

To evaluate Type I error, we used real genomic data from six genetic ancestry groups as defined in AoU^26^: African (AFR), Admixed American (AMR), East Asian (EAS), European (EUR), Middle Eastern (MID) and South Asian (SAS), to simulate null phenotypes under varying degrees of population stratification. Employing actual genotype data allows us to capture realistic LD structures within each ancestry. For both ancestry-group-specific and cross-ancestry-group scales, stratification was introduced by varying the percentage of the null phenotype’s variance explained by the first ten PCs, ranging from 0% to 5%.

### Percentage of Variance Explained by PCs

Before simulating phenotypes, we estimated the variance explained by the first ten PCs in real polygenic traits. Five representative traits were selected: height, high-density lipoprotein cholesterol (HDL), low-density lipoprotein cholesterol (LDL), total cholesterol (TC), and waist circumference. To quantify the variance explained by ancestry-group-specific PCs and cross-ancestry-group PCs, we applied a two-step regression approach. First, within each ancestry group, we regressed out age and sex from the trait values and obtained the residuals. Next, we estimated the proportion of variance explained (𝑅^2^) for each PC set by comparing models with different covariate adjustments. The variance explained by ancestry-group-specific PCs was computed as the difference in 𝑅^2^ between the full model, which included both cross-ancestry-group PCs and ancestry-group-specific PCs, and the reduced model, which included only cross-ancestry-group PCs. Similarly, the variance explained by cross-ancestry-group PCs was measured as the difference in 𝑅^2^ between the full model, which included both PC sets, and the reduced model, which included only ancestry-group-specific PCs. Confidence intervals are obtained via bootstrapping using 10,000 replicates (**Supplementary Table 1**). These empirical estimates guided our selection of 𝛼 values (0,0.2%, 0.5%, 1%, 2%, 5%) in the subsequent simulations.

### Null Phenotype Generation

We simulated population stratification separately on ancestry-group-specific and cross-ancestry-group scales. Empirical results (**Supplementary Table 1**) indicated that after accounting for ancestry-group-specific PCs, cross-ancestry PCs explained 0% variance, justifying this independent modelling. To simulate ancestry-group-specific population stratification , we set

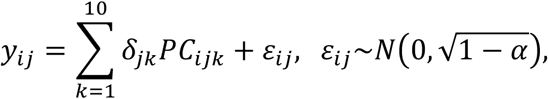

where 𝑃𝐶_𝑖j𝑘_ is the 𝑘^𝑡ℎ^ PC for individual 𝑖 in ancestry 𝑗. 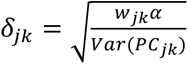, is the effect for 𝑘th ancestry-group-specific PC in ancestry 𝑗, 𝛼 is the total percentage of variance explained by the first ten ancestry-group-specific PCs, and 𝑤_j𝑘_ = 𝜆_j𝑘_⁄𝜆_j_, with 𝜆_j𝑘_ being the eigenvalue of the 𝑘^𝑡ℎ^ PC in ancestry 𝑗, and 𝜆_j_ the sum of the ten eigenvalues for ancestry 𝑗. This formulation of 𝛿_j𝑘_ ensures that population stratification effects are proportionally assigned across PCs while preserving phenotype variance. The resulting null phenotype has mean 0 and variance 1 within each ancestry group.

Similarly, to simulate cross-ancestry-group population stratification, we used

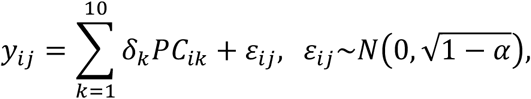

where 𝑃𝐶 is the 𝑘^𝑡ℎ^ cross-ancestry-group PC for individual 𝑖, 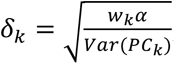, is the effect for 𝑘th cross-ancestry-group PC, 𝛼 is the total percentage of variance captured by the top ten cross-ancestry-group PCs, and 𝑤_𝑘_ = 𝜆_𝑘_⁄𝜆, with 𝜆_𝑘_ as the eigenvalue of the 𝑘^𝑡ℎ^ cross-ancestry-group PC, and 𝜆 as the sum of the eigenvalues of top ten cross-ancestry-group PCs. As in the ancestry-group-specific case, this design ensures that the null phenotype remains standardized with mean 0 and variance 1, while incorporating realistic population structure.

We simulated 𝛼 ∈ (0,0.2%, 0.5%, 1%, 2%, 5%) to assess different degrees of population stratification. These scenarios reflect both observed values (e.g., 0% − 0.5%), and hypothetical extremes (e.g. 1% − 5%). This setup allowed us to assess whether each GWAS method (pooled, meta-analysis, MR-MEGA) controlled type I error appropriately in the presence of population structure on different scales.

### Statistical Power Simulations

#### Continuous Phenotypes

We used a previously published simulated dataset of 600,000 independent subjects^13^ (120,000 from each of five ancestry groups: AFR, AMR, EAS, EUR, SAS), generated by HAPGEN2^28^ and the 1000 Genomes Project (1000G)^29^ as reference for LD and allele frequency structures (**Supplementary Figure 1**). To generate continuous phenotypes, we randomly selected 5% of variants from the HapMap3 variant list^30^ to be causal. Phenotypes for individual 𝑖 in population 𝑗 were simulated as:

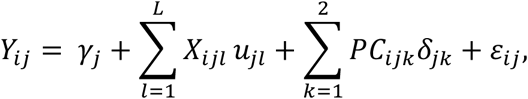

where 𝛾_j_ ∼ 𝑁(0,1) is an ancestry-group-specific intercept term. 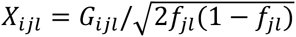 is the standardized genetic value for variant 𝑙 with mean 0 and variance 1. Under the assumption that variant effects on the standardized scale are constant across ancestry groups, we sampled causal effect sizes 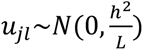, with 𝐿 being the number of causal variants, and the heritability ℎ^2^ being set to 0.4. This implies that the per-allele effect size varies with respect to the MAF in each group. Two ancestry-group-specific PCs were included, with 𝛿_j𝑘_ assigned based on the empirical estimates from UKB. The residual error variance was modeled as,

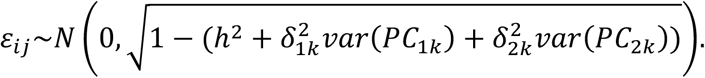

Ten replicates of continuous phenotypes were generated.

#### Binary Phenotypes

To generate binary traits, we simulated a larger dataset of five million individuals (one million per ancestry group) using HAPGEN2 with 1000G as reference, to ensure adequate sample sizes for case-control sampling and accurate representation of allele frequency and LD structures. We randomly selected 1% causal variants across the genome. The binary outcomes were generated using a logistic model:

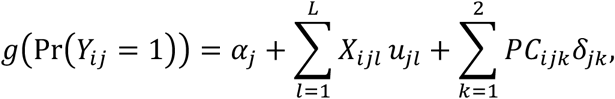

where 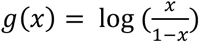, is the logit function. Similarly as continuous outcomes, we assume 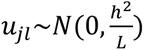, where logit-scale genetic variance ℎ^2^ was set to 0.57 to mimic breast cancer ^31^. The intercept term 𝛼_j_ was used to control disease prevalence. We assigned different 𝛼_j_ to mimic the estimated prevalence of breast cancer from [SEER*Stat](https://seer.cancer.gov/seerstat/) based on the past 28 years (**Supplementary Table 2**). After generating case-control status for all individuals, we randomly sampled cases and controls in a one-to-one ratio to create datasets with an average total sample size of 237,241 cases and 237,241 controls (detailed sample sizes by ancestry group described in **Supplementary Table 3**).

#### Admixed Individuals

To evaluate admixture effects, we simulated an additional dataset of 240,000 individuals using Admix-kit^25^ and 1000G as the reference. The simulated dataset included four groups: 1. 60,000 individuals with 100% EUR ancestry 2. 60,000 individuals with 100% EUR ancestry 3. 60,000 subjects with 50% EUR and 50% AFR admixture 4. 60,000 subjects with 20% EUR and 80% AFR admixture (Supplementary Figure 2).

Admix-kit is a simulation tool that generates realistic admixed genomes by combining haplotypes from multiple ancestral populations. It models admixture as a generational process, where individuals inherit ancestry segments based on recombination patterns over multiple generations. The simulation was conducted in two steps. First, we expanded the reference haplotype data using HAPGEN2, generating a larger pool of phased haplotypes for EUR and AFR populations. To improve computational efficiency, we restricted the variants to those included in the HapMap3 variant list^30^. In the second step, we simulated admixture by drawing haplotypes from the expanded EUR and AFR datasets using the admix-simu function in Admix-kit. We modelled ten generations of admixture (--n-gen 10), allowing for recombination to shape local ancestry patterns.

Ancestry proportions were assigned using the --admix-prop flag to reflect the desired admixture levels in each group. Continuous phenotypes for admixed individuals were simulated following the same methodology described in the previous section.

#### Statistical Power Evaluation

We used two metrics to evaluate the performance of GWAS methods: 1. Exact recovery of causal variants, which measures the percentage of causal variants that are directly identified at genome-wide significance with 𝑃 < 5 × 10^−8^. It is defined as the number of causal variants reaching genome-wide significance. 2. LD-based recovery of causal variants, which assesses the percentage of causal variants located within LD regions of genome-wide significant variants.

Specifically, it is calculated as the number of causal variants with at least one genome-wide significant variant within a 500 kb region, divided by the total number of causal variants.

### Real Data Analysis in the All of Us Research Program and UK Biobank

For both AoU and UKB datasets, we analyzed eight continuous traits and five binary traits. The continuous traits included height, waist circumference, LDL, TC, HDL, calcium, creatinine, and estimated glomerular filtration rate (eGFR). The binary traits examined were asthma, coronary artery disease (CAD), type II diabetes (T2D), breast cancer (BC), and prostate cancer (PC). The corresponding concept IDs for AoU and Data Field IDs for UKB are provided in **Supplementary Table 4**. Subjects with measurements outside predefined ranges for each phenotype were excluded (**Supplementary Table 4**).

### Data Processing and Sample Selection in AoU

For AoU analyses, we utilized phenotype and genotype array data from AoU version 7.1. Participant information was collected in accordance with the <u>AoU Research Program Operation Protocol</u>. Detailed procedures related to genotyping, ancestry classification, quality control measures, and the exclusion of related participants are comprehensively documented in the [AoU Research Genomic Research Data Quality Report](https://support.researchallofus.org/hc/en-us/articles/4617899955092-All-of-Us-Genomic-Quality-Report-ARCHIVED-C2022Q4R9-CDR-v7). The AoU data includes both genotype array data and short-read whole genome sequencing (srWGS) data. Although genotype array data encompasses a larger sample size, ancestry and family relatedness information were only available for individuals with srWGS data. To ensure consistent ancestry classification, we restricted analyses to individuals with srWGS data, but we used genotype array variants for GWAS. Variants were filtered based on a MAF threshold of > 0.01 within any ancestry group, resulting in approximately 1.2 million variants.

We excluded flagged related subjects, subjects with ‘sex_at_birth’ missing (e.g., “Skip”, “I prefer not to answer”, “No matching concept”, “None”), and individuals with a predicted ancestry probability was ≤ 0.75. This threshold was applied to ensure a consistent comparison between pooled and meta-analysis approaches, as meta-analysis requires pre-defined ancestry groups. Including these individuals, who are likely admixed, would have been inconsistent with the ancestry-group-based framework used in meta-analysis. After these exclusions, the final dataset included 207,305 subjects across six genetic ancestry groups: 47,207 AFR, 36,500 AMR, 5,153 EAS, 115,701 EUR, 497 MID and 2,247 SAS (**Supplementary Figure 3**). Phenotype values were taken from the most recent available measurement, and age was calculated as the difference between the date of birth and the date of the latest phenotype measurement.

#### Data Processing and Sample Selection in UKB

For UKB, we classified individuals into five major continental ancestry groups (AFR, AMR, EAS, EUR, SAS) using the 1000G^29^ as a reference. A random forest classification model was trained on the 1000G dataset, using the top 20 PCs as predictors and known ancestry group labels as outcomes. This trained model was then applied to the top 20 PCs within UKB to infer genetic ancestry. To minimize population structure confounding, we restricted analyses to unrelated individuals, resulting in a final dataset of 323,908 subjects across five genetic ancestry groups (6,864 AFR, 590 AMR, 586 EAS, 311,053 EUR and 5,734 SAS) (**Supplementary Figure 4**). Age in UKB was based on ‘age at visit’ (Data-Field 21003). Variants for UKB analyses were restricted to HapMap3^30^ + Multi-Ethnic Genotyping Arrays^32^ (MEGA) chip array, and further filtered using a MAF threshold of > 0.01 within at least one ancestry group, resulting in approximately 1.53 million variants.

#### PCA Computation

To compute PCs, we pruned autosomal variants with MAF >1% across the pooled dataset, using an 𝑟^2^ threshold of 0.1, a window size of 500kb with PLINK2^19^ (--indep-pairwise). This resulted in a set of 125,692 variants in AoU and 51,922 variants in UKB. The first 10 PCs were then computed separately within each genetic ancestry group and within the pooled dataset using PLINK2^19^ (--pca-approx).

## Results

We evaluated three multi-ancestry GWAS strategies: meta-analysis, MR-MEGA^6^ and pooled analysis (**Figure 1**). Meta-analysis was implemented with standard fixed-effect meta-analysis using inverse variance weighting (**Materials and Methods**). All primary analyses employed mixed-effect modelling with REGENIE^20^ in the main text, while analyses using fixed-effect modelling was performed with PLINK2^19^ in the Supplementary Material. In pooled analysis, individuals from all ancestry groups were combined into a single dataset, with cross-ancestry-group PCs controlling for population stratification. In contrast, meta-analysis stratified individuals into genetically homogeneous ancestry groups before combining their summary statistics. MR-MEGA refined meta-analysis by explicitly modelling allele frequency differences among ancestry groups.

**Figure 1:**
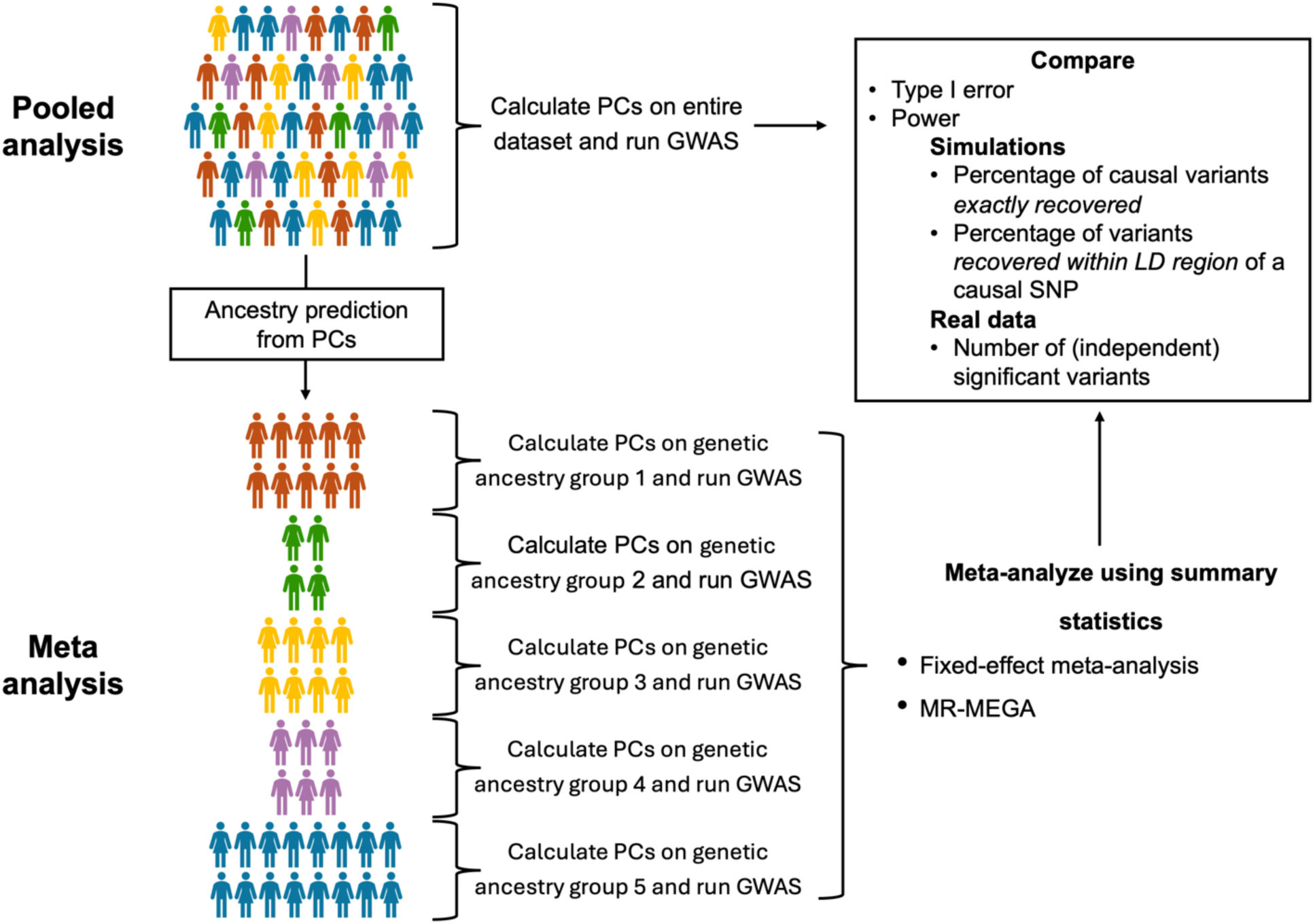
Overview of analysis. In the pooled analysis (top), principal components (PCs) are computed for the entire dataset, followed by a single GWAS. In the meta-analysis (bottom), PCs are calculated separately for each genetic ancestry group, and independent GWAS are conducted for each. Summary statistics are then combined through meta-analysis using inverse-variance weighting or MR-MEGA. The two approaches are compared based on type I error and statistical power. In simulations with causal variants statistical power is assessed by the percentage of causal variants exactly recovered and those within the 500kb linkage disequilibrium (LD) region of a causal variants. In real data analyses, statistical power is measured by the number of significant variants (𝑃 ≤ 5 × 10^−8^) and the number of independent signals (linkage disequilibrium 𝑟^2^ < 0.1).

Our comparisons of multi-ancestry GWAS methodologies focused on two core metrics: 1. Type I error, evaluating whether each method properly controls false positives in the presence of population stratification 2. Statistical power, measured by (a) exact causal variant detection, which measures the proportion of causal variants reaching genome-wide significance, and (b) LD-based detection, which measures proportion of significant variants within 500 kb of a causal variant.

### Statistical Power Discrepancies Between Pooled and Meta-analysis

Under the assumption of homogeneous allelic effects across ancestry groups, differences in statistical power between pooled and meta-analysis depend on allele frequency variance across populations (**Figure 2**). When MAFs are similar across populations, both methods yield comparable power. However, as MAF divergence increases, the relative advantage of pooled analysis becomes more pronounced (**Materials and Methods**). This advantage arises because pooled analysis integrates information across populations, allowing individuals from groups with high MAF to contribute more effectively to association detection. In contrast, meta-analysis treats each population separately before combining results, limiting the ability of high-MAF groups to enhance power in low-MAF populations. The power gain of pooled analysis is maximized when allele frequencies vary across ancestries and sample sizes are balanced. However, if one population is disproportionately large, the benefits of pooling diminish, as the dominant population drives association signals, reducing the influence of smaller ancestry groups. Overall, pooled analysis achieves better statistical power at a particular locus when allele frequencies differ across populations, with its advantage further amplified when highly divergent alleles are well-represented in larger sample sizes.

**Figure 2:**
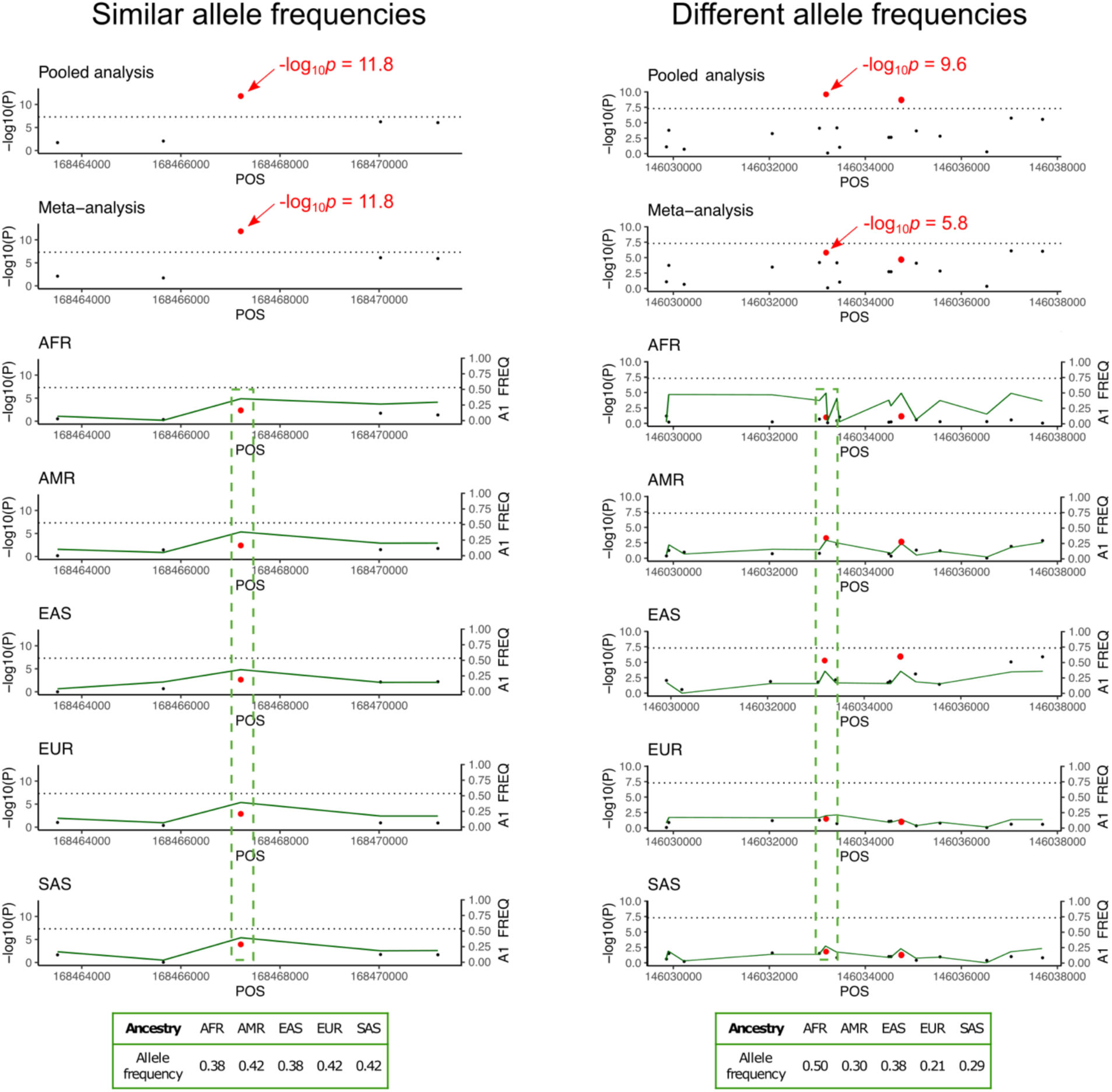
Illustration of pooled vs. meta-analysis approaches in multi-ancestry GWAS based on allele frequency differences. This figure demonstrates the theoretical differences in power between pooled and meta-analysis approaches in multi-ancestry GWAS are mainly driven by allele frequency differences. Manhattan plots show simulated results for two known causal variants under two scenarios: 1. Similar allele frequencies across ancestry groups (chromosome 4, rs11936438) result in equal statistical power for both pooled and meta-analysis (− log_10_ 𝑝 = 11.8, shown in left panel), 2. Divergent allele frequencies (chromosome 5, rs404167) show higher power in pooled analysis (−log_10_ 𝑝 = 9.6) compared to meta-analyses (−log_10_ 𝑝 = 5.8, shown in the right panel). The lower panels display ancestry-group-specific results for simulated ancestry groups: African (AFR), Admixed American (AMR), East Asian (EAS), European (EUR), and South Asian (SAS). The green line represents the minor allele frequency. The tables at the bottom summarize allele frequencies of the causal SNPs across ancestry groups, highlighting how different allele frequencies impact the statistical power of each approach.

### Type I Error Simulations

We evaluated type I error by simulating null phenotypes across six populations using AoU data, ensuring that LD patterns and population stratification reflected real genomic architectures. We investigated both local population stratification, where the phenotype was associated with ancestry-group-specific PCs, and global population stratification, where the phenotype was associated with cross-ancestry-group PCs. To model varying degrees of stratification, we varied the proportion of the null phenotype’s variance explained by the first ten ancestry-group-specific and cross-ancestry-group PCs (**Material and Methods**).

Under both fixed- and mixed-effect modelling approaches, type I error was well controlled with meta-analysis, regardless of the magnitude of ancestry-group-specific or cross-ancestry-group PC effects. Pooled analysis also controlled type I error when cross-ancestry-group PCs were associated with the outcome but exhibited inflation when more than 1% of the null phenotype’s variance was explained by ancestry-group-specific PCs. (**Figure 3** for mixed-effect modelling; and **Supplementary Figure 5** for fixed-effect modelling). However, real-world traits typically exhibit much lower ancestry-specific PC variance. In AoU, the median variance explained by the first ten ancestry-specific PCs is only 0.26% across the three largest ancestry groups (**Supplementary Table 1**). Higher ancestry-group-specific PC variance (>1%) was primarily observed in smaller ancestry groups such as EAS, SAS, and MID, where estimates may be less stable due to limited sample sizes. While large multi-ancestry GWAS may include some populations with small samples sizes, it is unlikely that most participating ancestry groups exhibit extreme PC variance exceeding 1%. Notably, our simulations only showed inflated p-values when multiple ancestries had PC variance above this threshold. This suggests that the level of ancestry-group-specific population structure required to induce meaningful bias in pooled analysis is substantially greater than what is typically observed in well-powered studies.

**Figure 3:**
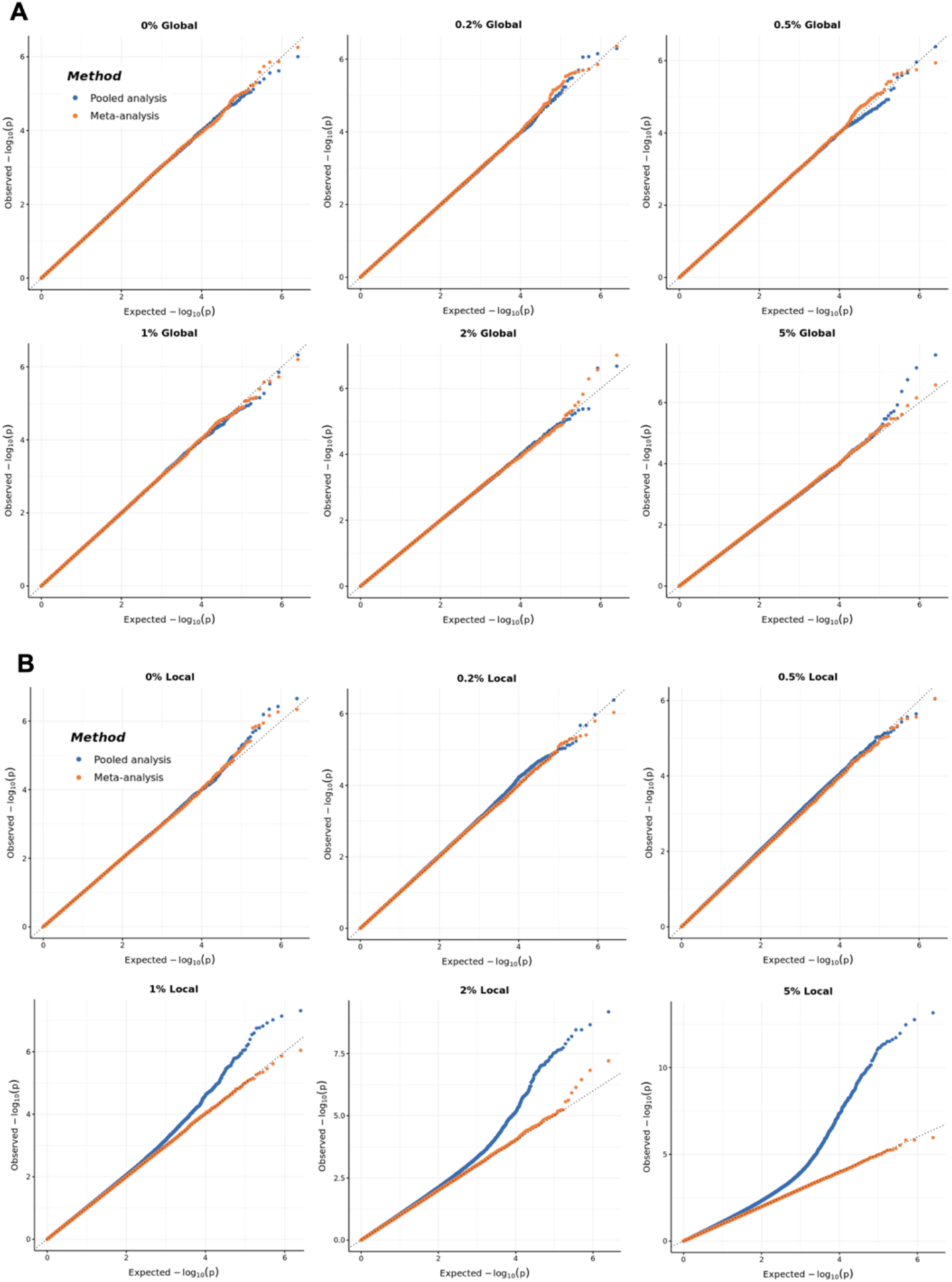
Quantile-Quantile (QQ) plots comparing the observed versus expected −log_10_(p) values for meta-analysis (orange) and pooled analysis (blue) under varying levels of population structure. Null phenotypes were generated using the All of Us dataset, which includes individuals from six genetic ancestry groups: 47,207 African (AFR), 36,500 Admixed American (AMR), 5,153 East Asian (EAS), 115,701 European (EUR), 497 Middle Eastern (MID) and 2,247 South Asian (SAS) (see **Methods**). Panel A shows QQ plots of mixed-effect GWAS results for null phenotypes with varying percentages (0%-5%) of phenotypic variance are explained by the first ten cross-ancestry-group principal components (PCs). Panel B illustrate the same analysis ancestry-group-specific PCs, with similar ranges of variance explained. Divergence from the diagonal line indicates p-values inflation due to population structure in both local and global population stratification scenarios.

### Statistical Power Simulations

#### Continuous phenotypes

To assess statistical power across different ancestry-group proportions, we considered three scenarios: (1) equal sample sizes across ancestries while increasing total sample size, (2) a fixed EUR sample size while gradually increasing non-EUR sample size, and (3) a fixed total sample size with increasing non-EUR sample sizes. In all scenarios, pooled analysis consistently outperformed meta-analysis and MR-MEGA (**Figure 4**).

**Figure 4:**
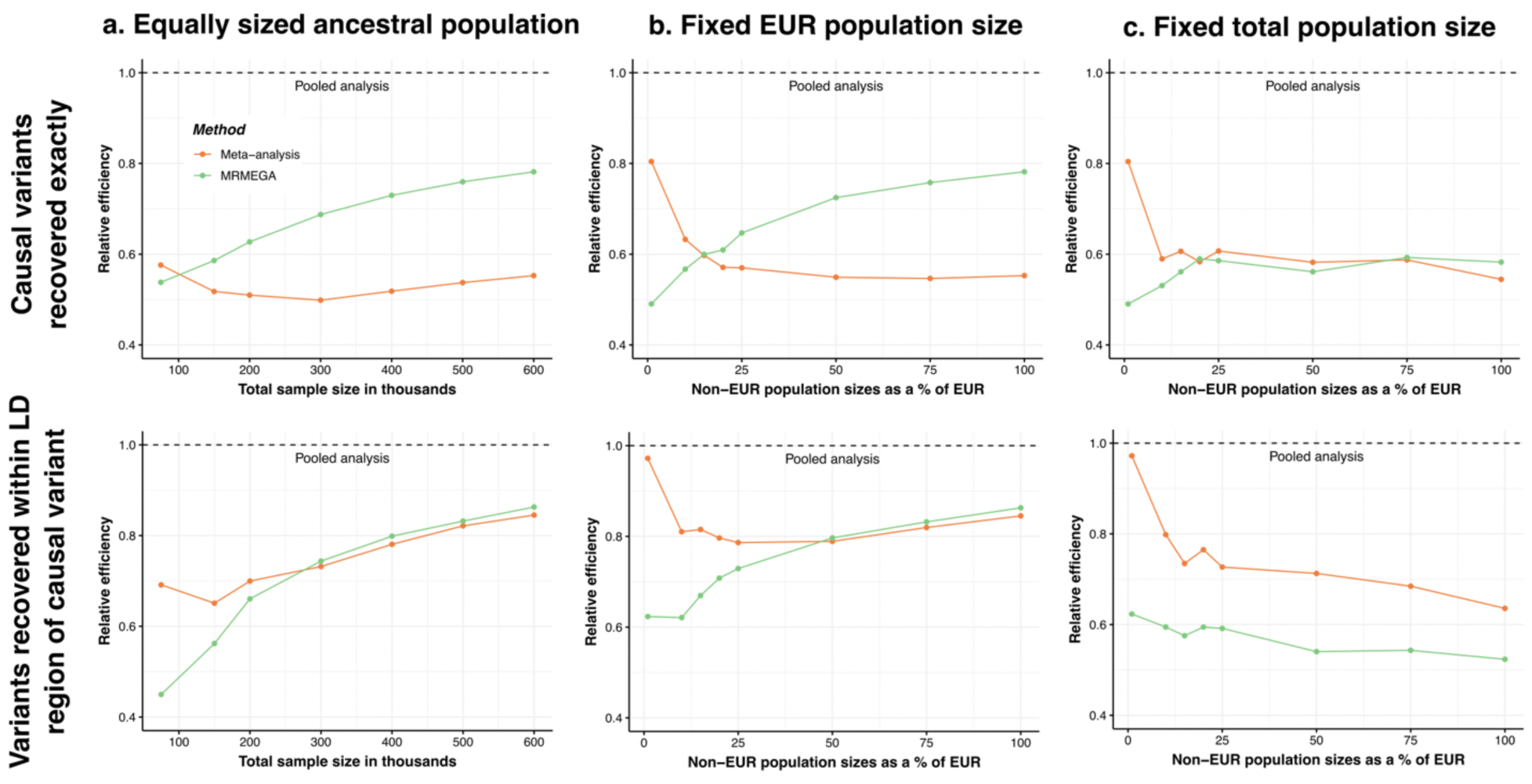
Comparison of meta-analysis and MR-MEGA with pooled analysis for detecting causal variants and variants in linkage disequilibrium (LD) across different ancestry-group proportions. This figure compares the relative efficiency of meta-analysis (orange) and MR-MEGA (green) in recovering causal variants and variants in LD, using pooled analysis (dashed line) as the benchmark (relative efficiency = 1.0). The top row (first panel) evaluates the exact recovery of causal variants, measured as the relative efficiency of identifying genome-wide significant (𝑃 ≤ 5 × 10^−8^) causal variants. The bottom row (second panel) assesses LD-based recovery of causal variants, which measures the relative efficiency of detecting causal variants that have at least one genome-wide significant variant within 500 kb. Each column represents a different study design: (a) equally sized ancestry groups (left), (b) a fixed EUR sample size of 120,000 with varying non-EUR sample sizes (middle), and (c) a fixed total sample size of 124,800 with varying non-EUR proportions (right). All methods are evaluated using GWAS summary statistics generated via REGENIE from simulations of continuous traits in a multi-ancestry dataset composed of African (AFR), Admixed American (AMR), East Asian (EAS), European (EUR), and South Asian (SAS) groups.

In the first scenario, where sample sizes were equal across ancestry groups (**Figure 4a**), meta-analysis initially outperformed MR-MEGA at smaller sample sizes, likely due to the additional parameter fitting required in MR-MEGA. However, once the total sample size exceeded 100,000 individuals (∼20,000 per ancestry across five ancestry groups), MR-MEGA surpassed meta-analysis in identifying causal variants. For LD-based detection (i.e., identifying variants within 500 kb of a causal SNP), MR-MEGA outperformed meta-analysis when each ancestry group had at least 56,000 individuals (total 𝑛 = 280,000).

In the second scenario, we fixed at 120,000 individuals, while gradually increasing non-EUR sample sizes (**Figure 4b**). This setup reflects the impact of recruiting non-EUR participants within an existing EUR-dominated biobank. When non-EUR groups reached approximately 20% of the EUR sample size (i.e., 120,000 EUR and 24,000 of each non-EUR ancestry, total 𝑛 = 216,000), MR-MEGA outperformed meta-analysis in LD-based region detection. This advantages of MR-MEGA became more pronounced when non-EUR groups reached approximately 50% of the EUR sample size.

For the third scenario, we maintained a fixed total sample size of 124,800 individuals, while varying non-EUR proportions (**Figure 4c**). Here, meta-analysis consistently outperformed MR-MEGA for LD-based detection, regardless of ancestry-group proportions. However, MR-MEGA showed a slight advantage in identifying causal variants when all five ancestry-group were equally represented (24,960 per ancestry group). Pooled analysis exhibited the highest statistical power across all ancestry-group compositions particularly when ancestry-group proportions were more balanced. This advantage was most evident in LD-based region detection, where pooled analysis consistently outperformed both meta-analysis and MR-MEGA.

Across all scenarios, pooled analysis demonstrated the highest statistical power, regardless of ancestry-group proportions or total sample size. When only ancestry-group-specific summary statistics were available, MR-MEGA performed best when each ancestry group had at least 20,000 individuals, while meta-analysis remained preferable when some ancestry groups were smaller.

#### Binary Phenotypes

The results for binary phenotypes were consistent with those observed for continuous phenotypes (**Table 1**). Simulations were conducted using an average of 237,241 cases and 237,241 controls, with detailed sample sizes provided in **Supplementary Table 3**. Across all evaluated metrics, pooled analysis exhibited the highest statistical power. When measuring power by the proportion of causal variants reaching genome-wide significance (𝑃 ≤ 5 × 10^−8^) pooled analysis identified 0.33% of causal variants, compared to 0.17% for meta-analysis and 0.19% for MR-MEGA. Similarly, for LD-based detection, pooled analysis achieved a power of 27.19%, exceeding both meta-analysis (21.25%) and MR-MEGA (19.18%).

**Table 1:** Power comparison of multi-ancestry GWAS approaches on simulated binary phenotypes. This table compares the performance of pooled analysis, meta-analysis and MR-MEGA, under mixed-effect modelling. The simulated dataset comprises of 237,241 cases and 237,241 controls from five ancestry groups: African (AFR), Admixed American (AMR), East Asian (EAS), European (EUR), and South Asian (SAS) populations. Detailed sample size by ancestry group is provided in **Supplementary Table 3**. The number of significant variants is determined using a genome-wide significance threshold of 𝑃 ≤ 5 × 10^−8^. Power 1 represents the percentage of causal variants recovered as GWAS-significant variants. Power 2 represents the percentage of causal variants recovered within a 500kb region of GWAS-significant variants. The number of significant variants, Power 1, and Power 2 are averaged over 10 replicates. Further details on the simulation setup for phenotype generation can be found in the **Material and Methods** section.

|  | No. samples<br>on average | No. variants<br>in GWAS<br>output | No. significant<br>variants | Power 1 | Power 2 |
| --- | --- | --- | --- | --- | --- |
| Pooled analysis | 474,482 | 2,017,677 | 4,567 | 0.33% | 27.19% |
| Meta-analysis | 474,482 | 2,024,717 | 2,375 | 0.17% | 21.25% |
| MR-MEGA | 474,482 | 1,304,330 | 2,551 | 0.19% | 19.18% |

#### Admixed Individuals

To assess statistical power in samples including admixed individuals, we simulated four groups: two reference groups (100% AFR and 100% EUR) and two groups of admixed individuals (50%/50% AFR/EUR and 80%/20% AFR/EUR) using the Admix-kit^25^ pipeline, each with 60,000 subjects (**Material and Methods**). Across all groups, pooled analysis demonstrated the highest power for both detecting causal SNPs and identifying variants within LD-based causal regions (**Table 2**). For exact causal variant detection, pooled analysis achieved a power of 1.37%, outperforming both meta-analysis (0.83%) and MR-MEGA (0.91%). For LD-based detection, pooled analysis reached 29.93%, exceeding meta-analysis (24.31%) and MR-MEGA (23.56%).

**Table 2:** Power comparison of multi-ancestry GWAS approaches on simulated continuous phenotypes in equally sized samples of admixed individuals. This table compares the performance of three multi-ancestry GWAS approaches: pooled analysis, meta-analysis and MR-MEGA, under mixed-effect modelling. The dataset comprises of one set of 60,000 samples simulated based on 1000G European (EUR) genotypes and one set of 60,000 samples simulated based on 1000G African (AFR) genotypes, one set of 60,000 individuals with 50/50 EUR/AFR admixture, and one set of 60,000 individuals with 20/80 EUR/AFR admixture. Simulations were performed using Admix-kit^25^. The number of significant variants was determined using a genome-wide significance threshold of 𝑃 ≤ 5 × 10^−8^. Power 1 represents the percentage of causal variants recovered as GWAS-significant variants. Power 2 represents the percentage of causal variants recovered within a 500kb of GWAS-significant variants. The number of significant variants, Power 1, and Power 2 are averaged over ten replicates. Further details on the simulation setup for phenotype generation can be found in the **Material and Methods** section.

|  | No. samples | No. variants in GWAS output | No. significant variants | Power 1 | Power 2 |
| --- | --- | --- | --- | --- | --- |
| Pooled analysis | 240,000 | 1,052,000 | 4,062 | 1.37% | 29.93% |
| Meta-analysis | 240,000 | 1,052,000 | 2,633 | 0.83% | 24.31% |
| MR-MEGA | 240,000 | 1,011,016 | 3,044 | 0.91% | 23.56% |

### Real Data Analysis in AoU and UKB

We conducted GWAS using mixed-effect modelling with REGENIE and fixed-effect modelling with PLINK2 on real data from AoU and UKB, analyzing eight continuous and five binary traits (**Materials and Methods**). Sample sizes for each trait within ancestry groups are provided in **Supplementary Tables 5-6**, while Manhattan and Q-Q plots for all GWAS results are shown in **Supplementary Figures 6-9**. To assess inflation in test statistics, we examined the scaled genomic inflation factor (𝜆_1000_), which adjusts for differences in sample size. Across all analyses, 𝜆_1000_ ranged from 0.999 to 1.003, indicating no or little systematic inflation was present. Since the true causal variants in real data are unknown, we measured statistical power by the number of genome-wide significant variants detected (𝑃 ≤ 5 × 10^−8^) . Specifically, we reported both the total number of genome-wide significant variants and the number of independent genome-wide significant variants after LD-clumping.

Under both mixed- and fixed-effect modelling, pooled analysis consistently identified the highest number of significant variants, particularly for highly polygenic traits such as height. Across both AoU (**Table 3, Supplementary Table 7**) and UKB (**Table 4, Supplementary Table 8**), pooled analysis outperformed both meta-analysis and MR-MEGA for all traits, in both continuous and binary outcomes. On average, across all analysed traits in UKB, pooled analysis detected 8% and 60% more independent genome-wide significant variants than meta-analysis and MR-MEGA, respectively. The advantage of pooled analysis was even more pronounced in AoU, where it identified, on average across all traits, 63% and 650% more independent significant variants than meta-analysis and MR-MEGA, respectively. This greater advantage of pooled analysis in AoU is driven by its larger proportion of non-European participants (∼41% non-EUR) compared to UKB (∼4% non-EUR). This pattern aligns with our theoretical derivations (**Materials and Methods**), which predict that when MAFs vary across ancestries, pooled analysis has a greater advantage over meta-analysis when ancestry proportions are more balanced.

**Table 3:** Comparison of significant and independent variants across multi-ancestry GWAS methods in All of Us with mixed-effect modelling. This table compares the number of significant and independent variants identified using three different multi-ancestry GWAS approaches: pooled analysis, meta-analysis and MR-MEGA. Variants were filtered based on a minor allele frequency (MAF) > 0.01 in at least one ancestry group, resulting in approximately 1.2 million variants analyzed. Significant variants were determined using a genome-wide significance threshold of 𝑃 ≤ 5 × 10^−8^. Independent variants are generated with linkage disequilibrium (LD) clumping with an r^2^ threshold of 0.1. The eight continuous phenotypes include height, waist circumference, low-density lipoproteins cholesterol (LDL), total cholesterol (TC), high-density lipoproteins cholesterol (HDL), calcium, creatinine, estimated glomerular filtration rate (eGFR). The five binary phenotypes include asthma, coronary artery disease (CAD), type II diabetes (T2D), breast cancer and prostate cancer. The number of subjects varies by phenotype, and the sample size for each trait is provided in **Supplementary Table 5**.

|  |  | Height | Waist | LDL | TC | HDL | Calcium | Creatinine | eGFR | Asthma | CAD | T2D | Breast cancer | Prostate cancer |
| --- | --- | --- | --- | --- | --- | --- | --- | --- | --- | --- | --- | --- | --- | --- |
| Number of significant variants | Pooled analysis | 5,435 | 2,120 | 354 | 921 | 1,258 | 73 | 513 | 37 | 101 | 125 | 268 | 20 | 35 |
|  | Meta-analysis | 4,668 | 823 | 339 | 550 | 760 | 62 | 73 | 11 | 99 | 124 | 287 | 20 | 35 |
|  | MR-MEGA | 3,695 | 627 | 303 | 475 | 628 | 31 | 25 | 3 | 40 | 120 | 280 | 20 | 22 |
| Number of independent significant variants | Pooled analysis | 580 | 316 | 42 | 50 | 101 | 18 | 213 | 1 | 7 | 3 | 28 | 2 | 11 |
|  | Meta-analysis | 559 | 80 | 36 | 39 | 87 | 15 | 43 | 1 | 7 | 3 | 24 | 2 | 8 |
|  | MR-MEGA | 394 | 44 | 22 | 27 | 52 | 6 | 3 | 1 | 4 | 2 | 16 | 2 | 5 |

**Table 4:** Comparison of significant and independent variants across multi-ancestry GWAS methods in the UK Biobank with mixed-effect modelling. This table compares the number of significant and independent variants identified using three multi-ancestry GWAS approaches: pooled analysis, meta-analysis and MR-MEGA. Variants were filtered using a MAF threshold of > 0.01 within at least one ancestry group, resulting in approximately 1.53 million variants analyzed. Significant variants were determined using a genome-wide significance threshold of 𝑃 ≤ 5 × 10^−8^. Independent variants are generated with linkage disequilibrium (LD) clumping with an 𝑟^2^ threshold of 0.1. The eight continuous phenotypes include height, waist circumference, low-density lipoproteins cholesterol (LDL), total cholesterol (TC), high-density lipoproteins cholesterol (HDL), calcium, creatinine, estimated glomerular filtration rate (eGFR). The five binary phenotypes include asthma, coronary artery disease (CAD), type II diabetes (T2D), breast cancer and prostate cancer. The number of subjects varies by phenotype, and sample sizes for each trait are provided in **Supplementary Table 6**.

|  |  | Height | Waist | LDL | TC | HDL | Calcium | Creatinine | eGFR | Asthma | CAD | T2D | Breast cancer | Prostate cancer |
| --- | --- | --- | --- | --- | --- | --- | --- | --- | --- | --- | --- | --- | --- | --- |
| Number of significant variants | Pooled analysis | 44,509 | 5,393 | 6,580 | 7,903 | 10,132 | 3,758 | 3,582 | 14,174 | 1,010 | 752 | 742 | 207 | 205 |
|  | Meta-analysis | 45,001 | 5,196 | 6,532 | 7,792 | 9,624 | 3,750 | 3,664 | 12,738 | 1,039 | 675 | 745 | 211 | 186 |
|  | MR-MEGA | 33,844 | 3,629 | 4,743 | 5,884 | 6,871 | 2,690 | 2,578 | 9,054 | 791 | 453 | 509 | 135 | 125 |
| Number of independent significant variants | Pooled analysis | 2,576 | 351 | 345 | 401 | 604 | 271 | 378 | 743 | 37 | 43 | 52 | 15 | 28 |
|  | Meta-analysis | 2,574 | 338 | 344 | 399 | 595 | 270 | 232 | 731 | 34 | 41 | 53 | 14 | 25 |
|  | MR-MEGA | 1,851 | 224 | 232 | 266 | 404 | 184 | 164 | 505 | 26 | 24 | 32 | 11 | 15 |

Additionally, mixed-effect models consistently outperformed fixed-effect models in detecting genome-wide significant variants. Across all three GWAS approaches, the mixed-effect model identified 6% more independent significant variants than the fixed-effect model in AoU (**Table 3, Supplementary Table 7**) and 18% more in UKB (**Table 4, Supplementary Table 8**). This increased statistical power of mixed-effect models is likely due to their ability to adjust for both known covariates (top PCs, age, and sex) through fixed effects and additional genomic structure through random effects. By modelling these sources of variation, mixed-effect models reduce residual error variance, thereby increasing statistical power.

## Discussion

We evaluated three multi-ancestry GWAS approaches, pooled analysis, meta-analysis, and MR-MEGA, under both fixed- and mixed-effect modelling. Our findings show that pooled analysis consistently provides the highest statistical power across various ancestry-group compositions and trait architectures, while maintaining well-controlled type I error in most realistic scenarios. This advantage is particularly pronounced when allele frequencies vary across ancestry groups, supporting our theoretical derivation that pooled analysis benefits from increased minor allele carrier counts across populations.

Through large-scale simulations, we showed that meta-analysis and MR-MEGA are effective but less powerful than pooled analysis, particularly when non-EUR individuals made up a larger proportion of the sample (**Figure 4c**). Our theoretical framework explains this disparity: as allele frequency divergence increases, meta-analysis becomes less efficient compared to pooled analysis. Real data analyses from UKB (𝑛 ≈ 324,000) and AoU (𝑛 ≈ 207,000) further validated our findings, with pooled analysis detecting the most independent genome-wide significant variants across both continuous and binary traits (**Tables 3-4, Supplementary Tables 7-8**).

Beyond allele frequency differences, sample size distribution across ancestry groups plays a key role in determining statistical power. When ancestry groups are more balanced, the power advantage of pooled analysis over meta-analysis and MR-MEGA is maximized. This is particularly evident in AoU, where a higher proportion of non-EUR participants amplifies the advantage of pooled analysis, further reinforcing our theoretical predictions.

Our findings also highlight the higher power of mixed-effect models over fixed-effect models (**Tables 3-4, Supplementary Tables 7-8**). By incorporating random effects, mixed-effect models account for genome-wide polygenic effects, which helps reduce residual error variance and improve the detection of true associations. Additionally, they adjust for cryptic relatedness and population structure, enhancing robustness while maintaining proper type I error control. These results support the continued adoption of efficient mixed-effect frameworks such as REGENIE for large-scale GWAS analyses.

Historically, fixed-effect meta-analysis has been the standard approach for multi-ancestry GWAS due to its ability to combine summary statistics from independent studies without requiring individual-level data^8,10,11,33,34^. However, our results suggest that pooled analysis is preferable when individual-level data are available, as it avoids binning individuals into discrete ancestry groups and leverages shared genetic architecture across populations. MR-MEGA, which models allele frequency differences, performed slightly better than standard meta-analysis when non-EUR sample sizes were sufficiently large, but it still lagged behind pooled analysis in most scenarios.

An important consideration is that pooled analysis inherently assumes homogeneous effect sizes across ancestry groups. While recent studies suggest that most genetic effects are shared across populations^35,36^, certain variants may exhibit ancestry-group-specific effect heterogeneity^37^, where meta-analysis might be more appropriate. Future methodological advancements, such as multi-trait^38^ or ancestry-aware fine-mapping approaches^39^, could refine pooled analysis to better capture ancestry-specific effects.

The observed power differences have important implications for PRS development and fine-mapping efforts in multi-ancestry cohorts. Pooled analysis may improve PRS portability by increasing statistical power for variant discovery, ultimately enhancing risk prediction models in different populations^40,41^. Additionally, the superior power of pooled analysis suggests that functional fine-mapping and transcriptome-wide association studies may benefit from using pooled GWAS summary statistics, as these analyses rely heavily on accurate effect size estimates^42^. Another key implication is for admixture mapping studies. Our findings indicate that pooled analysis provides better power than meta-analysis when analyzing admixed individuals, supporting its use in large, diverse cohorts such as AoU and the Million Veteran Program^43^. While meta-analysis may still be necessary when individual-level data are unavailable, future work should explore hybrid approaches that combine pooled analysis with ancestry-aware meta-analysis.

While our study provides strong evidence for the advantages of pooled analysis, there are important limitations. First, pooled analysis requires individual-level genotype data, which is often restricted due to privacy concerns and regulatory limitations. This constraint continues to favor meta-analysis for large-scale cross-cohort collaborations, highlighting the need for novel meta-analytic methods that retain the power of pooled approaches. Second, our simulations assume homogeneous allelic effects across ancestries, which may not fully reflect complex gene-environment interactions or ancestry-group-specific selection pressures. Future work should explore adaptive models that account for effect heterogeneity while maintaining the power benefits of pooled analysis. Third, while we demonstrated that pooled analysis maintains well-controlled type I error in most realistic settings, caution is needed when ancestry-group-specific PCs explain a large proportion (>1%) of trait variance, as this may lead to spurious associations. To mitigate this, we recommend evaluating the percentage of variance explained by ancestry-group-specific PCs prior to running pooled GWAS, particularly for traits with strong population structure effects.

In summary, our study demonstrates that pooled analysis provides the highest statistical power for multi-ancestry GWAS, particularly when allele frequencies vary across populations. While meta-analysis remains a valuable approach when individual-level data are unavailable, pooled analysis should be prioritized when feasible. Our findings emphasize the importance of balanced ancestry representation and highlight the advantages of mixed-effect modelling for large-scale genetic studies. Future research should focus on developing hybrid approaches that integrate the strengths of pooled and meta-analysis, ensuring that multi-ancestry GWAS continue to uncover novel genetic insights across diverse populations.

## Data Availability

The simulated data for 600,000 subjects from five ancestries were downloaded from here: https://dataverse.harvard.edu/dataset.xhtml?persistentId=doi:10.7910/DVN/COXHAP. The UK Biobank phenotype and genotype data used in this study are available to registered researchers through the UKB data-access protocol and require an approved access application. All of Us phenotype and genotype data can be accessed through the All of Us Research Workbench (https://workbench.researchallofus.org/). All data used in this study are available to registered researchers with controlled-tier access through the All of Us data-access protocol.

## Acknowledgements

The analysis utilized the high-performance computation Faculty of Arts and Sciences Research Computing Cluster at Harvard University, Biowulf cluster at National Institutes of Health, USA, Polaris cluster at Argonne National Laboratories, USA, and All of Us Researcher Workbench. We gratefully acknowledge All of Us participants for their contributions, without whom this research would not have been possible. We also thank the National Institutes of Health’s All of Us Research Program for making available the participant data examined in this study. We also acknowledge the UK Biobank, a major biomedical database, along with their participants. This research has been conducted using the UK Biobank Resource under Application Number 52008. This work is funded by: NIH Research Grant U01CA261339 (J.A.D.), NIH Training Grant T32GM135117 and NSF Graduate Research Fellowship DGE-2140743 (T.C.), NIH intramural research funding (H.X., X.W., P.K and H.Z.).

## Author contributions

J.A.D., P.K. and H.Z. conceived the project. J.A.D carried out all data analyses and derivations, simulated admixed genotypes, ran GWAS on AoU data using the All of Us research workbench and ran GWAS on UKB data with supervision from P.K. and H.Z..

T.C. advised on best practices when running cluster analyses and pre-processed genotype and phenotype data for UKB analyses. J.A.D. and X.H. pre-processed the AoU data. X.W. advised on methods for binary analyses. R.M. and A.R. provided computing resources from Argonne National Laboratory to simulate genotypes and phenotypes. J.A.D., P.K. and H.Z. drafted the manuscript. T.C., X.H., X.W., A. A. R., R.

K. M. provided comments. All co-authors reviewed and approved the final version of the manuscript.

## Code Availability

Simulation and data analyses code is available at: GitHub (https://github.com/juliealexia/multiancestryGWAS)

Tutorial and software implementing Admix-kit is available at: GitHub (https://github.com/KangchengHou/admix-kit)

Tutorial and software implementing REGENIE is available at: GitHub (https://rgcgithub.github.io/regenie/install)

MR-MEGA: https://tools.gi.ut.ee/tools/MR-MEGA_v0.2.zip PLINK2: https://www.cog-genomics.org/plink/2.0/

The majority of our statistical analyses are performed using the following R packages: ggplot2 version 3.5.1, dplyr version 1.1.4, data.table version 1.16.2, magrittr version 2.0.3, mvtnorm version 1.3-3, stringr version 1.5.1, tidyr version 1.3.1, qqman version 0.1.9, ggthemes version 5.1.0, RColorBrewer version 1.1-3, boot version 1.3-31

## References

1. Sollis, E. et al. The NHGRI-EBI GWAS Catalog: knowledgebase and deposition resource. Nucleic Acids Res 51, (2023).

2. Buniello, A. et al. The NHGRI-EBI GWAS Catalog of published genome-wide association studies, targeted arrays and summary statistics 2019. Nucleic Acids Res 47, (2019).

3. Mills, M. C. & Rahal, C. The GWAS Diversity Monitor tracks diversity by disease in real time. Nature Genetics vol. 52 (2020).

4. Carlson, C. S. et al. Generalization and Dilution of Association Results from European GWAS in Populations of Non-European Ancestry: The PAGE Study. PLoS Biol 11, (2013).

5. Ju, D., Hui, D., Hammond, D. A., Wonkam, A. & Tishkoff, S. A. Importance of Including Non-European Populations in Large Human Genetic Studies to Enhance Precision Medicine. Annual Review of Biomedical Data Science vol. 5 (2022).

6. Mägi, R. et al. Trans-ethnic meta-regression of genome-wide association studies accounting for ancestry increases power for discovery and improves fine-mapping resolution. Hum Mol Genet 26, (2017).

7. Kim, J. J. et al. Multi-ancestry genome-wide association meta-analysis of Parkinson’s disease. Nat Genet 56, (2024).

8. Shrine, N. et al. Multi-ancestry genome-wide association analyses improve resolution of genes and pathways influencing lung function and chronic obstructive pulmonary disease risk. Nat Genet 55, (2023).

9. Ishigaki, K. et al. Multi-ancestry genome-wide association analyses identify novel genetic mechanisms in rheumatoid arthritis. Nat Genet 54, (2022).

10. Mahajan, A. et al. Multi-ancestry genetic study of type 2 diabetes highlights the power of diverse populations for discovery and translation. Nat Genet 54, (2022).

11. Graham, S. E. et al. The power of genetic diversity in genome-wide association studies of lipids. Nature 600, (2021).

12. Kachuri, L. et al. Principles and methods for transferring polygenic risk scores across global populations. Nature Reviews Genetics vol. 25 (2024).

13. Zhang, H. et al. A new method for multiancestry polygenic prediction improves performance across diverse populations. Nat Genet 55, (2023).

14. Jin, J. et al. MUSSEL: Enhanced Bayesian polygenic risk prediction leveraging information across multiple ancestry groups. Cell genomics 4, 100539 (2024).

15. Zhang, J. et al. An ensemble penalized regression method for multi-ancestry polygenic risk prediction. Nat Commun 15, 3238 (2024).

16. Ruan, Y. et al. Improving polygenic prediction in ancestrally diverse populations. Nat Genet 54, 573–580 (2022).

17. Hoggart, C. J. et al. BridgePRS leverages shared genetic effects across ancestries to increase polygenic risk score portability. Nat Genet 56, 180–186 (2024).

18. Willer, C. J., Li, Y. & Abecasis, G. R. METAL: fast and efficient meta-analysis of genomewide association scans. Bioinformatics 26, 2190–1 (2010).

19. Chang, C. C. et al. Second-generation PLINK: Rising to the challenge of larger and richer datasets. Gigascience 4, (2015).

20. Mbatchou, J. et al. Computationally efficient whole-genome regression for quantitative and binary traits. Nat Genet 53, 1097–1103 (2021).

21. Jiang, L. et al. A resource-efficient tool for mixed model association analysis of large-scale data. Nat Genet 51, (2019).

22. Chen, H. et al. Control for Population Structure and Relatedness for Binary Traits in Genetic Association Studies via Logistic Mixed Models. Am J Hum Genet 98, (2016).

23. Zhou, W. et al. Efficiently controlling for case-control imbalance and sample relatedness in large-scale genetic association studies. Nat Genet 50, (2018).

24. Gogarten, S. M. et al. Genetic association testing using the GENESIS R/Bioconductor package. Bioinformatics 35, (2019).

25. Hou, K. et al. Admix-kit: an integrated toolkit and pipeline for genetic analyses of admixed populations. Bioinformatics 40, (2024).

26. All of Us Research Program Genomics Investigators. Genomic data in the All of Us Research Program. Nature 627, 340–346 (2024).

27. Bycroft, C. et al. The UK Biobank resource with deep phenotyping and genomic data. Nature 562, 203–209 (2018).

28. Su, Z., Marchini, J. & Donnelly, P. HAPGEN2: Simulation of multiple disease SNPs. Bioinformatics 27, (2011).

29. Auton, A. et al. A global reference for human genetic variation. Nature vol. 526 (2015).

30. Belmont, J. W. et al. The international HapMap project. Nature 426, (2003).

31. Michailidou, K. et al. Association analysis identifies 65 new breast cancer risk loci. Nature 551, (2017).

32. Bien, S. A. et al. Strategies for enriching variant coverage in candidate disease loci on a multiethnic genotyping array. PLoS One 11, (2016).

33. Wang, A. et al. Characterizing prostate cancer risk through multi-ancestry genome-wide discovery of 187 novel risk variants. Nat Genet 55, (2023).

34. Purdue, M. P. et al. Multi-ancestry genome-wide association study of kidney cancer identifies 63 susceptibility regions. Nat Genet 56, 809–818 (2024).

35. Hou, K. et al. Causal effects on complex traits are similar for common variants across segments of different continental ancestries within admixed individuals. Nat Genet 55, (2023).

36. Taylor, D. J. et al. Sources of gene expression variation in a globally diverse human cohort. Nature 632, (2024).

37. Hodonsky, C. J. et al. Ancestry-specific associations identified in genome-wide combined-phenotype study of red blood cell traits emphasize benefits of diversity in genomics. BMC Genomics 21, (2020).

38. Turley, P. et al. Multi-trait analysis of genome-wide association summary statistics using MTAG. Nat Genet 50, (2018).

39. Gao, B. & Zhou, X. MESuSiE enables scalable and powerful multi-ancestry fine-mapping of causal variants in genome-wide association studies. Nat Genet 56, (2024).

40. Martin, A. R. et al. Human Demographic History Impacts Genetic Risk Prediction across Diverse Populations. Am J Hum Genet 100, (2017).

41. Duncan, L. et al. Analysis of polygenic risk score usage and performance in diverse human populations. Nat Commun 10, (2019).

42. Gusev, A. et al. Integrative approaches for large-scale transcriptome-wide association studies. Nat Genet 48, (2016).

43. Verma, A. et al. Diversity and scale: Genetic architecture of 2068 traits in the VA Million Veteran Program. Science 385, eadj1182 (2024).

