## Supplementary Figures for "Evaluating Multi-Ancestry Genome-Wide Association Methods: Statistical Power, Population Structure, and Practical Implications"

**Supplementary Figure 1: Global PCA projection into five continental ancestries for simulated dataset onto PCs 1-2.** These PCs are defined by 109,335 SNPs pruned using plink2 --indep-pairwise using a step size of 1, a window of 500kb and a $r^2$ threshold of 0.05 into five continental ancestries shown in color dots. Represented ancestries are African [AFR], Admixed American [AMR], East Asian [EAS], European [EUR] and South Asian [SAS], each with 120,000 simulated individuals.

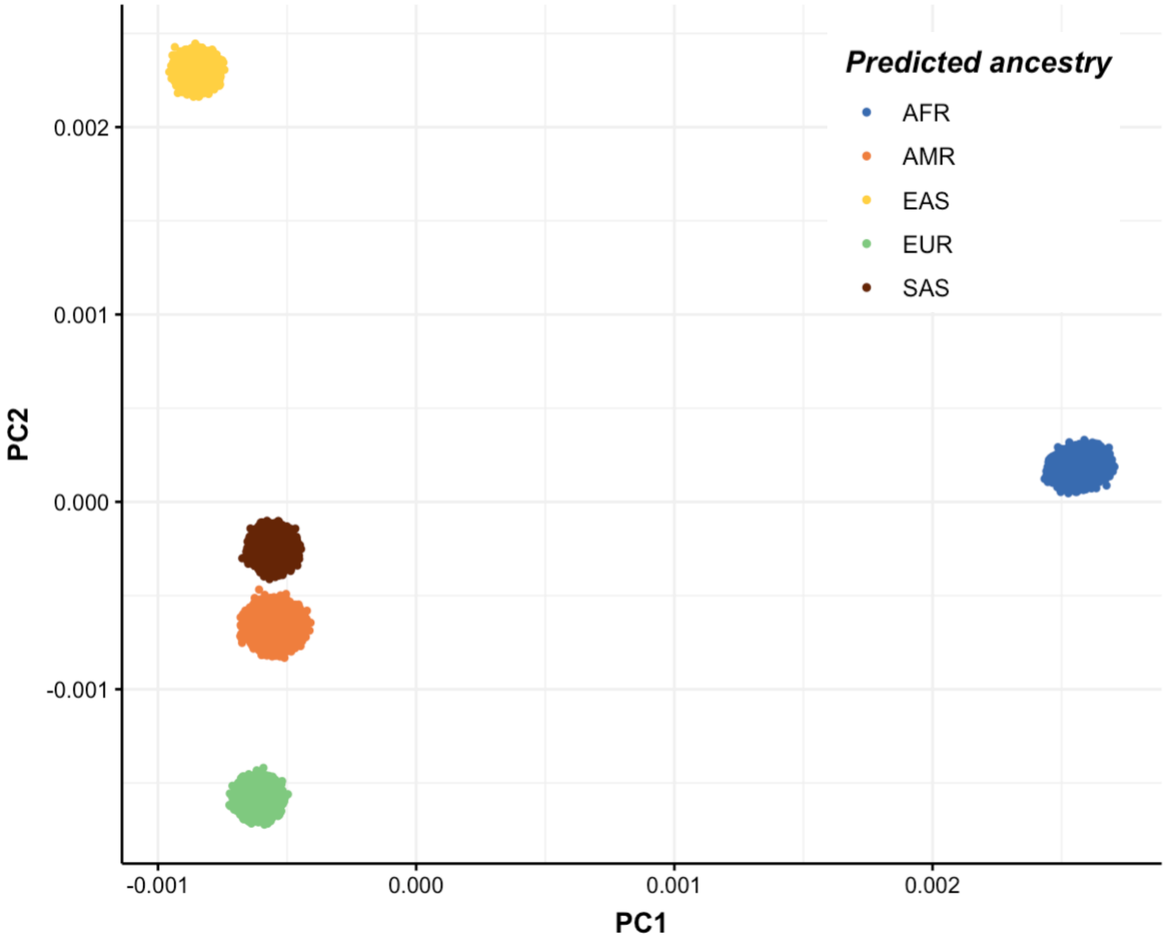

**Supplementary Figure 2: Global PCA projection into two continental ancestries and two admixed ancestries for admixed simulated dataset onto PCs 1-2.** These PCs are defined by 51,401 SNPs pruned using plink2 --indep-pairwise using a step size of 1, a window of 500kb and a $r^2$ threshold of 0.05 into four ancestries shown in color dots. The dataset comprises of one set of 60,000 samples simulated based on 1000 Genomes European (EUR) genotypes and one set of 60,000 samples simulated based on 1000 Genomes African (AFR) genotypes, one set of 60,000 individuals with 50/50 EUR/AFR admixture (ADMIX1), and one set of 60,000 individuals with 20/80 EUR/AFR admixture (ADMIX2). Simulations were performed using Admix-kit[25].

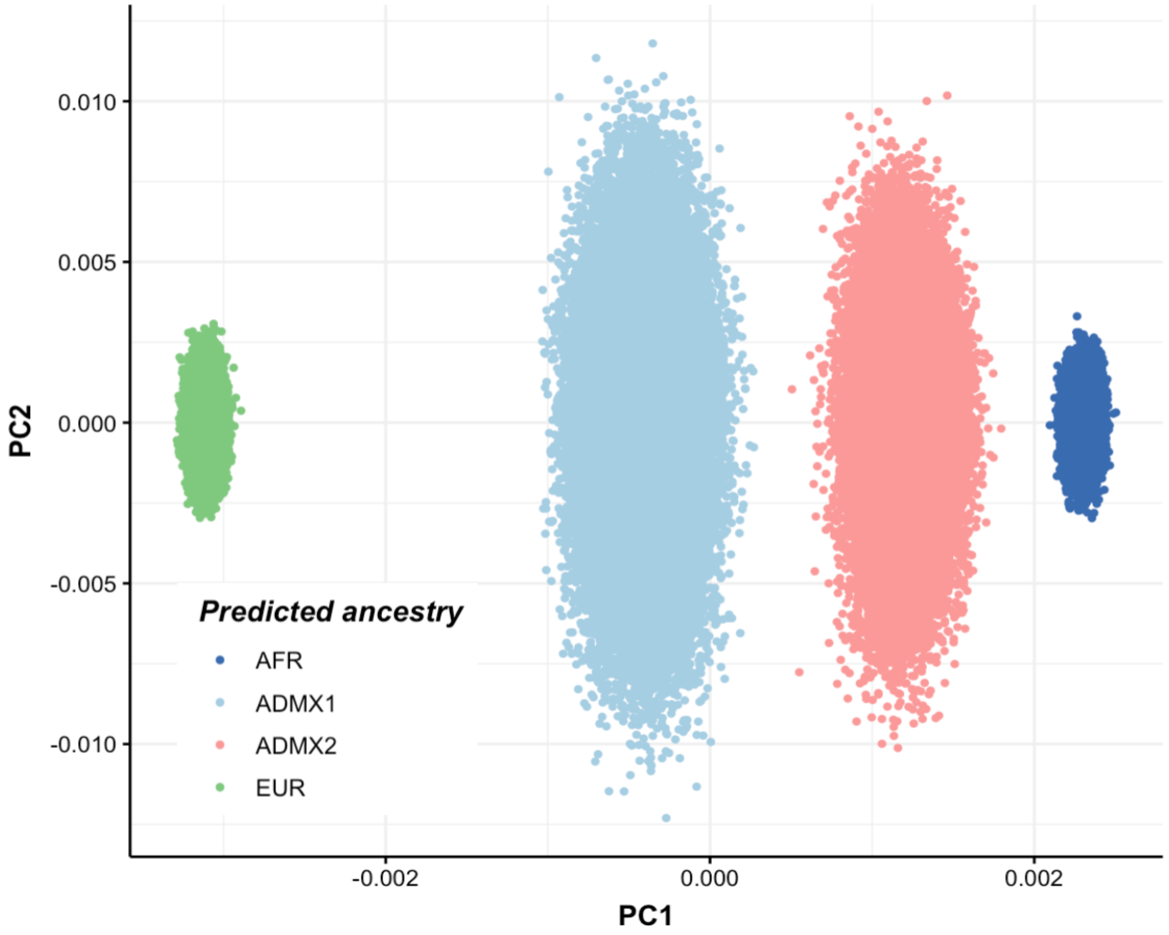

**Supplementary Figure 3: Global PCA projection of All of Us data into six continental ancestries onto PCs 1-2.** These PCs are defined by 125,692 SNPs pruned using plink2 --indep-pairwise using a step size of 1, a window of 500kb and a $r^2$ threshold of 0.1 into six continental ancestries shown in color dots. Represented are 47,207 African [AFR], 36,500 Admixed American [AMR], 5,153 East Asian [EAS], 115,701 European [EUR], 497 Middle Eastern [MID] and 2,247 South Asian [SAS].

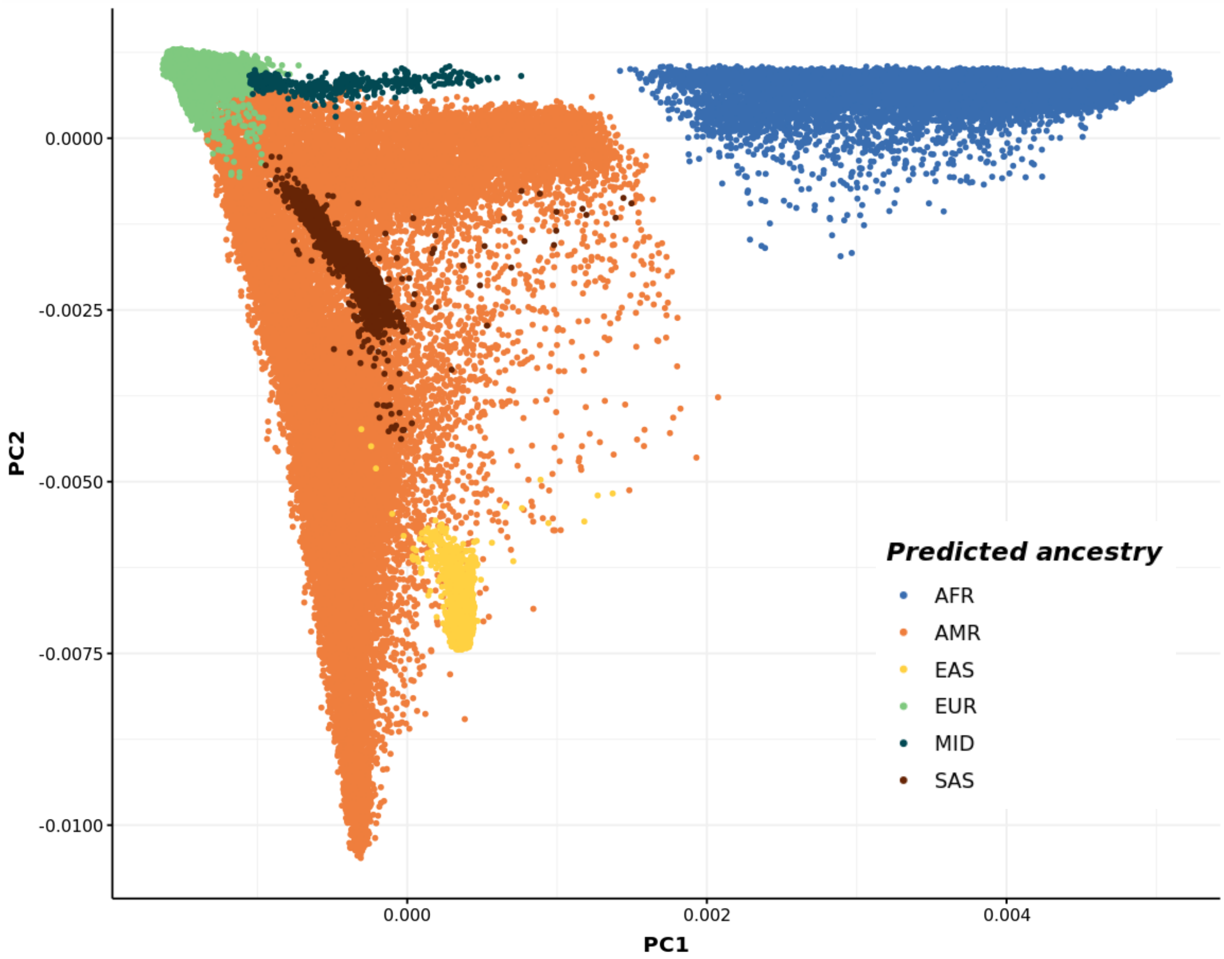

**Supplementary Figure 4: Global PCA projection of UK Biobank data into five continental ancestries onto PCs 1-2.** These PCs are defined by 51,922 SNPs pruned using plink2 –indep-pairwise using a step size of 0.1, a window of 500kb and a $r^2$ threshold of 0.05 into five continental ancestries shown in color dots. Represented are 6,864 African [AFR], 590 Admixed American [AMR], 586 East Asian [EAS], 311,053 European [EUR] and 5,734 South Asian [SAS].

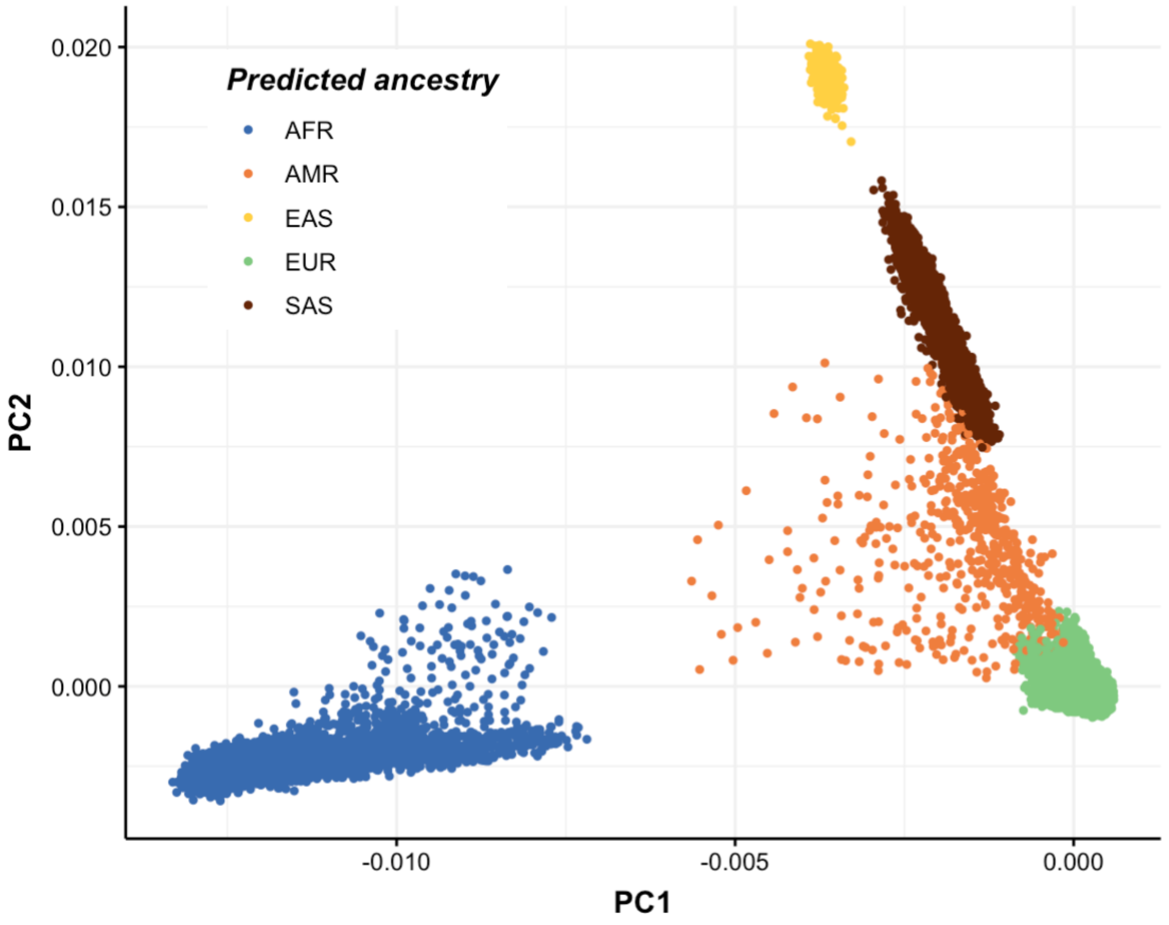

**Supplementary Figure 5: Quantile-Quantile (QQ) plots comparing the observed versus expected $-\log_{10}(p)$ values for meta-analysis (orange) and pooled analysis (blue) under varying levels of population structure**. Null phenotypes were generated using the All of Us dataset, which includes individuals from six genetic ancestry groups: African (AFR), Admixed American (AMR), East Asian (EAS), European (EUR), and South Asian (SAS). Panel A shows QQ plots of fixed-effect GWAS results for null phenotypes with varying percentages (0%-5%) of phenotypic variance are explained by the first 10 global (cross-ancestry) principal components (PCs). Panel B illustrate the same analysis for local (ancestry-specific) PCs, with similar ranges of variance explained.  Divergence from the diagonal line indicates p-values inflation due to population structure in both local and global population stratification scenarios.

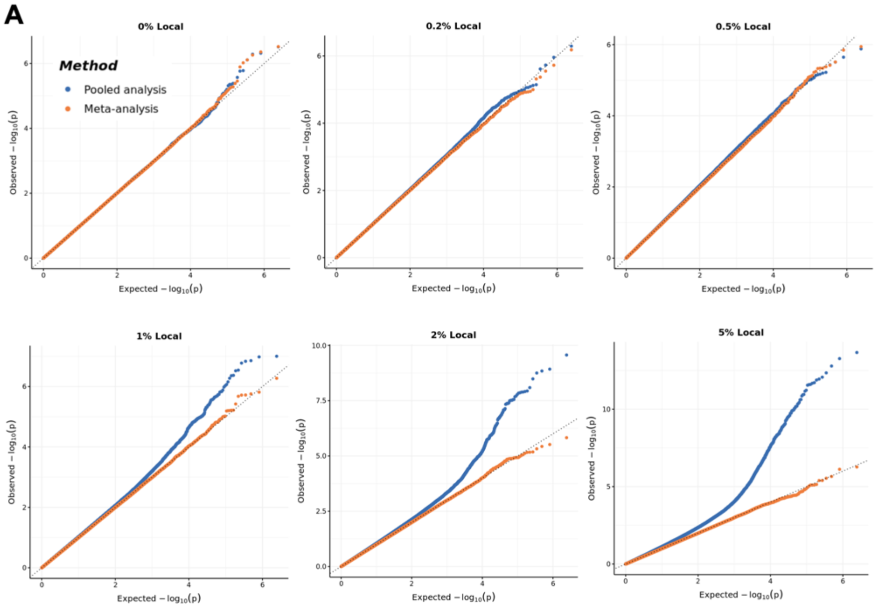


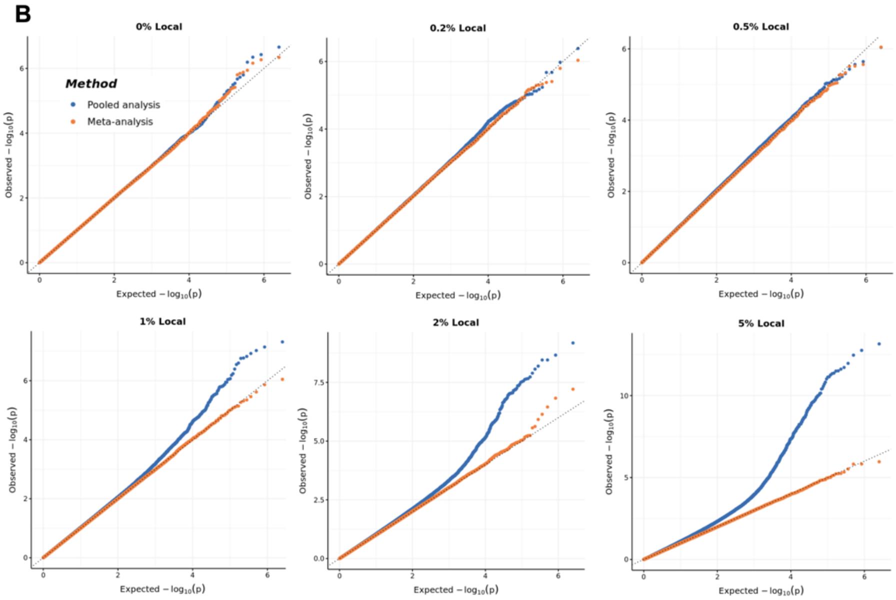

**Supplementary Figure 6-a: Manhattan plot and QQ plot[1] based on the All of Us multi-ancestry GWAS summary statistics for height in six populations (European, African American, Latino, East Asian, South Asian, Middle Eastern), obtained for pooled analysis, meta-analysis and MR-MEGA using mixed-effect modelling** (from top panel to bottom panel). The red, blue, green and purple shaded regions around the diagonal line in the QQ plots indicate the 95% confidence intervals expected under the null hypothesis of no association between genetic markers and the trait of interest, for minor allele frequencies (MAF) within the ranges (0.05, 0.5], (0.01, 0.05], (0.001, 0.005] and (0,0.001] respectively. Under the null hypothesis, the p-value follows a uniform (0,1) distribution. The jth order statistic follows a Beta (j, N-j+1) distribution, where N is the total number of variants given a specific MAF cutoff.

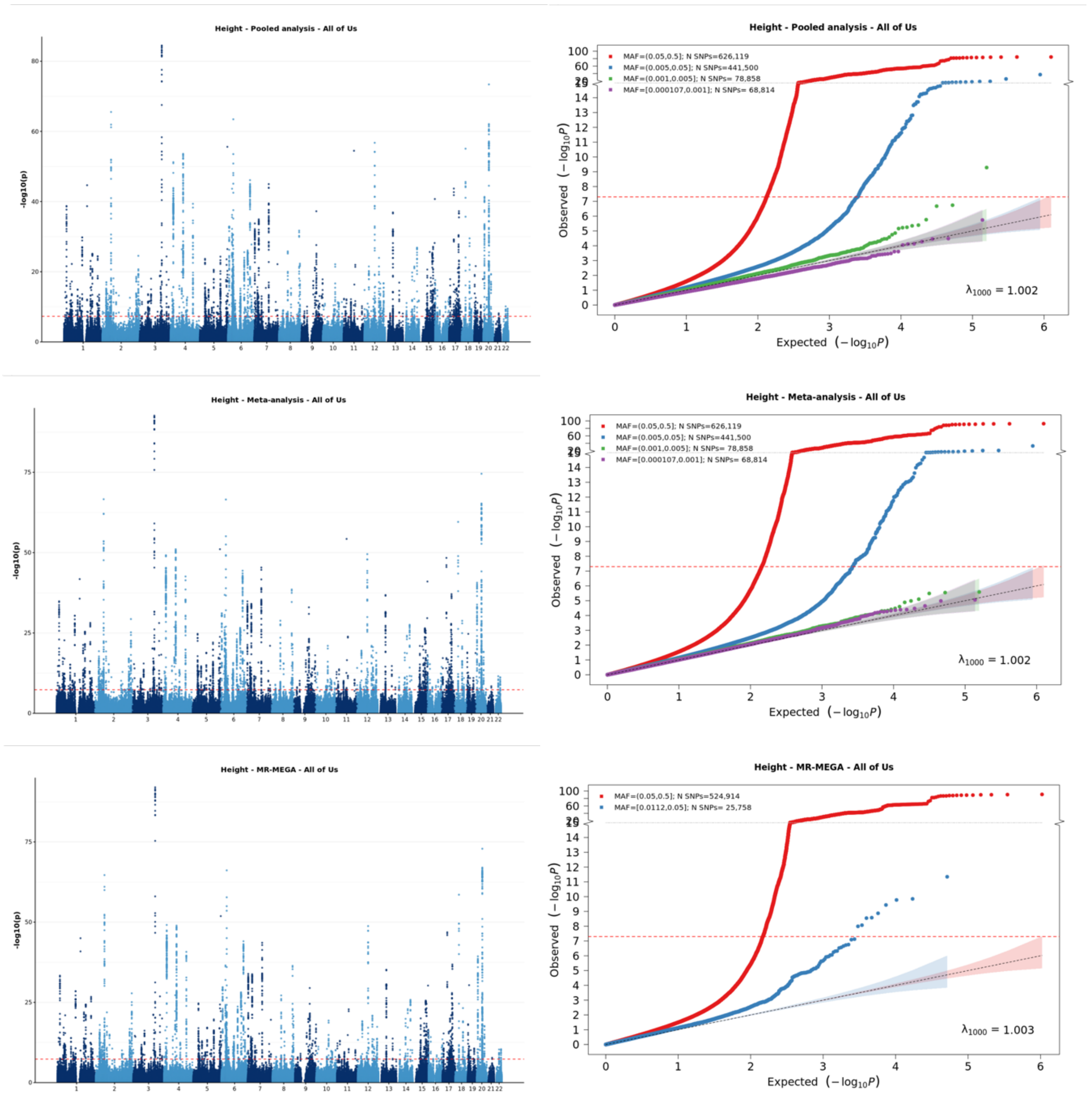


[1] For continuous traits, $\lambda_{1000}$ scales the genomic inflation factor $\lambda$ to a study with 1000 subjects using $\lambda_{1000} = 1 + 1000 * (\lambda - 1)/N$, where N is the total sample size. For binary traits, $\lambda_{1000}$ scales $\lambda$ to a study with 1000 cases and 1000 controls using $\lambda_{1000} = 1 + 1000 * (\lambda - 1) * (\frac{1}{N_{case}} + \frac{1}{N_{control}})$

**Supplementary Figure 6-b: Manhattan plot and QQ plot[1] based on the All of Us multi-ancestry GWAS summary statistics for waist circumference in six populations (European, African American, Latino, East Asian, South Asian, Middle Eastern), obtained for pooled analysis, meta-analysis and MR-MEGA using mixed-effect modelling** (from top panel to bottom panel). The red, blue, green and purple shaded regions around the diagonal line in the QQ plots indicate the 95% confidence intervals expected under the null hypothesis of no association between genetic markers and the trait of interest, for minor allele frequencies (MAF) within the ranges (0.05, 0.5], (0.01, 0.05], (0.001, 0.005] and (0,0.001] respectively. Under the null hypothesis, the p-value follows a uniform (0,1) distribution. The jth order statistic follows a Beta (j, N-j+1) distribution, where N is the total number of variants given a specific MAF cutoff.

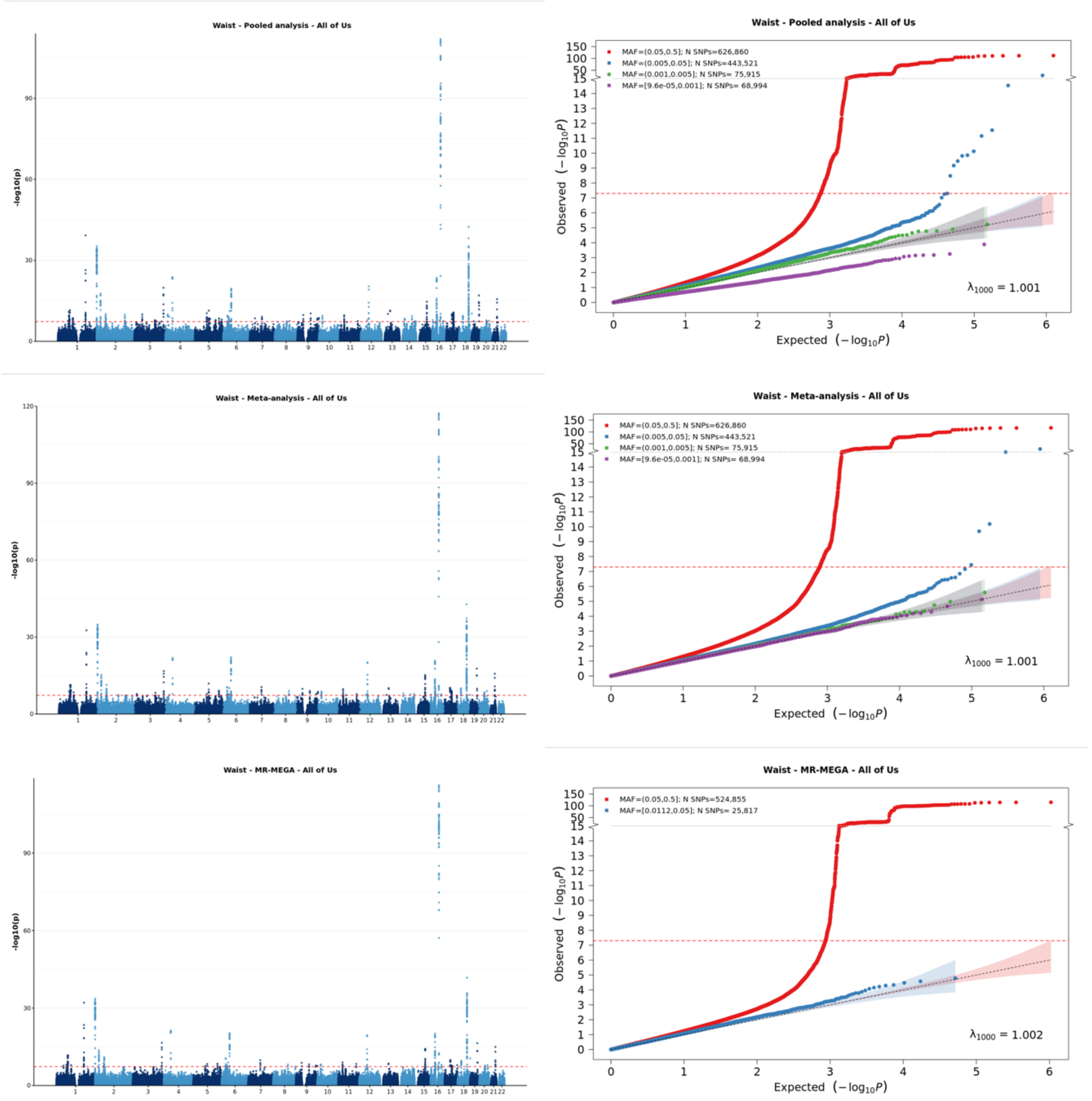


[1] For continuous traits, $\lambda_{1000}$ scales the genomic inflation factor $\lambda$ to a study with 1000 subjects using $\lambda_{1000} = 1 + 1000 * (\lambda - 1)/N$, where N is the total sample size. For binary traits, $\lambda_{1000}$ scales $\lambda$ to a study with 1000 cases and 1000 controls using $\lambda_{1000} = 1 + 1000 * (\lambda - 1) * (\frac{1}{N_{case}} + \frac{1}{N_{control}})$

**Supplementary Figure 6-c: Manhattan plot and QQ plot[1] based on the All of Us multi-ancestry GWAS summary statistics for LDL in six populations (European, African American, Latino, East Asian, South Asian, Middle Eastern), obtained for pooled analysis, meta-analysis and MR-MEGA using mixed-effect modelling** (from top panel to bottom panel). The red, blue, green and purple shaded regions around the diagonal line in the QQ plots indicate the 95% confidence intervals expected under the null hypothesis of no association between genetic markers and the trait of interest, for minor allele frequencies (MAF) within the ranges (0.05, 0.5], (0.01, 0.05], (0.001, 0.005] and (0,0.001] respectively. Under the null hypothesis, the p-value follows a uniform (0,1) distribution. The jth order statistic follows a Beta (j, N-j+1) distribution, where N is the total number of variants given a specific MAF cutoff.

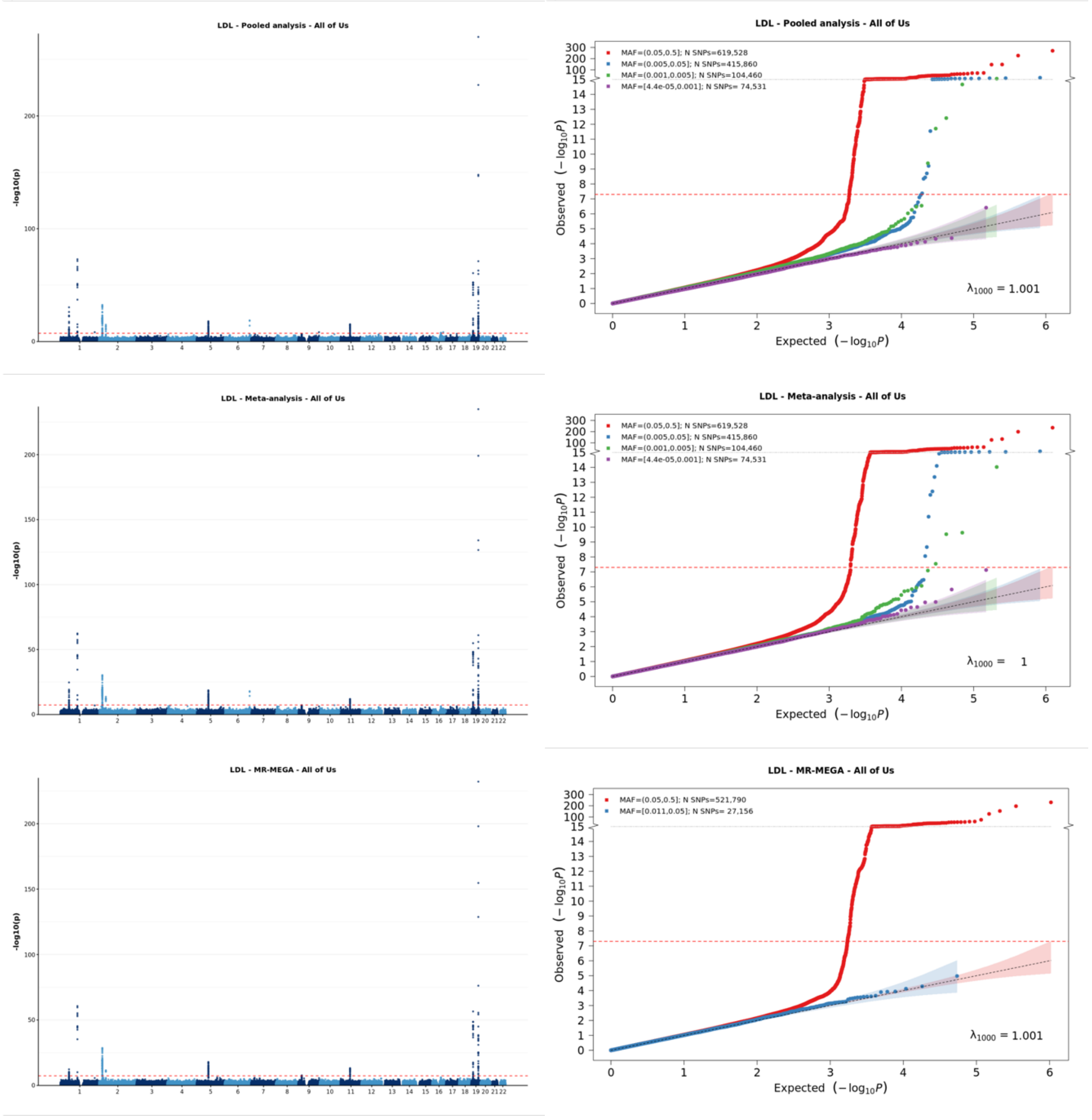


[1] For continuous traits, $\lambda_{1000}$ scales the genomic inflation factor $\lambda$ to a study with 1000 subjects using $\lambda_{1000} = 1 + 1000 * (\lambda - 1)/N$, where N is the total sample size. For binary traits, $\lambda_{1000}$ scales $\lambda$ to a study with 1000 cases and 1000 controls using $\lambda_{1000} = 1 + 1000 * (\lambda - 1) * (\frac{1}{N_{case}} + \frac{1}{N_{control}})$

**Supplementary Figure 6-d: Manhattan plot and QQ plot[1] based on the All of Us multi-ancestry GWAS summary statistics for HDL in six populations (European, African American, Latino, East Asian, South Asian, Middle Eastern), obtained for pooled analysis, meta-analysis and MR-MEGA using mixed-effect modelling** (from top panel to bottom panel). The red, blue, green and purple shaded regions around the diagonal line in the QQ plots indicate the 95% confidence intervals expected under the null hypothesis of no association between genetic markers and the trait of interest, for minor allele frequencies (MAF) within the ranges (0.05, 0.5], (0.01, 0.05], (0.001, 0.005] and (0,0.001] respectively. Under the null hypothesis, the p-value follows a uniform (0,1) distribution. The jth order statistic follows a Beta (j, N-j+1) distribution, where N is the total number of variants given a specific MAF cutoff.

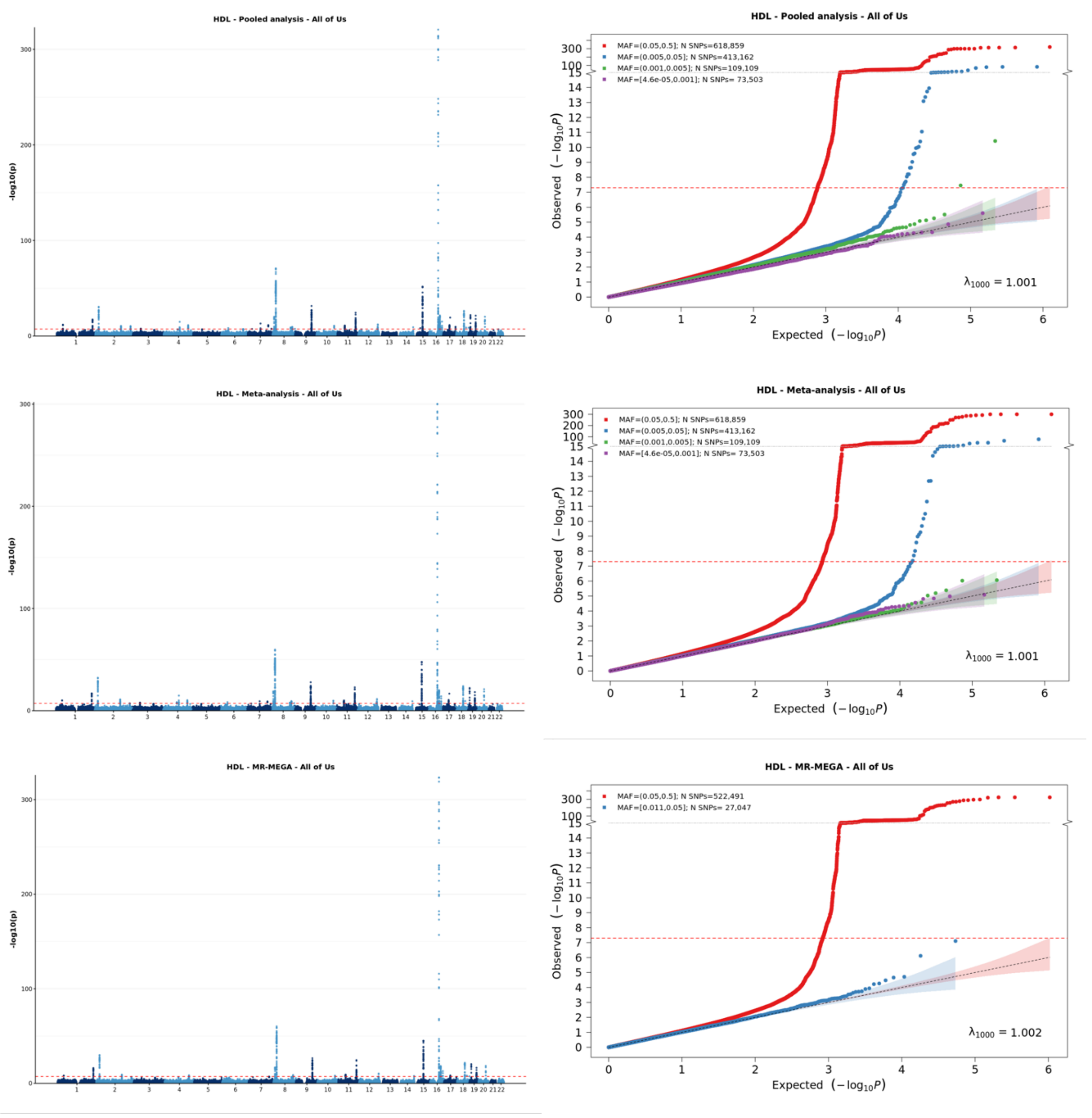


[1] For continuous traits, $\lambda_{1000}$ scales the genomic inflation factor $\lambda$ to a study with 1000 subjects using $\lambda_{1000} = 1 + 1000 * (\lambda - 1)/N$, where N is the total sample size. For binary traits, $\lambda_{1000}$ scales $\lambda$ to a study with 1000 cases and 1000 controls using $\lambda_{1000} = 1 + 1000 * (\lambda - 1) * (\frac{1}{N_{case}} + \frac{1}{N_{control}})$

**Supplementary Figure 6-e: Manhattan plot and QQ plot[1] based on the All of Us multi-ancestry GWAS summary statistics for total cholesterol (TC) in six populations (European, African American, Latino, East Asian, South Asian, Middle Eastern), obtained for pooled analysis, meta-analysis and MR-MEGA using mixed-effect modelling** (from top panel to bottom panel). The red, blue, green and purple shaded regions around the diagonal line in the QQ plots indicate the 95% confidence intervals expected under the null hypothesis of no association between genetic markers and the trait of interest, for minor allele frequencies (MAF) within the ranges (0.05, 0.5], (0.01, 0.05], (0.001, 0.005] and (0,0.001] respectively. Under the null hypothesis, the p-value follows a uniform (0,1) distribution. The jth order statistic follows a Beta (j, N-j+1) distribution, where N is the total number of variants given a specific MAF cutoff.

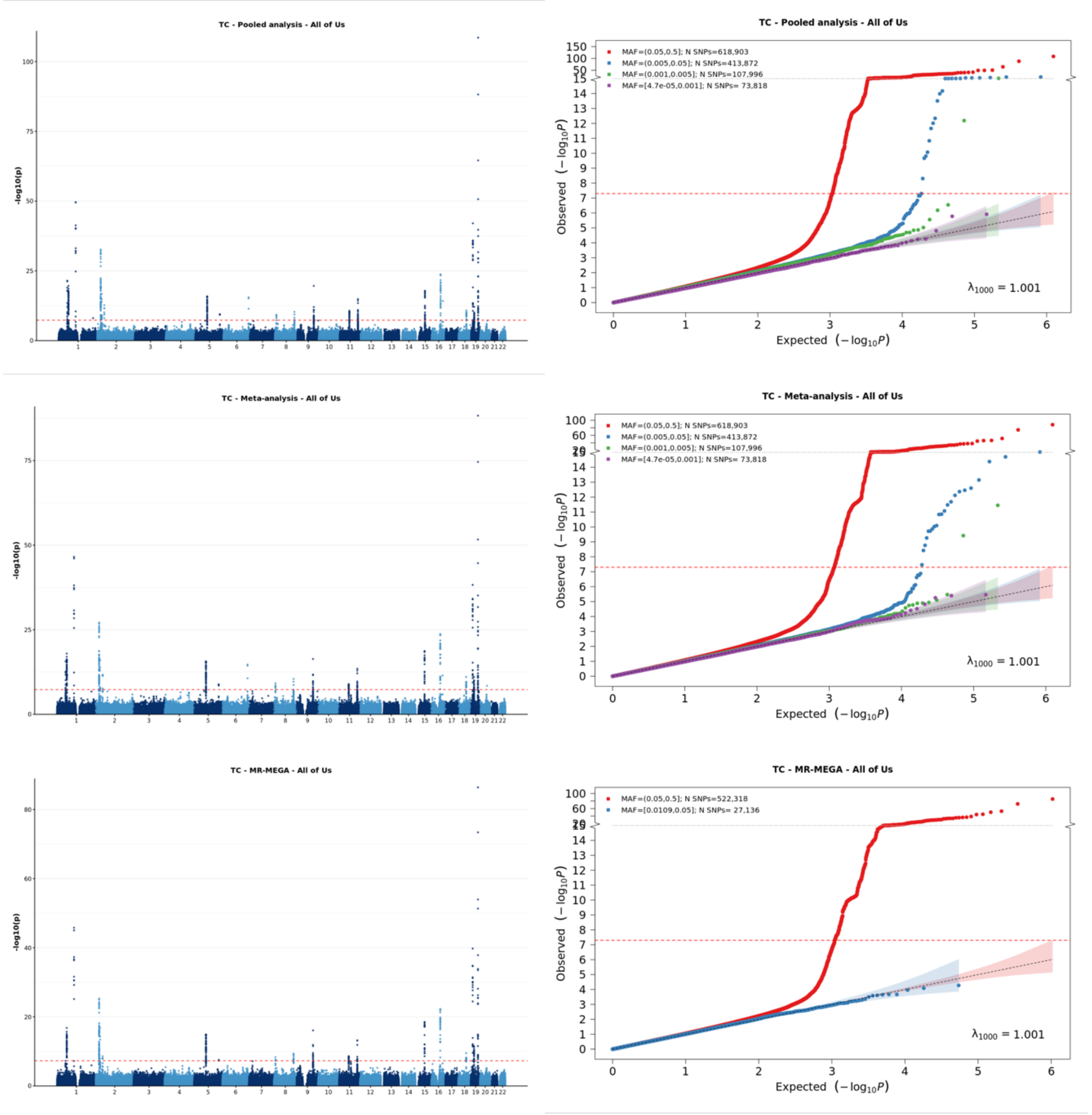


[1] For continuous traits, $\lambda_{1000}$ scales the genomic inflation factor $\lambda$ to a study with 1000 subjects using $\lambda_{1000} = 1 + 1000 * (\lambda - 1)/N$, where N is the total sample size. For binary traits, $\lambda_{1000}$ scales $\lambda$ to a study with 1000 cases and 1000 controls using $\lambda_{1000} = 1 + 1000 * (\lambda - 1) * (\frac{1}{N_{case}} + \frac{1}{N_{control}})$

**Supplementary Figure 6-f: Manhattan plot and QQ plot[1] based on the All of Us multi-ancestry GWAS summary statistics for calcium in six populations (European, African American, Latino, East Asian, South Asian, Middle Eastern), obtained for pooled analysis, meta-analysis and MR-MEGA using mixed-effect modelling** (from top panel to bottom panel). The red, blue, green and purple shaded regions around the diagonal line in the QQ plots indicate the 95% confidence intervals expected under the null hypothesis of no association between genetic markers and the trait of interest, for minor allele frequencies (MAF) within the ranges (0.05, 0.5], (0.01, 0.05], (0.001, 0.005] and (0,0.001] respectively. Under the null hypothesis, the p-value follows a uniform (0,1) distribution. The jth order statistic follows a Beta (j, N-j+1) distribution, where N is the total number of variants given a specific MAF cutoff.

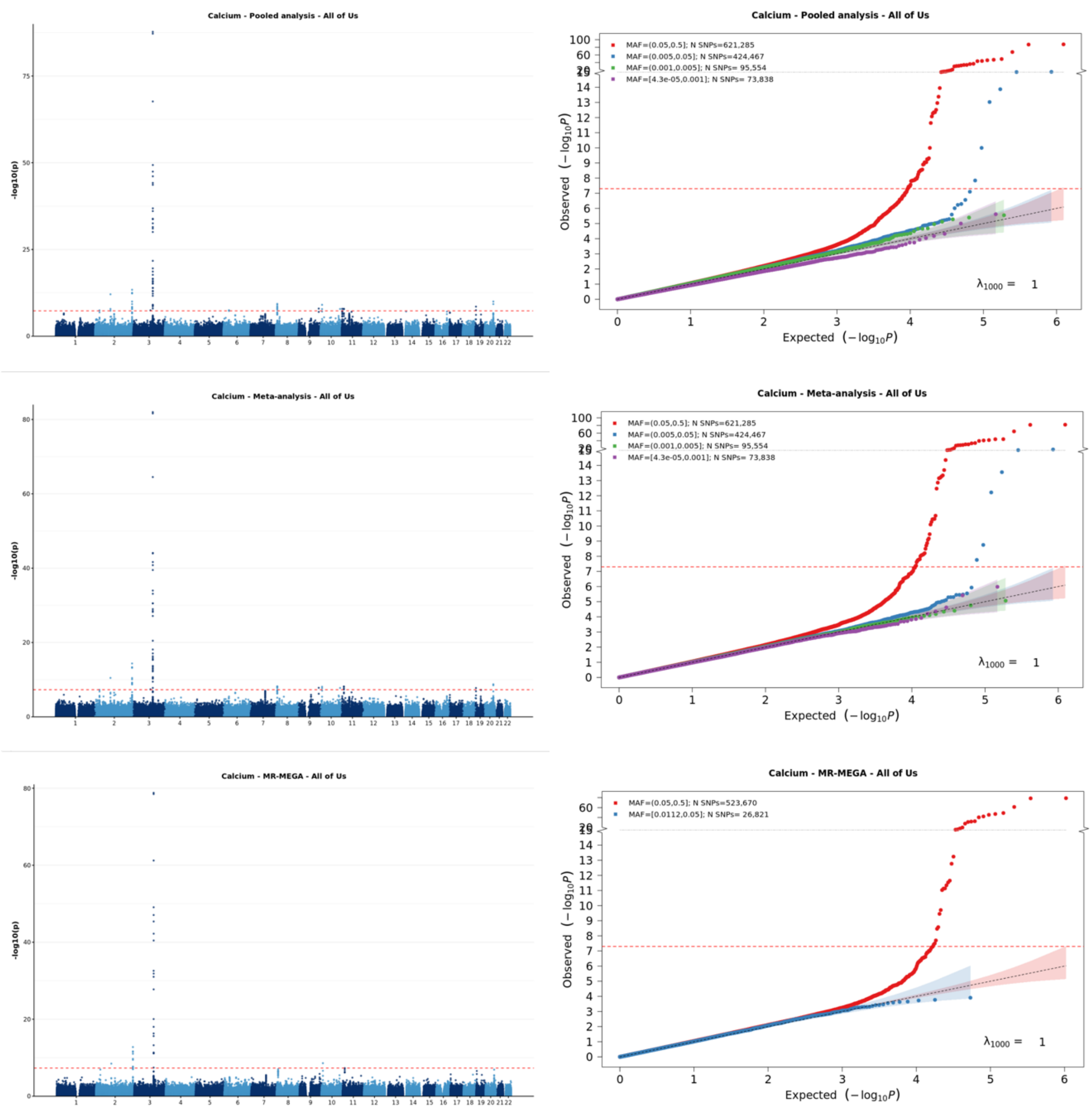


[1] For continuous traits, $\lambda_{1000}$ scales the genomic inflation factor $\lambda$ to a study with 1000 subjects using $\lambda_{1000} = 1 + 1000 * (\lambda - 1)/N$, where N is the total sample size. For binary traits, $\lambda_{1000}$ scales $\lambda$ to a study with 1000 cases and 1000 controls using $\lambda_{1000} = 1 + 1000 * (\lambda - 1) * (\frac{1}{N_{case}} + \frac{1}{N_{control}})$

**Supplementary Figure 6-g: Manhattan plot and QQ plot[1] based on the All of Us multi-ancestry GWAS summary statistics for creatinine in six populations (European, African American, Latino, East Asian, South Asian, Middle Eastern), obtained for pooled analysis, meta-analysis and MR-MEGA using mixed-effect modelling** (from top panel to bottom panel). The red, blue, green and purple shaded regions around the diagonal line in the QQ plots indicate the 95% confidence intervals expected under the null hypothesis of no association between genetic markers and the trait of interest, for minor allele frequencies (MAF) within the ranges (0.05, 0.5], (0.01, 0.05], (0.001, 0.005] and (0,0.001] respectively. Under the null hypothesis, the p-value follows a uniform (0,1) distribution. The jth order statistic follows a Beta (j, N-j+1) distribution, where N is the total number of variants given a specific MAF cutoff.

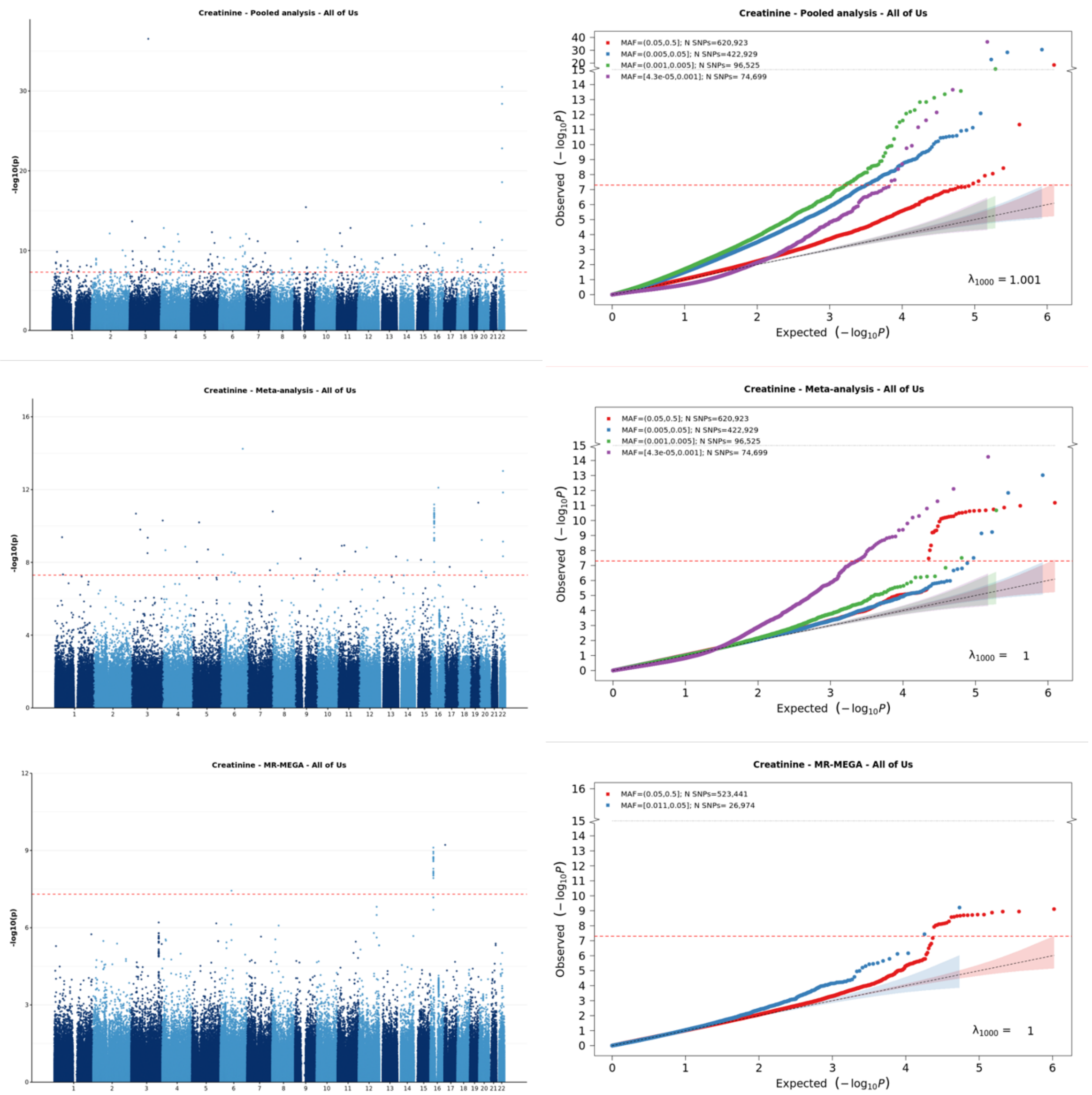


[1] For continuous traits, $\lambda_{1000}$ scales the genomic inflation factor $\lambda$ to a study with 1000 subjects using $\lambda_{1000} = 1 + 1000 * (\lambda - 1)/N$, where N is the total sample size. For binary traits, $\lambda_{1000}$ scales $\lambda$ to a study with 1000 cases and 1000 controls using $\lambda_{1000} = 1 + 1000 * (\lambda - 1) * (\frac{1}{N_{case}} + \frac{1}{N_{control}})$

**Supplementary Figure 6-h: Manhattan plot and QQ plot[1] based on the All of Us multi-ancestry GWAS summary statistics for EGFR in six populations (European, African American, Latino, East Asian, South Asian, Middle Eastern), obtained for pooled analysis, meta-analysis and MR-MEGA using mixed-effect modelling** (from top panel to bottom panel). The red, blue, green and purple shaded regions around the diagonal line in the QQ plots indicate the 95% confidence intervals expected under the null hypothesis of no association between genetic markers and the trait of interest, for minor allele frequencies (MAF) within the ranges (0.05, 0.5], (0.01, 0.05], (0.001, 0.005] and (0,0.001] respectively. Under the null hypothesis, the p-value follows a uniform (0,1) distribution. The jth order statistic follows a Beta (j, N-j+1) distribution, where N is the total number of variants given a specific MAF cutoff.

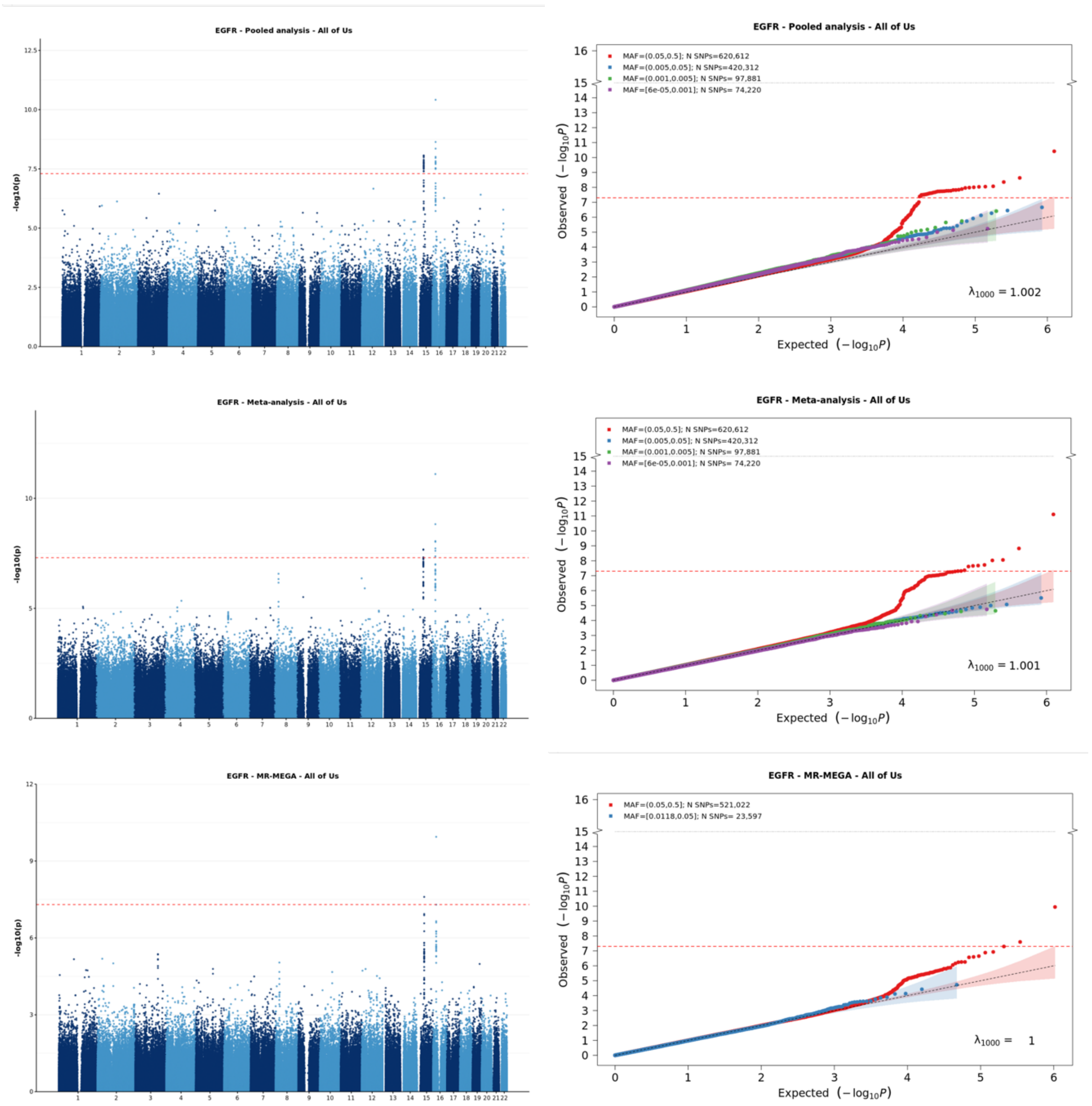


[1] For continuous traits, $\lambda_{1000}$ scales the genomic inflation factor $\lambda$ to a study with 1000 subjects using $\lambda_{1000} = 1 + 1000 * (\lambda - 1)/N$, where N is the total sample size. For binary traits, $\lambda_{1000}$ scales $\lambda$ to a study with 1000 cases and 1000 controls using $\lambda_{1000} = 1 + 1000 * (\lambda - 1) * (\frac{1}{N_{case}} + \frac{1}{N_{control}})$

**Supplementary Figure 6-i: Manhattan plot and QQ plot[1] based on the All of Us multi-ancestry GWAS summary statistics for asthma in six populations (European, African American, Latino, East Asian, South Asian, Middle Eastern), obtained for pooled analysis, meta-analysis and MR-MEGA using mixed-effect modelling** (from top panel to bottom panel). The red, blue, green and purple shaded regions around the diagonal line in the QQ plots indicate the 95% confidence intervals expected under the null hypothesis of no association between genetic markers and the trait of interest, for minor allele frequencies (MAF) within the ranges (0.05, 0.5], (0.01, 0.05], (0.001, 0.005] and (0,0.001] respectively. Under the null hypothesis, the p-value follows a uniform (0,1) distribution. The jth order statistic follows a Beta (j, N-j+1) distribution, where N is the total number of variants given a specific MAF cutoff.

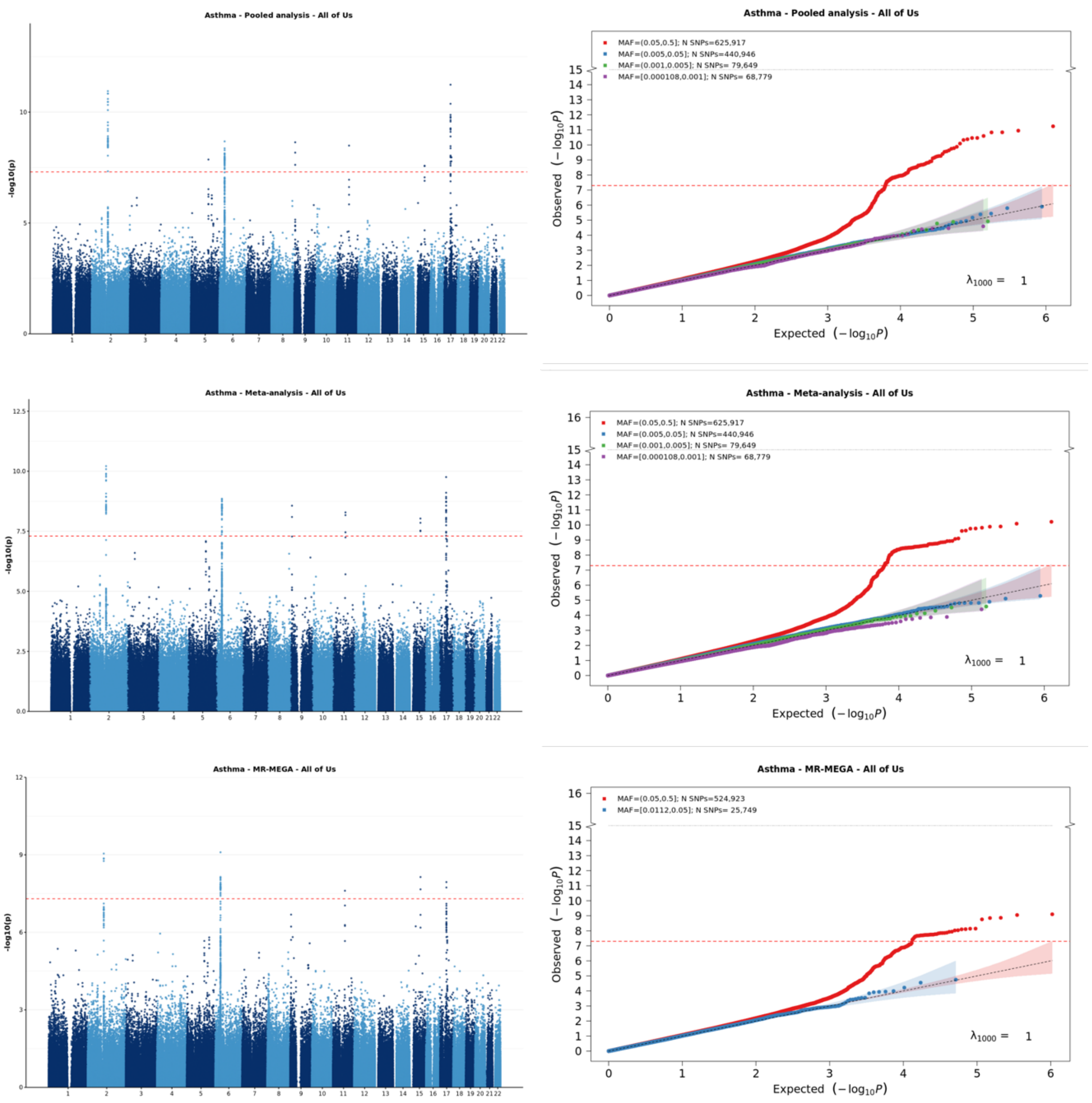


[1] For continuous traits, $\lambda_{1000}$ scales the genomic inflation factor $\lambda$ to a study with 1000 subjects using $\lambda_{1000} = 1 + 1000 * (\lambda - 1)/N$, where N is the total sample size. For binary traits, $\lambda_{1000}$ scales $\lambda$ to a study with 1000 cases and 1000 controls using $\lambda_{1000} = 1 + 1000 * (\lambda - 1) * (\frac{1}{N_{case}} + \frac{1}{N_{control}})$

**Supplementary Figure 6-j: Manhattan plot and QQ plot[1] based on the All of Us multi-ancestry GWAS summary statistics for coronary artery disease (CAD) in six populations (European, African American, Latino, East Asian, South Asian, Middle Eastern), obtained for pooled analysis, meta-analysis and MR-MEGA using mixed-effect modelling** (from top panel to bottom panel). The red, blue, green and purple shaded regions around the diagonal line in the QQ plots indicate the 95% confidence intervals expected under the null hypothesis of no association between genetic markers and the trait of interest, for minor allele frequencies (MAF) within the ranges (0.05, 0.5], (0.01, 0.05], (0.001, 0.005] and (0,0.001] respectively. Under the null hypothesis, the p-value follows a uniform (0,1) distribution. The jth order statistic follows a Beta (j, N-j+1) distribution, where N is the total number of variants given a specific MAF cutoff.

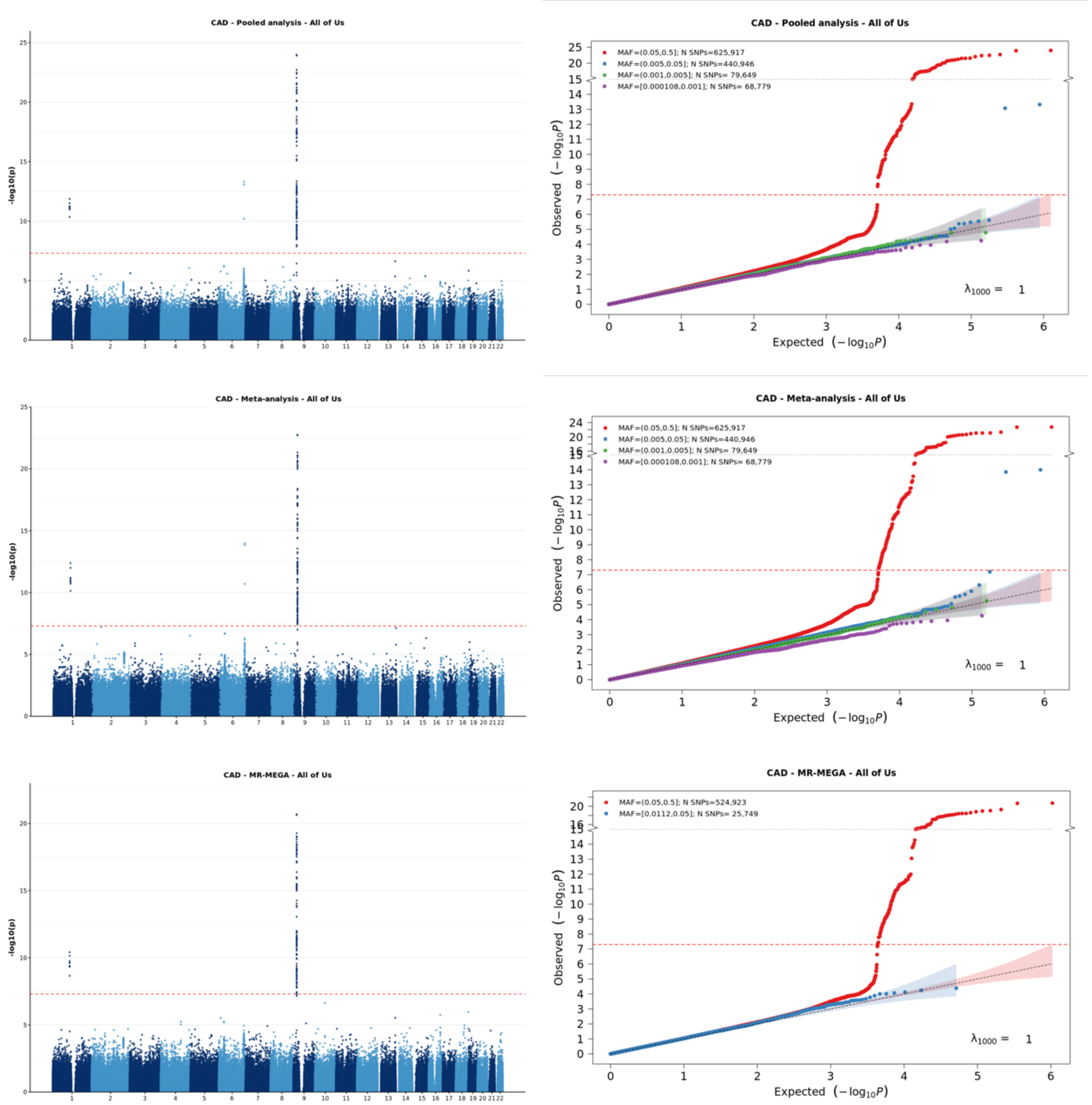


[1] For continuous traits, $\lambda_{1000}$ scales the genomic inflation factor $\lambda$ to a study with 1000 subjects using $\lambda_{1000} = 1 + 1000 * (\lambda - 1)/N$, where N is the total sample size. For binary traits, $\lambda_{1000}$ scales $\lambda$ to a study with 1000 cases and 1000 controls using $\lambda_{1000} = 1 + 1000 * (\lambda - 1) * (\frac{1}{N_{case}} + \frac{1}{N_{control}})$

**Supplementary Figure 6-k: Manhattan plot and QQ plot[1] based on the All of Us multi-ancestry GWAS summary statistics for type 2 diabetes in six populations (European, African American, Latino, East Asian, South Asian, Middle Eastern), obtained for pooled analysis, meta-analysis and MR-MEGA using mixed-effect modelling** (from top panel to bottom panel). The red, blue, green and purple shaded regions around the diagonal line in the QQ plots indicate the 95% confidence intervals expected under the null hypothesis of no association between genetic markers and the trait of interest, for minor allele frequencies (MAF) within the ranges (0.05, 0.5], (0.01, 0.05], (0.001, 0.005] and (0,0.001] respectively. Under the null hypothesis, the p-value follows a uniform (0,1) distribution. The jth order statistic follows a Beta (j, N-j+1) distribution, where N is the total number of variants given a specific MAF cutoff.

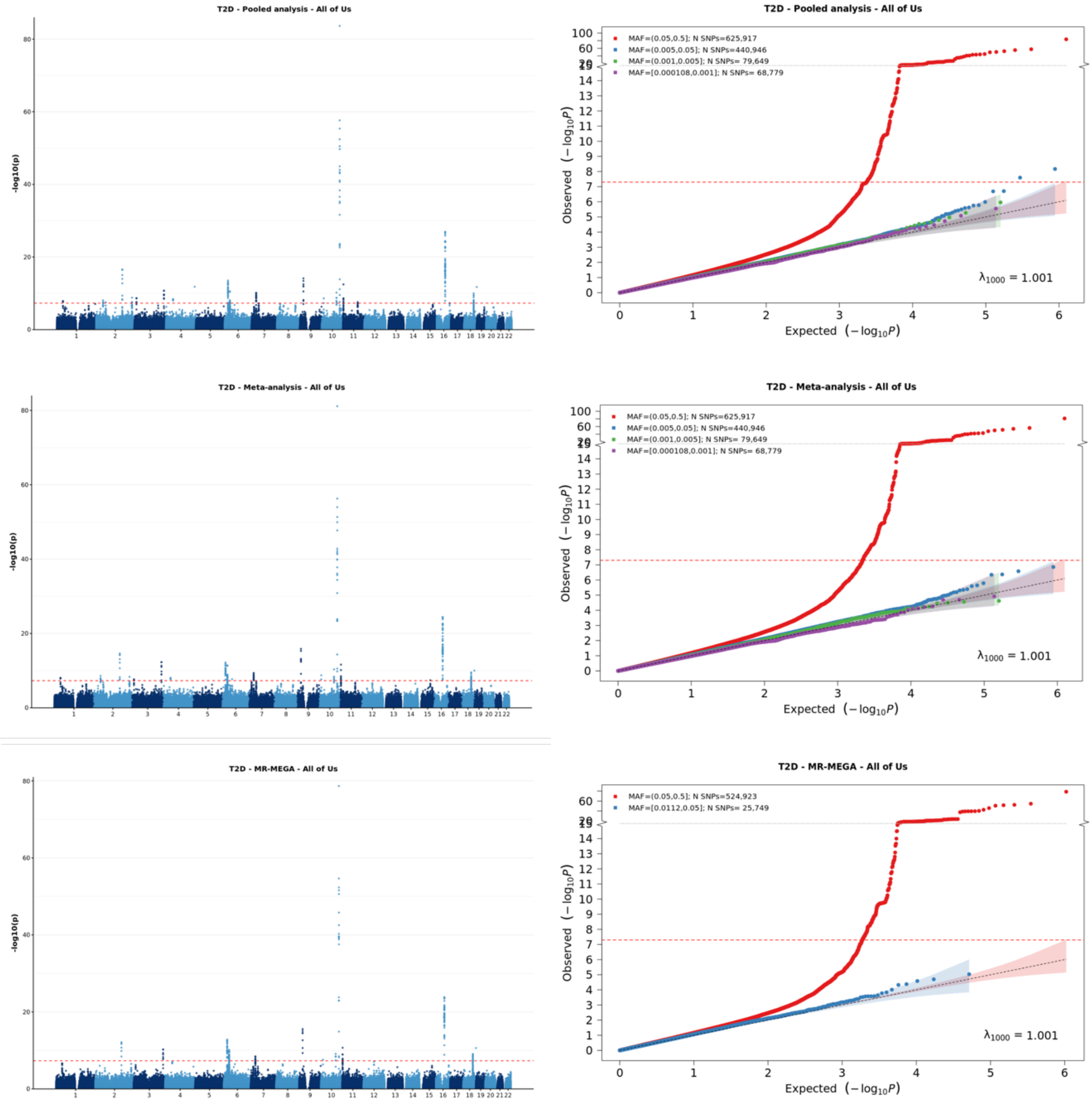


[1] For continuous traits, $\lambda_{1000}$ scales the genomic inflation factor $\lambda$ to a study with 1000 subjects using $\lambda_{1000} = 1 + 1000 * (\lambda - 1)/N$, where N is the total sample size. For binary traits, $\lambda_{1000}$ scales $\lambda$ to a study with 1000 cases and 1000 controls using $\lambda_{1000} = 1 + 1000 * (\lambda - 1) * (\frac{1}{N_{case}} + \frac{1}{N_{control}})$

**Supplementary Figure 6-I: Manhattan plot and QQ plot[1] based on the All of Us multi-ancestry GWAS summary statistics for breast cancer in five populations (European, African American, Latino, East Asian, South Asian), obtained for pooled analysis, meta-analysis and MR-MEGA using mixed-effect modelling** (from top panel to bottom panel). The red, blue, green and purple shaded regions around the diagonal line in the QQ plots indicate the 95% confidence intervals expected under the null hypothesis of no association between genetic markers and the trait of interest, for minor allele frequencies (MAF) within the ranges (0.05, 0.5], (0.01, 0.05], (0.001, 0.005] and (0,0.001] respectively. Under the null hypothesis, the p-value follows a uniform (0,1) distribution. The jth order statistic follows a Beta (j, N-j+1) distribution, where N is the total number of variants given a specific MAF cutoff.

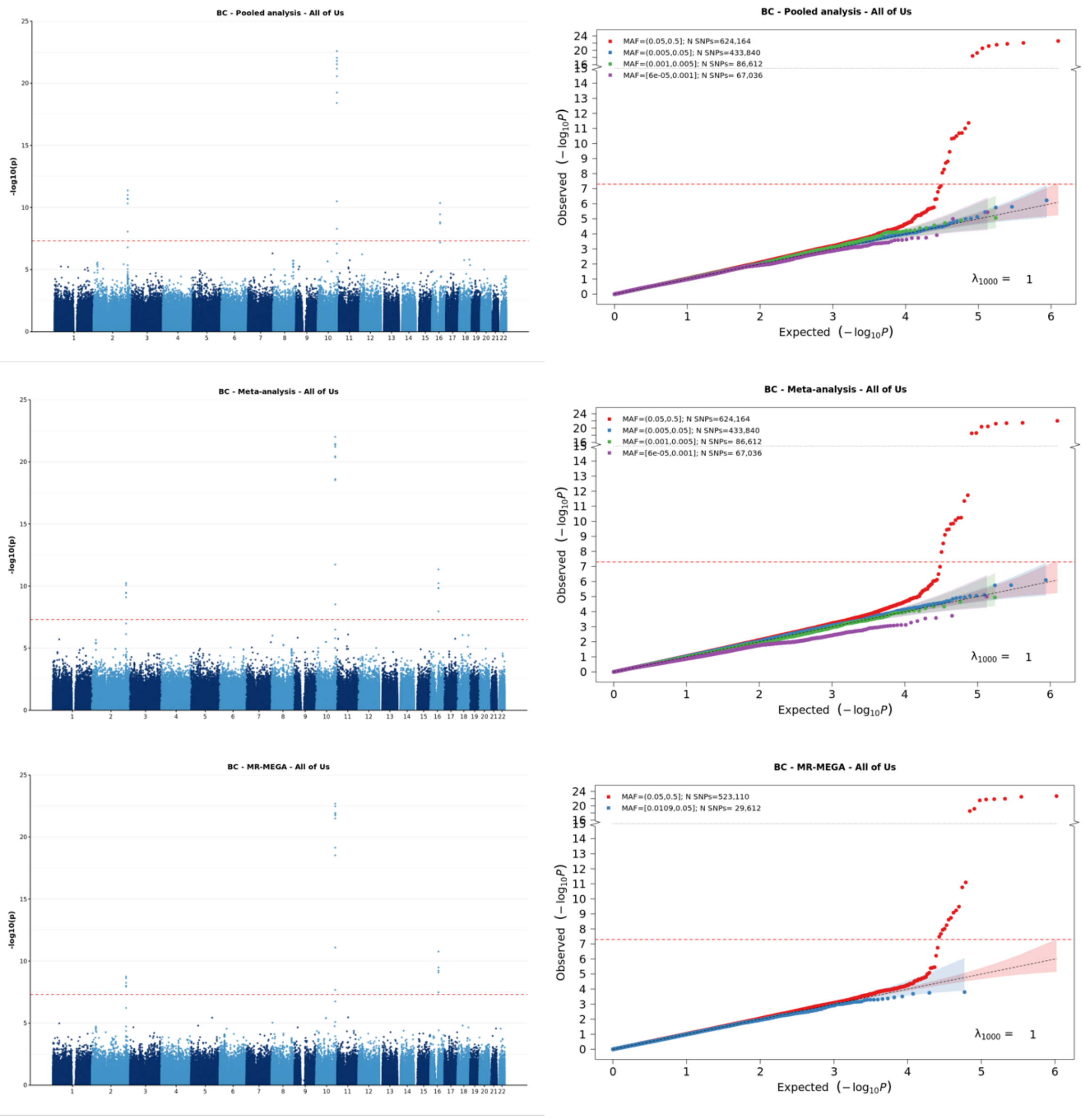


[1] For continuous traits, $\lambda_{1000}$ scales the genomic inflation factor $\lambda$ to a study with 1000 subjects using $\lambda_{1000} = 1 + 1000 * (\lambda - 1)/N$, where N is the total sample size. For binary traits, $\lambda_{1000}$ scales $\lambda$ to a study with 1000 cases and 1000 controls using $\lambda_{1000} = 1 + 1000 * (\lambda - 1) * (\frac{1}{N_{case}} + \frac{1}{N_{control}})$

**Supplementary Figure 6-m: Manhattan plot and QQ plot[1] based on the All of Us multi-ancestry GWAS summary statistics for prostate cancer in five populations (European, African American, Latino, East Asian, South Asian), obtained for pooled analysis, meta-analysis and MR-MEGA using mixed-effect modelling** (from top panel to bottom panel). The red, blue, green and purple shaded regions around the diagonal line in the QQ plots indicate the 95% confidence intervals expected under the null hypothesis of no association between genetic markers and the trait of interest, for minor allele frequencies (MAF) within the ranges (0.05, 0.5], (0.01, 0.05], (0.001, 0.005] and (0,0.001] respectively. Under the null hypothesis, the p-value follows a uniform (0,1) distribution. The jth order statistic follows a Beta (j, N-j+1) distribution, where N is the total number of variants given a specific MAF cutoff.

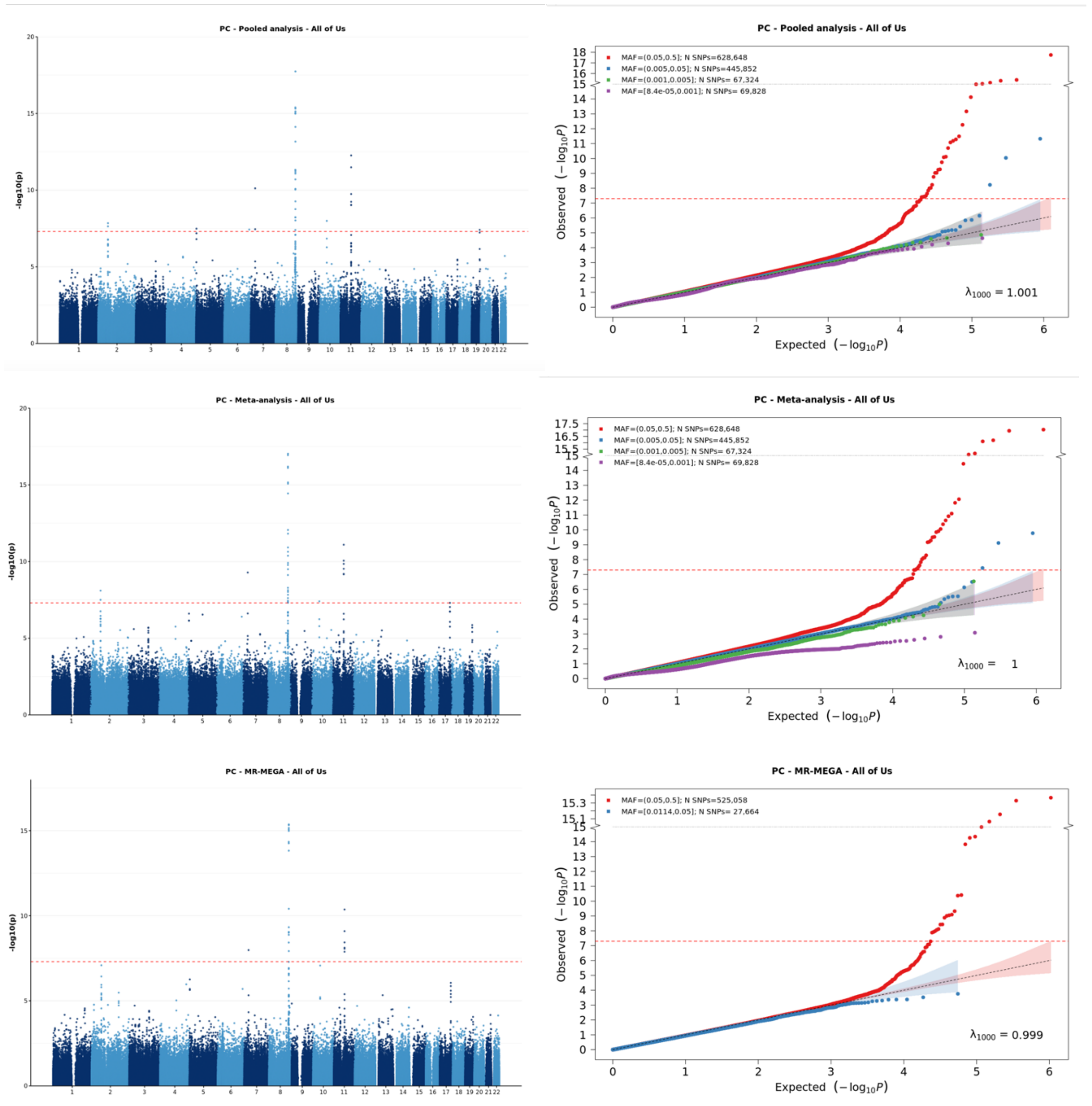


[1] For continuous traits, $\lambda_{1000}$ scales the genomic inflation factor $\lambda$ to a study with 1000 subjects using $\lambda_{1000} = 1 + 1000 * (\lambda - 1)/N$, where N is the total sample size. For binary traits, $\lambda_{1000}$ scales $\lambda$ to a study with 1000 cases and 1000 controls using $\lambda_{1000} = 1 + 1000 * (\lambda - 1) * (\frac{1}{N_{case}} + \frac{1}{N_{control}})$

**Supplementary Figure 7-a: Manhattan plot and QQ plot[1] based on the UK Biobank multi-ancestry GWAS summary statistics for height in five populations (European, African American, Latino, East Asian and South Asian), obtained for pooled analysis, meta-analysis and MR-MEGA using mixed-effect modelling** (from top panel to bottom panel). The red, blue, green and purple shaded regions around the diagonal line in the QQ plots indicate the 95% confidence intervals expected under the null hypothesis of no association between genetic markers and the trait of interest, for minor allele frequencies (MAF) within the ranges (0.05, 0.5], (0.01, 0.05], (0.001, 0.005] and (0,0.001] respectively. Under the null hypothesis, the p-value follows a uniform (0,1) distribution. The jth order statistic follows a Beta (j, N-j+1) distribution, where N is the total number of variants given a specific MAF cutoff. HERE

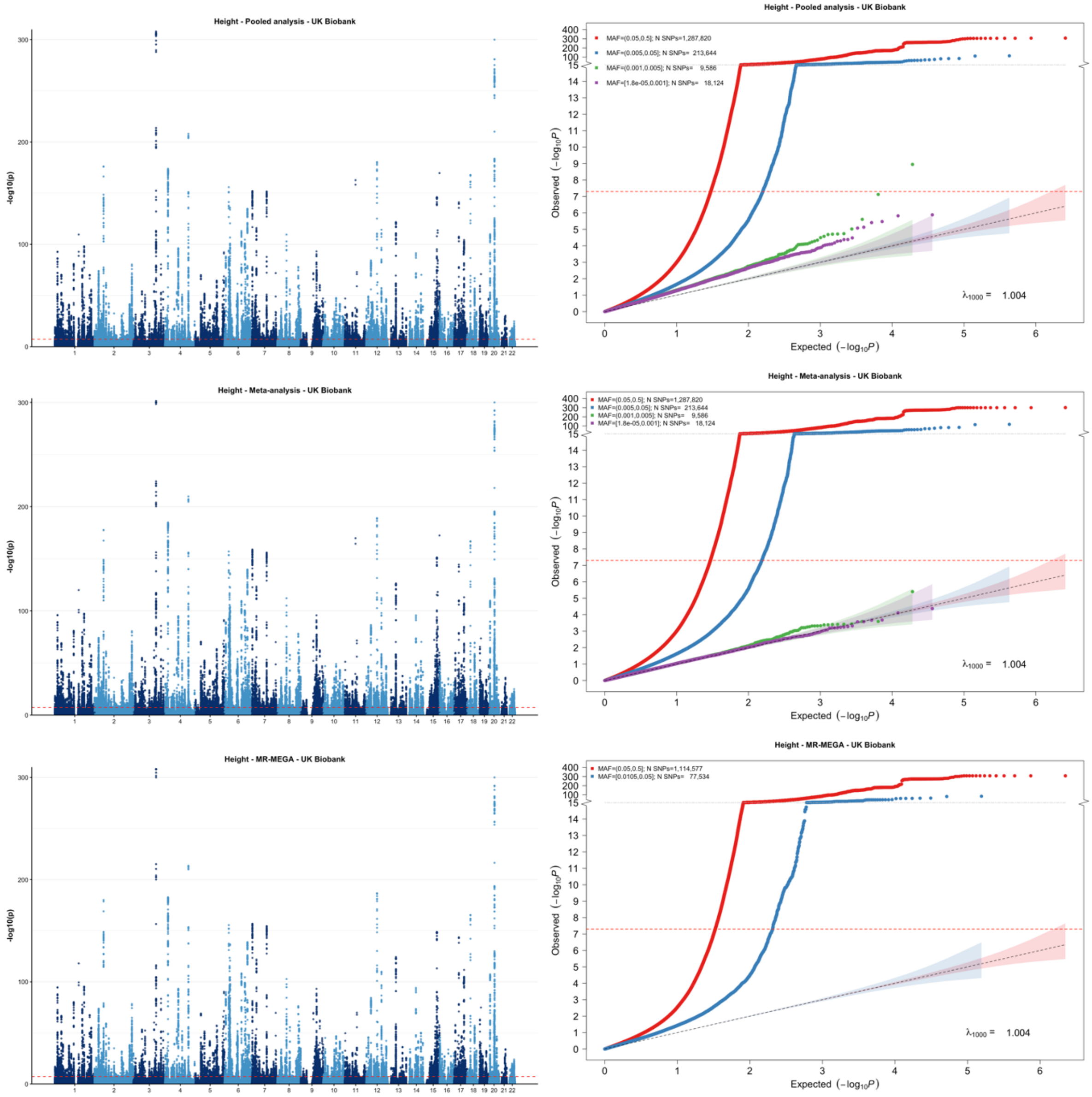


[1] For continuous traits, $\lambda_{1000}$ scales the genomic inflation factor $\lambda$ to a study with 1000 subjects using $\lambda_{1000} = 1 + 1000 * (\lambda - 1)/N$, where N is the total sample size. For binary traits, $\lambda_{1000}$ scales $\lambda$ to a study with 1000 cases and 1000 controls using $\lambda_{1000} = 1 + 1000 * (\lambda - 1) * (\frac{1}{N_{case}} + \frac{1}{N_{control}})$

**Supplementary Figure 7-b: Manhattan plot and QQ plot[1] based on the UK Biobank multi-ancestry GWAS summary statistics for waist circumference in five populations (European, African American, Latino, East Asian and South Asian), obtained for pooled analysis, meta-analysis and MR-MEGA using mixed-effect modelling** (from top panel to bottom panel). The red, blue, green and purple shaded regions around the diagonal line in the QQ plots indicate the 95% confidence intervals expected under the null hypothesis of no association between genetic markers and the trait of interest, for minor allele frequencies (MAF) within the ranges (0.05, 0.5], (0.01, 0.05], (0.001, 0.005] and (0,0.001] respectively. Under the null hypothesis, the p-value follows a uniform (0,1) distribution. The jth order statistic follows a Beta (j, N-j+1) distribution, where N is the total number of variants given a specific MAF cutoff.

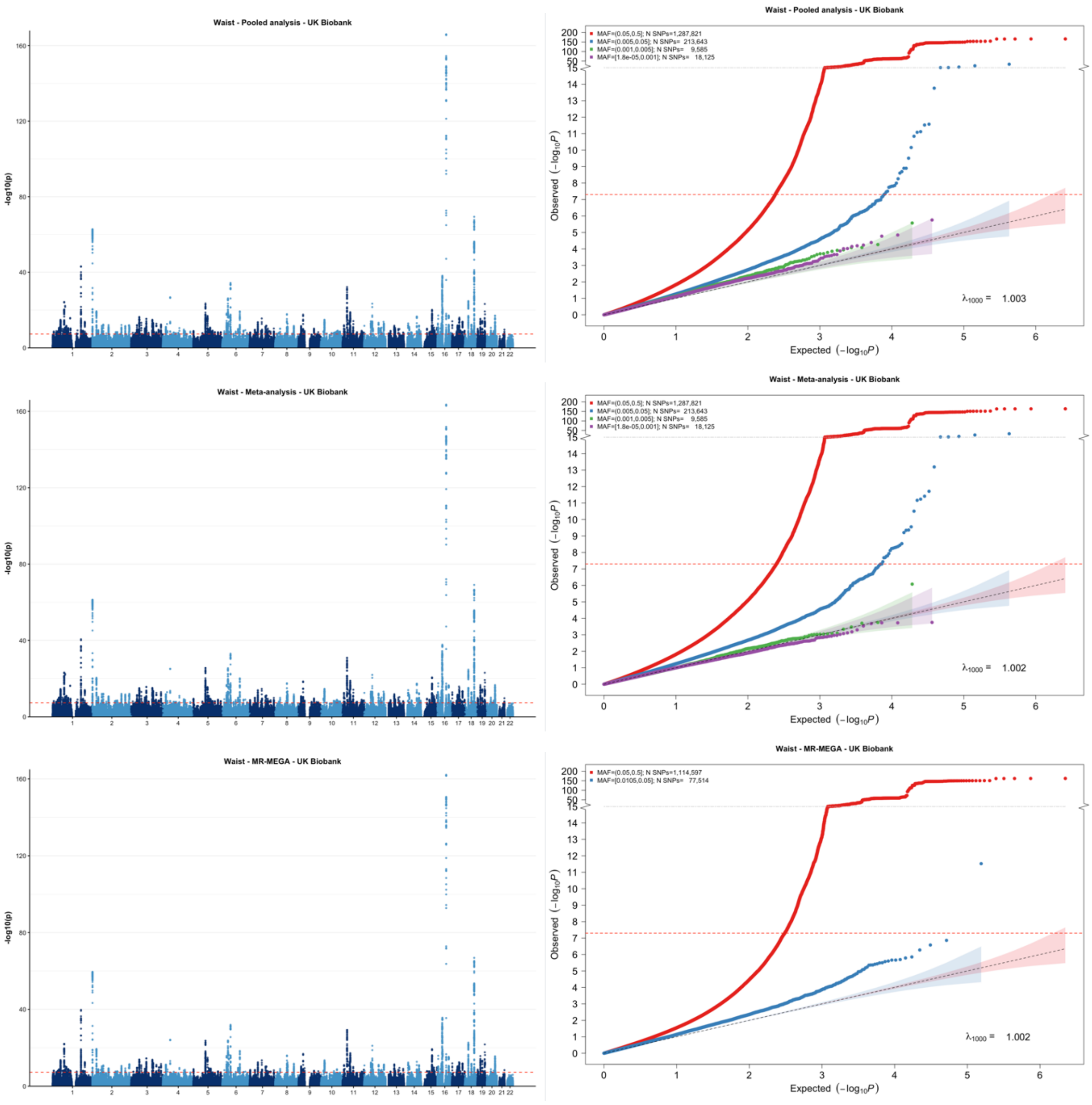


[1] For continuous traits, $\lambda_{1000}$ scales the genomic inflation factor $\lambda$ to a study with 1000 subjects using $\lambda_{1000} = 1 + 1000 * (\lambda - 1)/N$, where N is the total sample size. For binary traits, $\lambda_{1000}$ scales $\lambda$ to a study with 1000 cases and 1000 controls using $\lambda_{1000} = 1 + 1000 * (\lambda - 1) * (\frac{1}{N_{case}} + \frac{1}{N_{control}})$

**Supplementary Figure 7-c: Manhattan plot and QQ plot[1] based on the UK Biobank multi-ancestry GWAS summary statistics for LDL in five populations (European, African American, Latino, East Asian and South Asian), obtained for pooled analysis, meta-analysis and MR-MEGA using mixed-effect modelling** (from top panel to bottom panel). The red, blue, green and purple shaded regions around the diagonal line in the QQ plots indicate the 95% confidence intervals expected under the null hypothesis of no association between genetic markers and the trait of interest, for minor allele frequencies (MAF) within the ranges (0.05, 0.5], (0.01, 0.05], (0.001, 0.005] and (0,0.001] respectively. Under the null hypothesis, the p-value follows a uniform (0,1) distribution. The jth order statistic follows a Beta (j, N-j+1) distribution, where N is the total number of variants given a specific MAF cutoff.

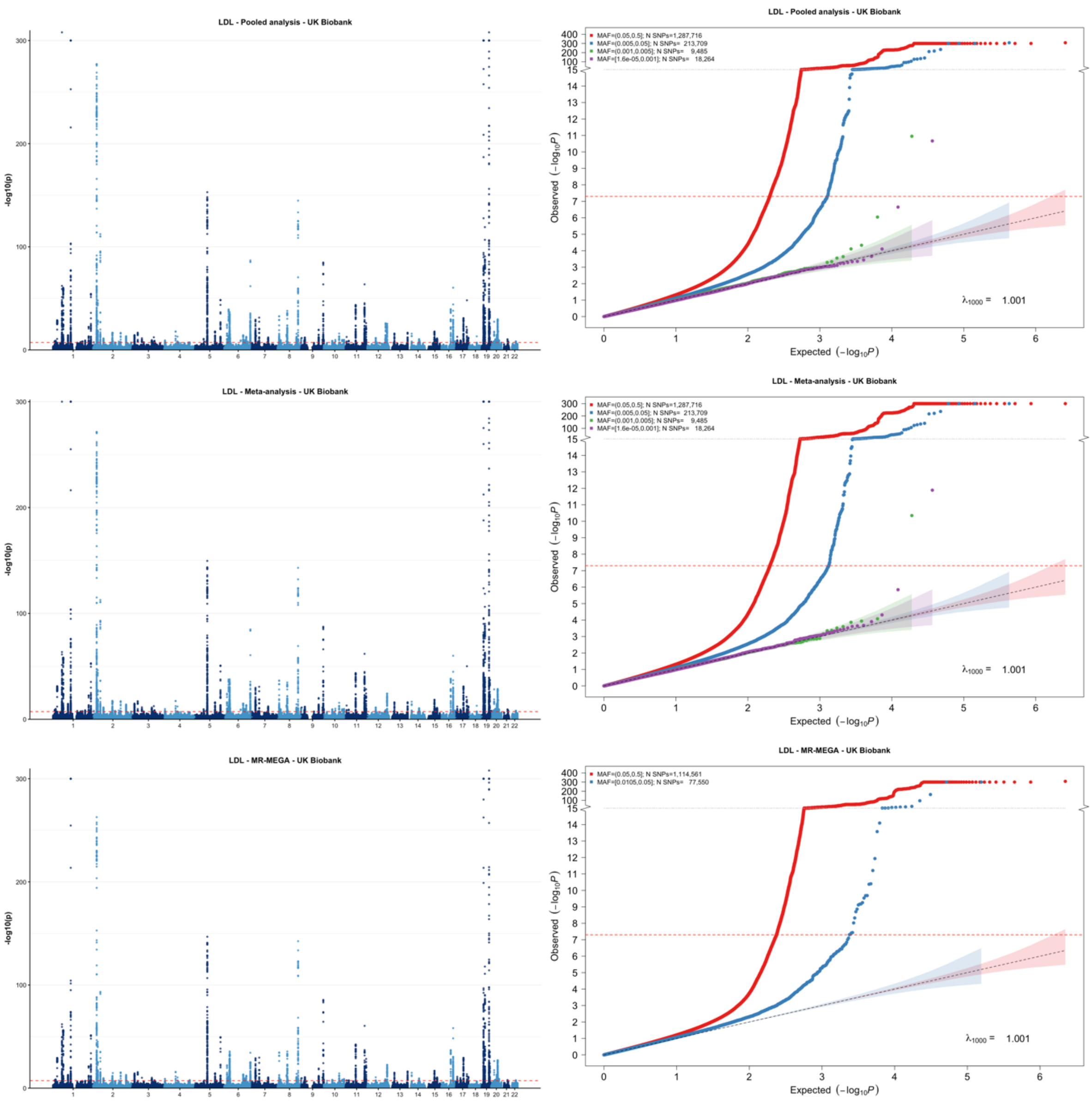


[1] For continuous traits, $\lambda_{1000}$ scales the genomic inflation factor $\lambda$ to a study with 1000 subjects using $\lambda_{1000} = 1 + 1000 * (\lambda - 1)/N$, where N is the total sample size. For binary traits, $\lambda_{1000}$ scales $\lambda$ to a study with 1000 cases and 1000 controls using $\lambda_{1000} = 1 + 1000 * (\lambda - 1) * (\frac{1}{N_{case}} + \frac{1}{N_{control}})$

**Supplementary Figure 7-d: Manhattan plot and QQ plot[1] based on the UK Biobank multi-ancestry GWAS summary statistics for HDL in six populations (European, African American, Latino, East Asian, South Asian, Middle Eastern), obtained for pooled analysis, meta-analysis and MR-MEGA using mixed-effect modelling** (from top panel to bottom panel). The red, blue, green and purple shaded regions around the diagonal line in the QQ plots indicate the 95% confidence intervals expected under the null hypothesis of no association between genetic markers and the trait of interest, for minor allele frequencies (MAF) within the ranges (0.05, 0.5], (0.01, 0.05], (0.001, 0.005] and (0,0.001] respectively. Under the null hypothesis, the p-value follows a uniform (0,1) distribution. The jth order statistic follows a Beta (j, N-j+1) distribution, where N is the total number of variants given a specific MAF cutoff.

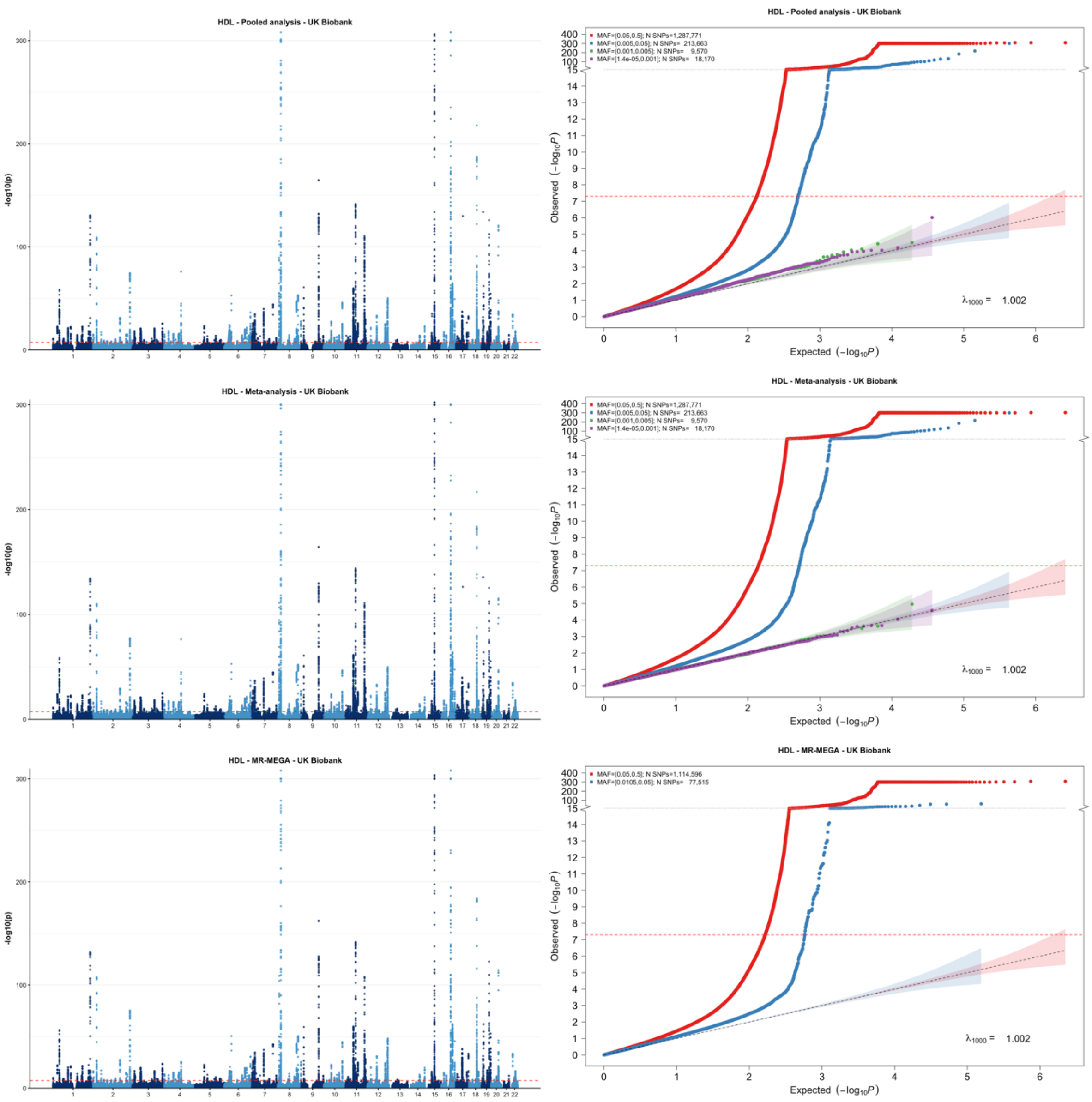


[1] For continuous traits, $\lambda_{1000}$ scales the genomic inflation factor $\lambda$ to a study with 1000 subjects using $\lambda_{1000} = 1 + 1000 * (\lambda - 1)/N$, where N is the total sample size. For binary traits, $\lambda_{1000}$ scales $\lambda$ to a study with 1000 cases and 1000 controls using $\lambda_{1000} = 1 + 1000 * (\lambda - 1) * (\frac{1}{N_{case}} + \frac{1}{N_{control}})$

**Supplementary Figure 7-e: Manhattan plot and QQ plot[1] based on the UK Biobank multi-ancestry GWAS summary statistics for total cholesterol (TC) in five populations (European, African American, Latino, East Asian and South Asian), obtained for pooled analysis, meta-analysis and MR-MEGA using mixed-effect modelling** (from top panel to bottom panel). The red, blue, green and purple shaded regions around the diagonal line in the QQ plots indicate the 95% confidence intervals expected under the null hypothesis of no association between genetic markers and the trait of interest, for minor allele frequencies (MAF) within the ranges (0.05, 0.5], (0.01, 0.05], (0.001, 0.005] and (0,0.001] respectively. Under the null hypothesis, the p-value follows a uniform (0,1) distribution. The jth order statistic follows a Beta (j, N-j+1) distribution, where N is the total number of variants given a specific MAF cutoff.

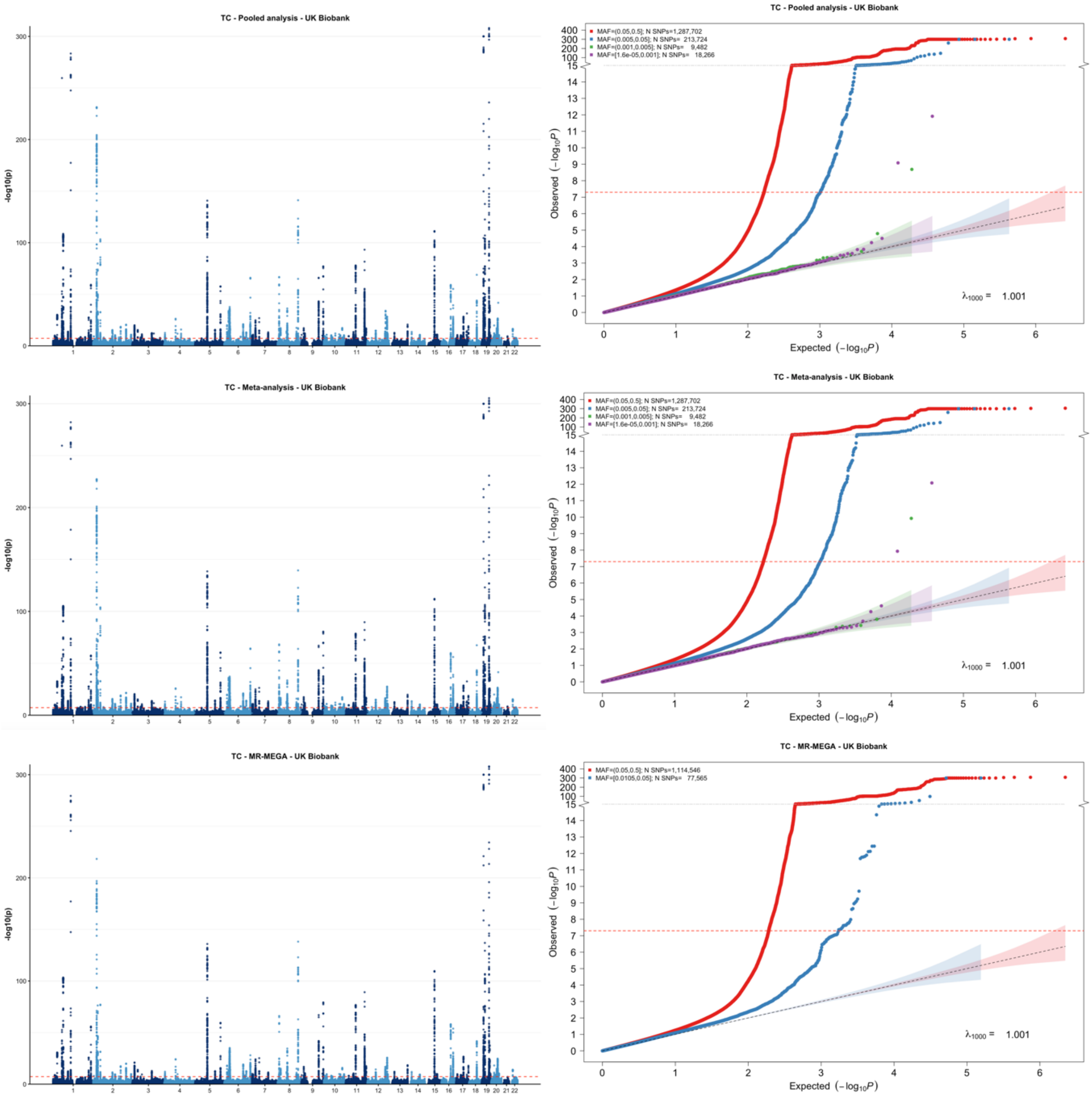


[1] For continuous traits, $\lambda_{1000}$ scales the genomic inflation factor $\lambda$ to a study with 1000 subjects using $\lambda_{1000} = 1 + 1000 * (\lambda - 1)/N$, where N is the total sample size. For binary traits, $\lambda_{1000}$ scales $\lambda$ to a study with 1000 cases and 1000 controls using $\lambda_{1000} = 1 + 1000 * (\lambda - 1) * (\frac{1}{N_{case}} + \frac{1}{N_{control}})$

**Supplementary Figure 7-f: Manhattan plot and QQ plot[1] based on the UK Biobank multi-ancestry GWAS summary statistics for calcium in five populations (European, African American, Latino, East Asian and South Asian), obtained for pooled analysis, meta-analysis and MR-MEGA using mixed-effect modelling** (from top panel to bottom panel). The red, blue, green and purple shaded regions around the diagonal line in the QQ plots indicate the 95% confidence intervals expected under the null hypothesis of no association between genetic markers and the trait of interest, for minor allele frequencies (MAF) within the ranges (0.05, 0.5], (0.01, 0.05], (0.001, 0.005] and (0,0.001] respectively. Under the null hypothesis, the p-value follows a uniform (0,1) distribution. The jth order statistic follows a Beta (j, N-j+1) distribution, where N is the total number of variants given a specific MAF cutoff.

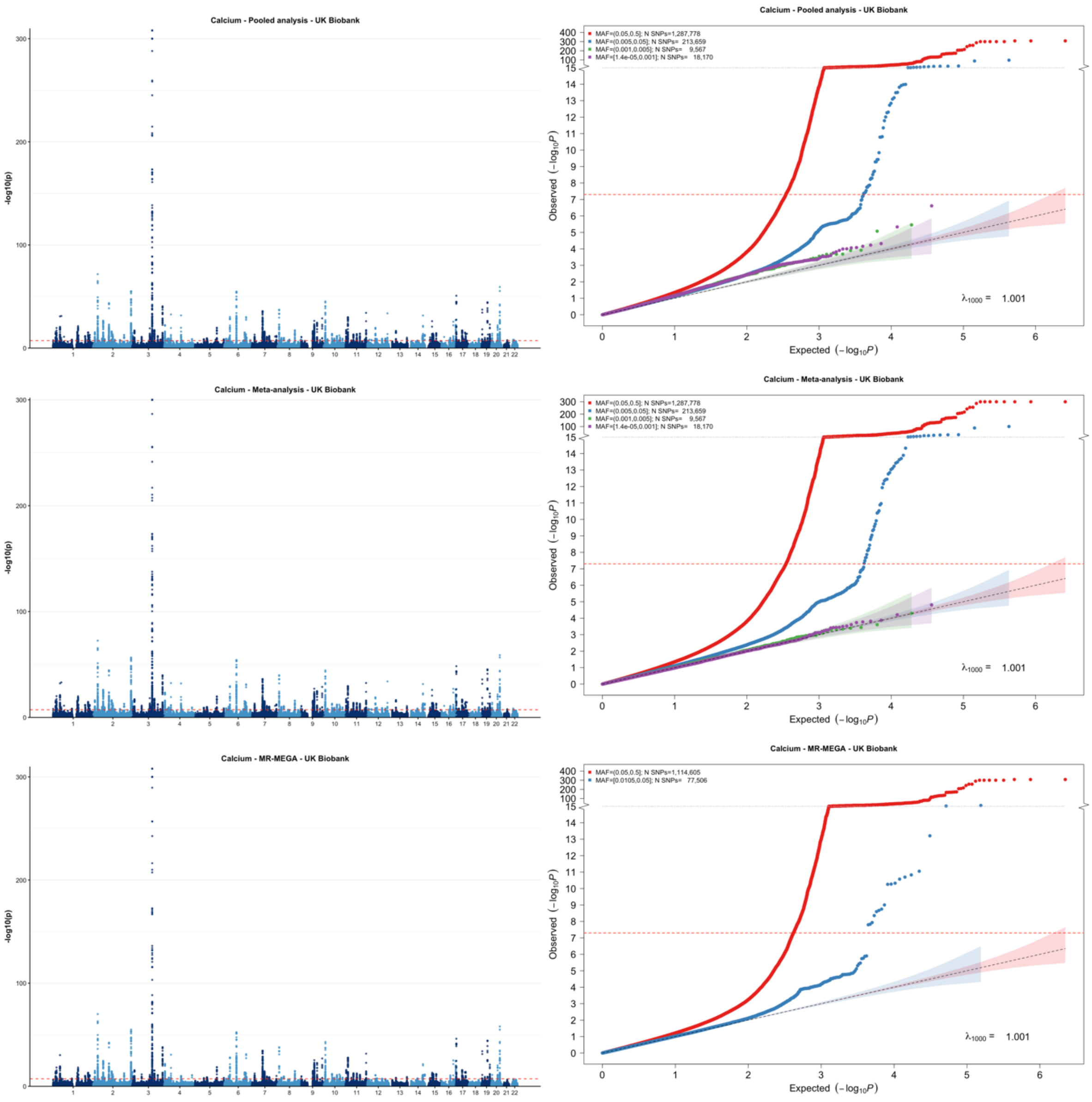


[1] For continuous traits, $\lambda_{1000}$ scales the genomic inflation factor $\lambda$ to a study with 1000 subjects using $\lambda_{1000} = 1 + 1000 * (\lambda - 1)/N$, where N is the total sample size. For binary traits, $\lambda_{1000}$ scales $\lambda$ to a study with 1000 cases and 1000 controls using $\lambda_{1000} = 1 + 1000 * (\lambda - 1) * (\frac{1}{N_{case}} + \frac{1}{N_{control}})$

**Supplementary Figure 7-g: Manhattan plot and QQ plot[1] based on the UK Biobank multi-ancestry GWAS summary statistics for creatinine in five populations (European, African American, Latino, East Asian and South Asian), obtained for pooled analysis, meta-analysis and MR-MEGA using mixed-effect modelling** (from top panel to bottom panel). The red, blue, green and purple shaded regions around the diagonal line in the QQ plots indicate the 95% confidence intervals expected under the null hypothesis of no association between genetic markers and the trait of interest, for minor allele frequencies (MAF) within the ranges (0.05, 0.5], (0.01, 0.05], (0.001, 0.005] and (0,0.001] respectively. Under the null hypothesis, the p-value follows a uniform (0,1) distribution. The jth order statistic follows a Beta (j, N-j+1) distribution, where N is the total number of variants given a specific MAF cutoff.

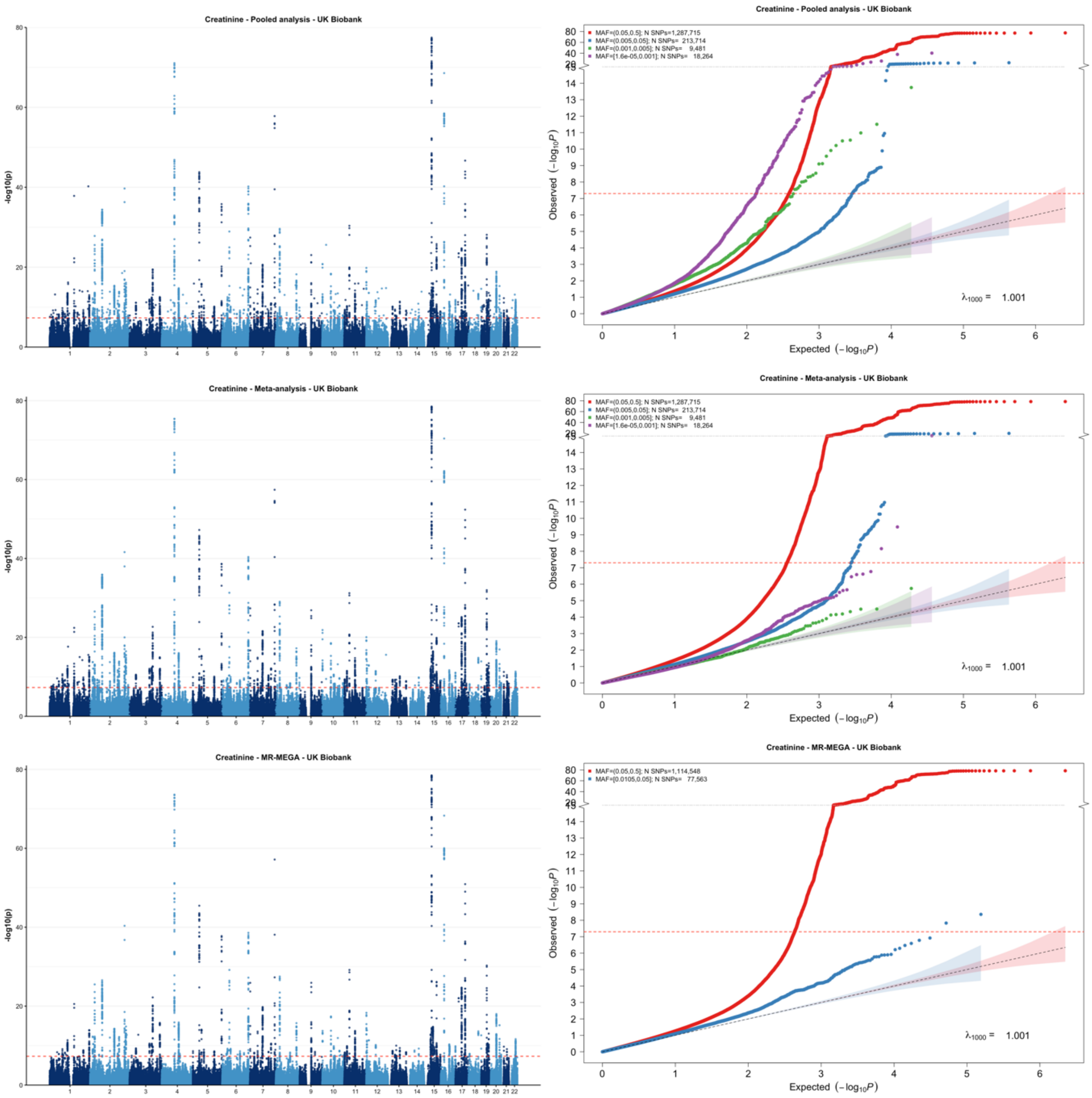


[1] For continuous traits, $\lambda_{1000}$ scales the genomic inflation factor $\lambda$ to a study with 1000 subjects using $\lambda_{1000} = 1 + 1000 * (\lambda - 1)/N$, where N is the total sample size. For binary traits, $\lambda_{1000}$ scales $\lambda$ to a study with 1000 cases and 1000 controls using $\lambda_{1000} = 1 + 1000 * (\lambda - 1) * (\frac{1}{N_{case}} + \frac{1}{N_{control}})$

**Supplementary Figure 7-h: Manhattan plot and QQ plot[1] based on the UK Biobank multi-ancestry GWAS summary statistics for EGFR in five populations (European, African American, Latino, East Asian and South Asian), obtained for pooled analysis, meta-analysis and MR-MEGA using mixed-effect modelling** (from top panel to bottom panel). The red, blue, green and purple shaded regions around the diagonal line in the QQ plots indicate the 95% confidence intervals expected under the null hypothesis of no association between genetic markers and the trait of interest, for minor allele frequencies (MAF) within the ranges (0.05, 0.5], (0.01, 0.05], (0.001, 0.005] and (0,0.001] respectively. Under the null hypothesis, the p-value follows a uniform (0,1) distribution. The jth order statistic follows a Beta (j, N-j+1) distribution, where N is the total number of variants given a specific MAF cutoff.

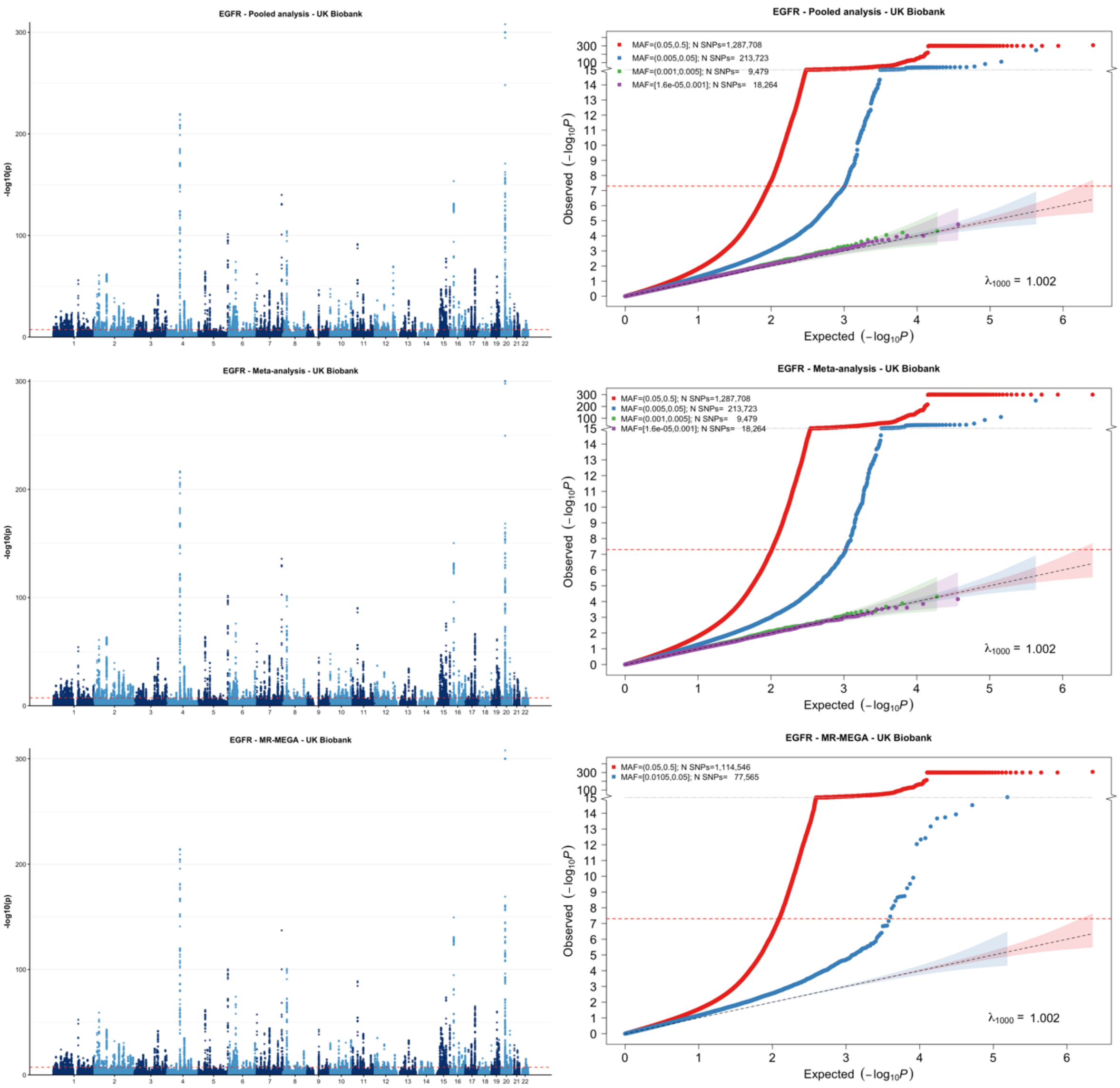


[1] For continuous traits, $\lambda_{1000}$ scales the genomic inflation factor $\lambda$ to a study with 1000 subjects using $\lambda_{1000} = 1 + 1000 * (\lambda - 1)/N$, where N is the total sample size. For binary traits, $\lambda_{1000}$ scales $\lambda$ to a study with 1000 cases and 1000 controls using $\lambda_{1000} = 1 + 1000 * (\lambda - 1) * (\frac{1}{N_{case}} + \frac{1}{N_{control}})$

**Supplementary Figure 7-i: Manhattan plot and QQ plot[1] based on the UK Biobank multi-ancestry GWAS summary statistics for asthma in five populations (European, African American, Latino, East Asian and South Asian), obtained for pooled analysis, meta-analysis and MR-MEGA using mixed-effect modelling** (from top panel to bottom panel). The red, blue, green and purple shaded regions around the diagonal line in the QQ plots indicate the 95% confidence intervals expected under the null hypothesis of no association between genetic markers and the trait of interest, for minor allele frequencies (MAF) within the ranges (0.05, 0.5], (0.01, 0.05], (0.001, 0.005] and (0,0.001] respectively. Under the null hypothesis, the p-value follows a uniform (0,1) distribution. The jth order statistic follows a Beta (j, N-j+1) distribution, where N is the total number of variants given a specific MAF cutoff.

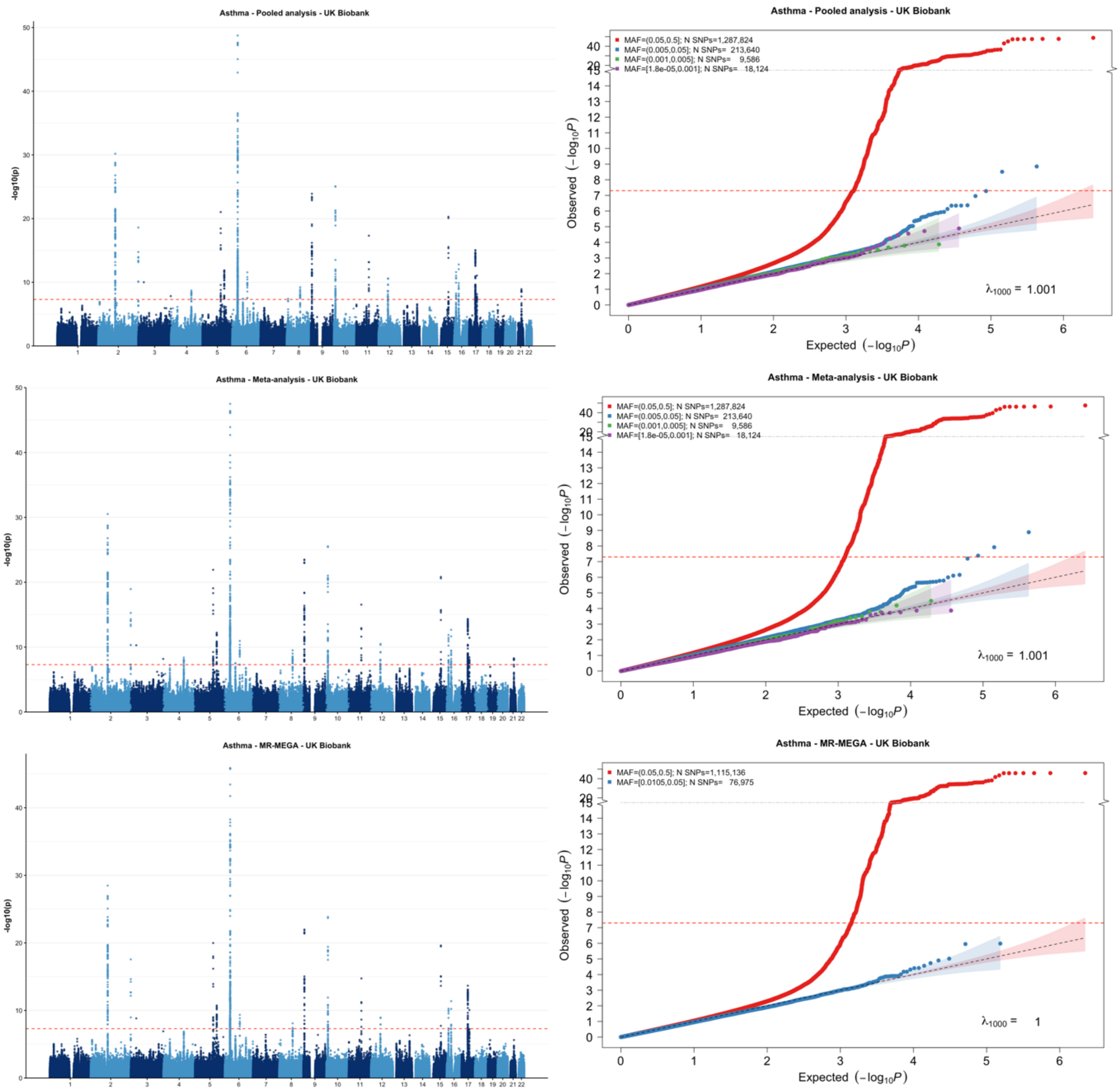


[1] For continuous traits, $\lambda_{1000}$ scales the genomic inflation factor $\lambda$ to a study with 1000 subjects using $\lambda_{1000} = 1 + 1000 * (\lambda - 1)/N$, where N is the total sample size. For binary traits, $\lambda_{1000}$ scales $\lambda$ to a study with 1000 cases and 1000 controls using $\lambda_{1000} = 1 + 1000 * (\lambda - 1) * (\frac{1}{N_{case}} + \frac{1}{N_{control}})$

**Supplementary Figure 7-j: Manhattan plot and QQ plot[1] based on the UK Biobank multi-ancestry GWAS summary statistics for CAD in five populations (European, African American, Latino, East Asian and South Asian), obtained for pooled analysis, meta-analysis and MR-MEGA using mixed-effect modelling** (from top panel to bottom panel). The red, blue, green and purple shaded regions around the diagonal line in the QQ plots indicate the 95% confidence intervals expected under the null hypothesis of no association between genetic markers and the trait of interest, for minor allele frequencies (MAF) within the ranges (0.05, 0.5], (0.01, 0.05], (0.001, 0.005] and (0,0.001] respectively. Under the null hypothesis, the p-value follows a uniform (0,1) distribution. The jth order statistic follows a Beta (j, N-j+1) distribution, where N is the total number of variants given a specific MAF cutoff.

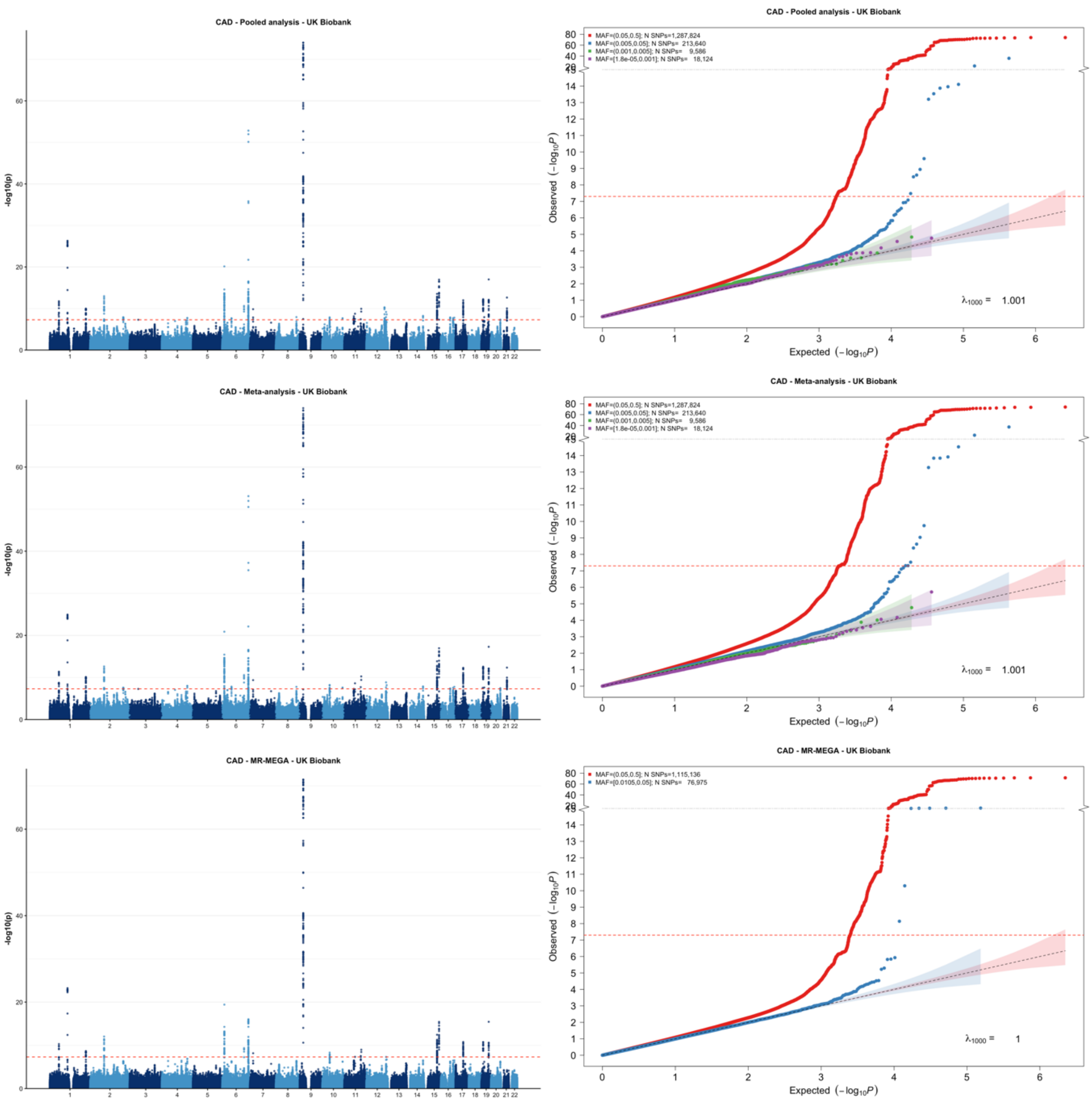


[1] For continuous traits, $\lambda_{1000}$ scales the genomic inflation factor $\lambda$ to a study with 1000 subjects using $\lambda_{1000} = 1 + 1000 * (\lambda - 1)/N$, where N is the total sample size. For binary traits, $\lambda_{1000}$ scales $\lambda$ to a study with 1000 cases and 1000 controls using $\lambda_{1000} = 1 + 1000 * (\lambda - 1) * (\frac{1}{N_{case}} + \frac{1}{N_{control}})$

**Supplementary Figure 7-k: Manhattan plot and QQ plot[1] based on the UK Biobank multi-ancestry GWAS summary statistics for type 2 diabetes in five populations (European, African American, Latino, East Asian and South Asian), obtained for pooled analysis, meta-analysis and MR-MEGA using mixed-effect modelling** (from top panel to bottom panel). The red, blue, green and purple shaded regions around the diagonal line in the QQ plots indicate the 95% confidence intervals expected under the null hypothesis of no association between genetic markers and the trait of interest, for minor allele frequencies (MAF) within the ranges (0.05, 0.5], (0.01, 0.05], (0.001, 0.005] and (0,0.001] respectively. Under the null hypothesis, the p-value follows a uniform (0,1) distribution. The jth order statistic follows a Beta (j, N-j+1) distribution, where N is the total number of variants given a specific MAF cutoff.

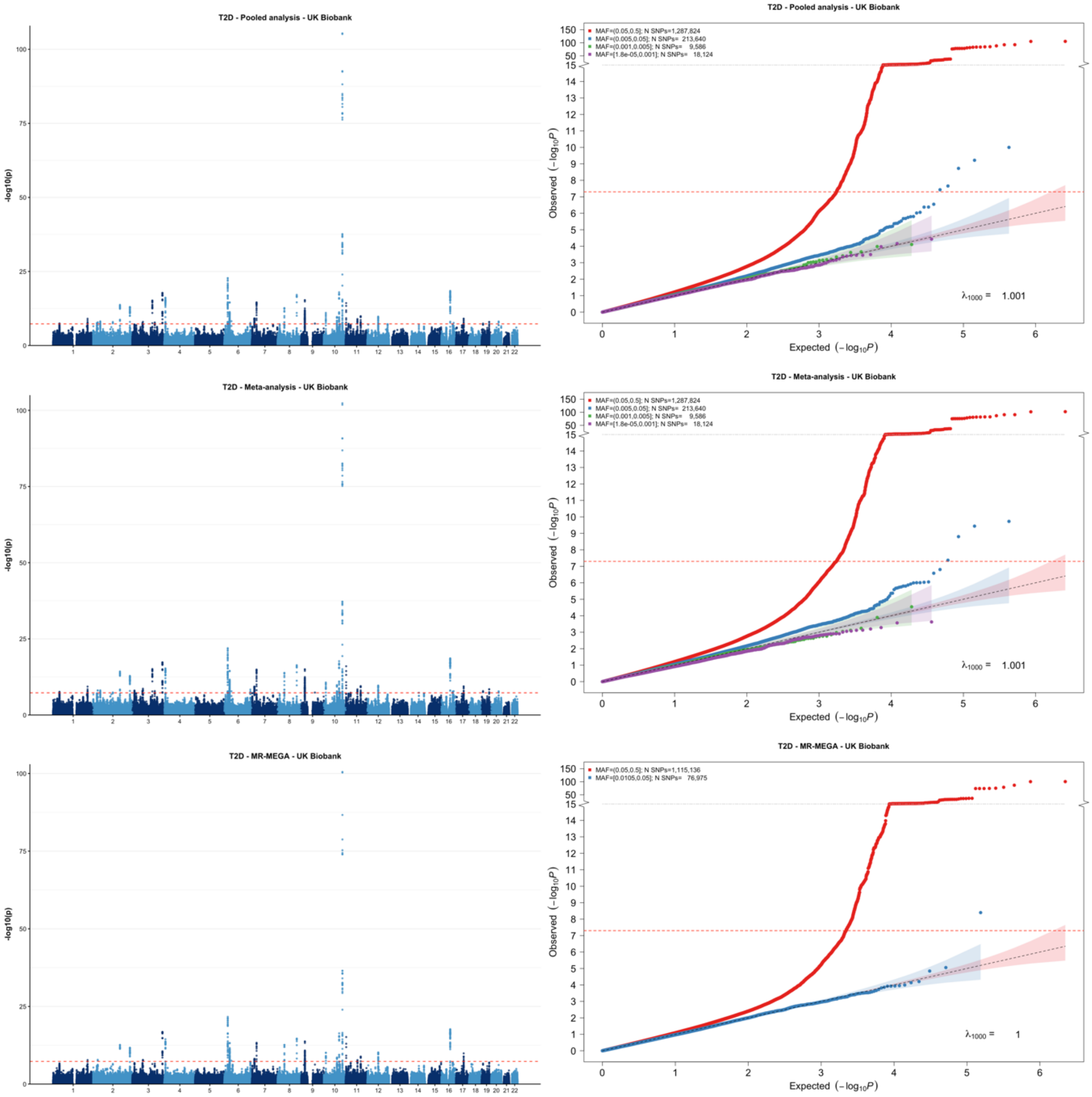


[1] For continuous traits, $\lambda_{1000}$ scales the genomic inflation factor $\lambda$ to a study with 1000 subjects using $\lambda_{1000} = 1 + 1000 * (\lambda - 1)/N$, where N is the total sample size. For binary traits, $\lambda_{1000}$ scales $\lambda$ to a study with 1000 cases and 1000 controls using $\lambda_{1000} = 1 + 1000 * (\lambda - 1) * (\frac{1}{N_{case}} + \frac{1}{N_{control}})$

**Supplementary Figure 7-I: Manhattan plot and QQ plot[1] based on the UK Biobank multi-ancestry GWAS summary statistics for breast cancer (BC) in five populations (European, African American, Latino, East Asian and South Asian), obtained for pooled analysis, meta-analysis and MR-MEGA using mixed-effect modelling** (from top panel to bottom panel). The red, blue, green and purple shaded regions around the diagonal line in the QQ plots indicate the 95% confidence intervals expected under the null hypothesis of no association between genetic markers and the trait of interest, for minor allele frequencies (MAF) within the ranges (0.05, 0.5], (0.01, 0.05], (0.001, 0.005] and (0,0.001] respectively. Under the null hypothesis, the p-value follows a uniform (0,1) distribution. The jth order statistic follows a Beta (j, N-j+1) distribution, where N is the total number of variants given a specific MAF cutoff.

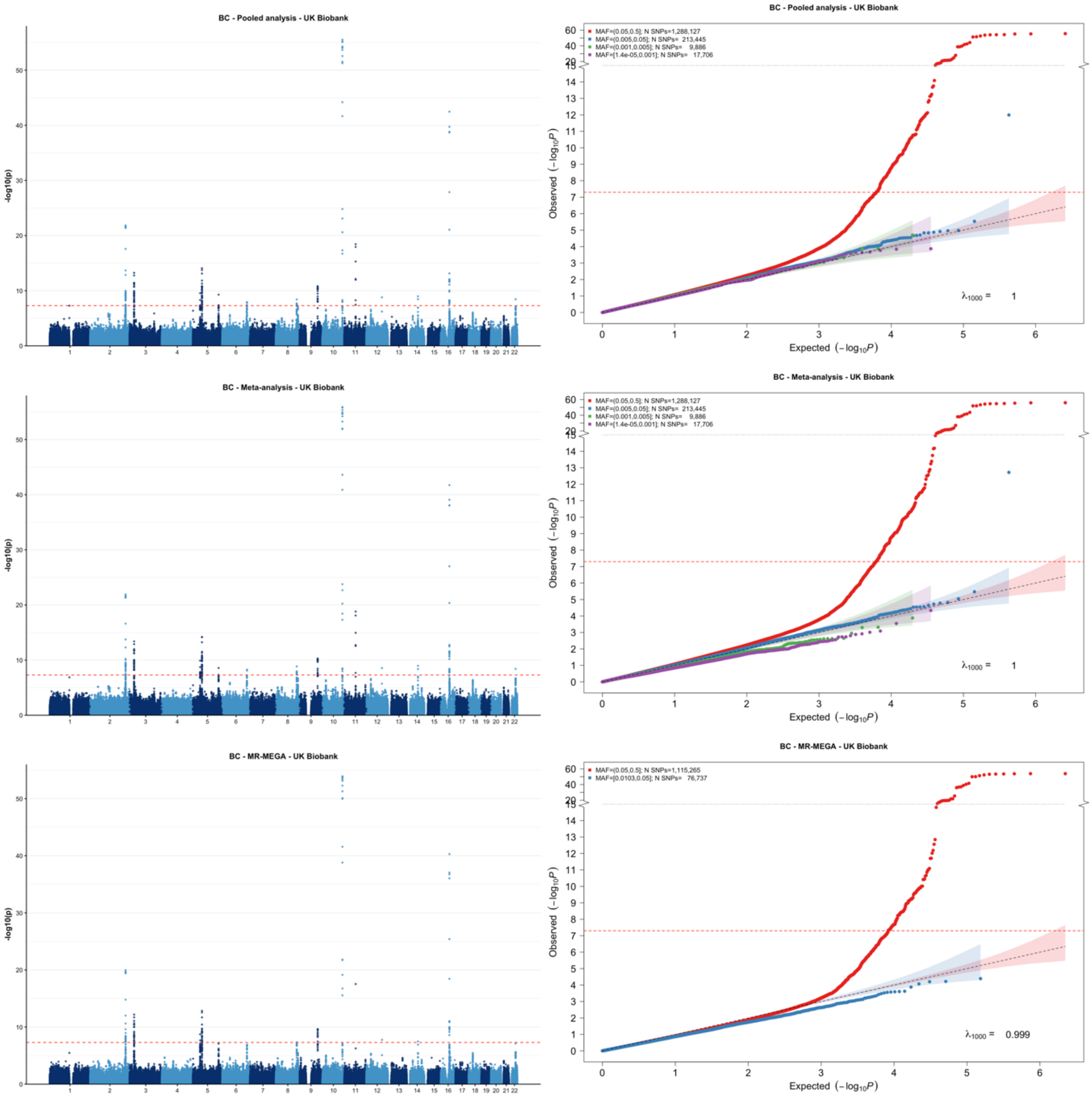


[1] For continuous traits, $\lambda_{1000}$ scales the genomic inflation factor $\lambda$ to a study with 1000 subjects using $\lambda_{1000} = 1 + 1000 * (\lambda - 1)/N$, where N is the total sample size. For binary traits, $\lambda_{1000}$ scales $\lambda$ to a study with 1000 cases and 1000 controls using $\lambda_{1000} = 1 + 1000 * (\lambda - 1) * (\frac{1}{N_{case}} + \frac{1}{N_{control}})$

**Supplementary Figure 7-m: Manhattan plot and QQ plot[1] based on the UK Biobank multi-ancestry GWAS summary statistics for prostate cancer in three populations (European, African American and Latino), obtained for pooled analysis and meta-analysis using mixed-effect modelling** (from top panel to bottom panel). The red, blue, green and purple shaded regions around the diagonal line in the QQ plots indicate the 95% confidence intervals expected under the null hypothesis of no association between genetic markers and the trait of interest, for minor allele frequencies (MAF) within the ranges (0.05, 0.5], (0.01, 0.05], (0.001, 0.005] and (0,0.001] respectively. Under the null hypothesis, the p-value follows a uniform (0,1) distribution. The jth order statistic follows a Beta (j, N-j+1) distribution, where N is the total number of variants given a specific MAF cutoff.

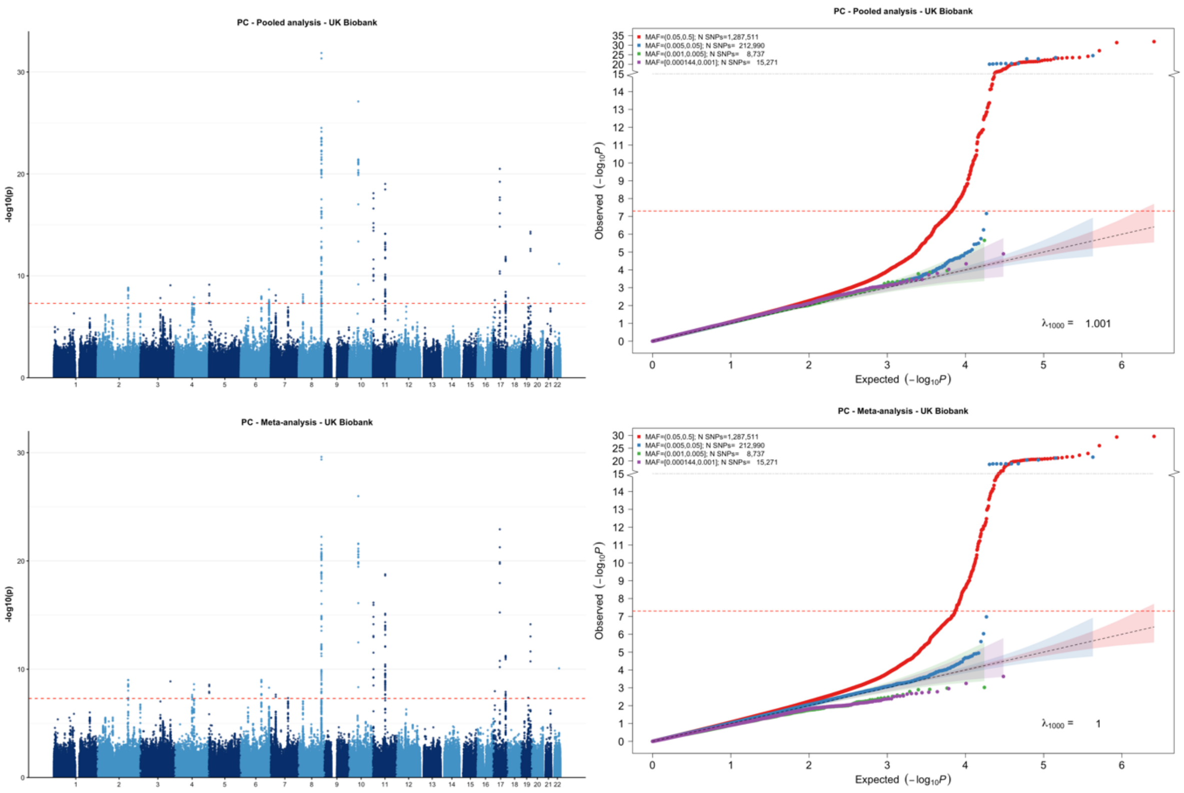


[1] For continuous traits, $\lambda_{1000}$ scales the genomic inflation factor $\lambda$ to a study with 1000 subjects using $\lambda_{1000} = 1 + 1000 * (\lambda - 1)/N$, where N is the total sample size. For binary traits, $\lambda_{1000}$ scales $\lambda$ to a study with 1000 cases and 1000 controls using $\lambda_{1000} = 1 + 1000 * (\lambda - 1) * (\frac{1}{N_{case}} + \frac{1}{N_{control}})$

**Supplementary Figure 8-a: Manhattan plot and QQ plot[1] based on the All of Us multi-ancestry GWAS summary statistics for height in six populations (European, African American, Latino, East Asian, South Asian, Middle Eastern), obtained for pooled analysis, meta-analysis and MR-MEGA using fixed-effect modelling** (from top panel to bottom panel). The red, blue, green and purple shaded regions around the diagonal line in the QQ plots indicate the 95% confidence intervals expected under the null hypothesis of no association between genetic markers and the trait of interest, for minor allele frequencies (MAF) within the ranges (0.05, 0.5], (0.01, 0.05], (0.001, 0.005] and (0,0.001] respectively. Under the null hypothesis, the p-value follows a uniform (0,1) distribution. The jth order statistic follows a Beta (j, N-j+1) distribution, where N is the total number of variants given a specific MAF cutoff.

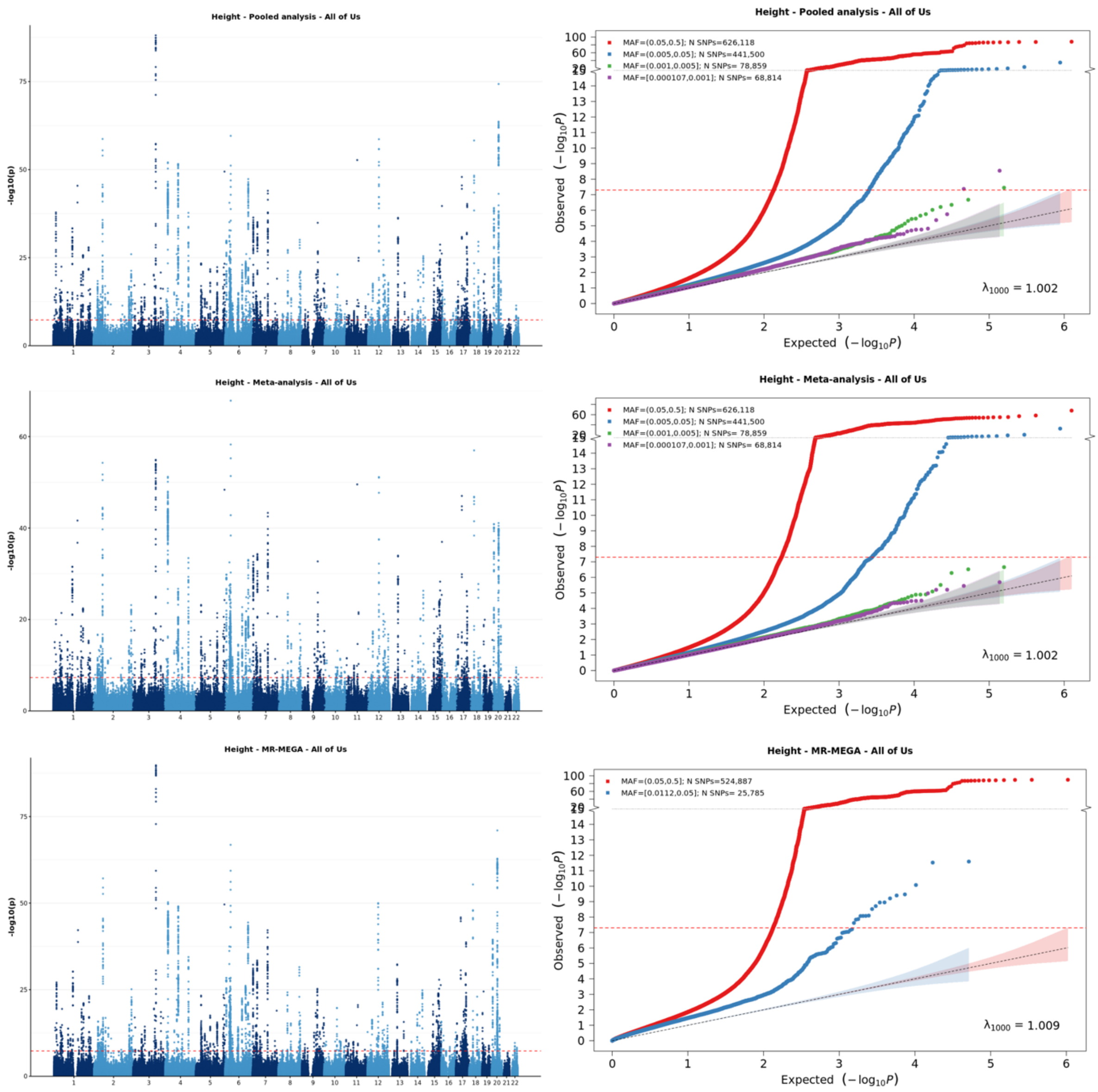


[1] For continuous traits, $\lambda_{1000}$ scales the genomic inflation factor $\lambda$ to a study with 1000 subjects using $\lambda_{1000} = 1 + 1000 * (\lambda - 1)/N$, where N is the total sample size. For binary traits, $\lambda_{1000}$ scales $\lambda$ to a study with 1000 cases and 1000 controls using $\lambda_{1000} = 1 + 1000 * (\lambda - 1) * (\frac{1}{N_{case}} + \frac{1}{N_{control}})$

**Supplementary Figure 8-b: Manhattan plot and QQ plot[1] based on the All of Us multi-ancestry GWAS summary statistics for waist circumference in six populations (European, African American, Latino, East Asian, South Asian, Middle Eastern), obtained for pooled analysis, meta-analysis and MR-MEGA using fixed-effect modelling** (from top panel to bottom panel). The red, blue, green and purple shaded regions around the diagonal line in the QQ plots indicate the 95% confidence intervals expected under the null hypothesis of no association between genetic markers and the trait of interest, for minor allele frequencies (MAF) within the ranges (0.05, 0.5], (0.01, 0.05], (0.001, 0.005] and (0,0.001] respectively. Under the null hypothesis, the p-value follows a uniform (0,1) distribution. The jth order statistic follows a Beta (j, N-j+1) distribution, where N is the total number of variants given a specific MAF cutoff.

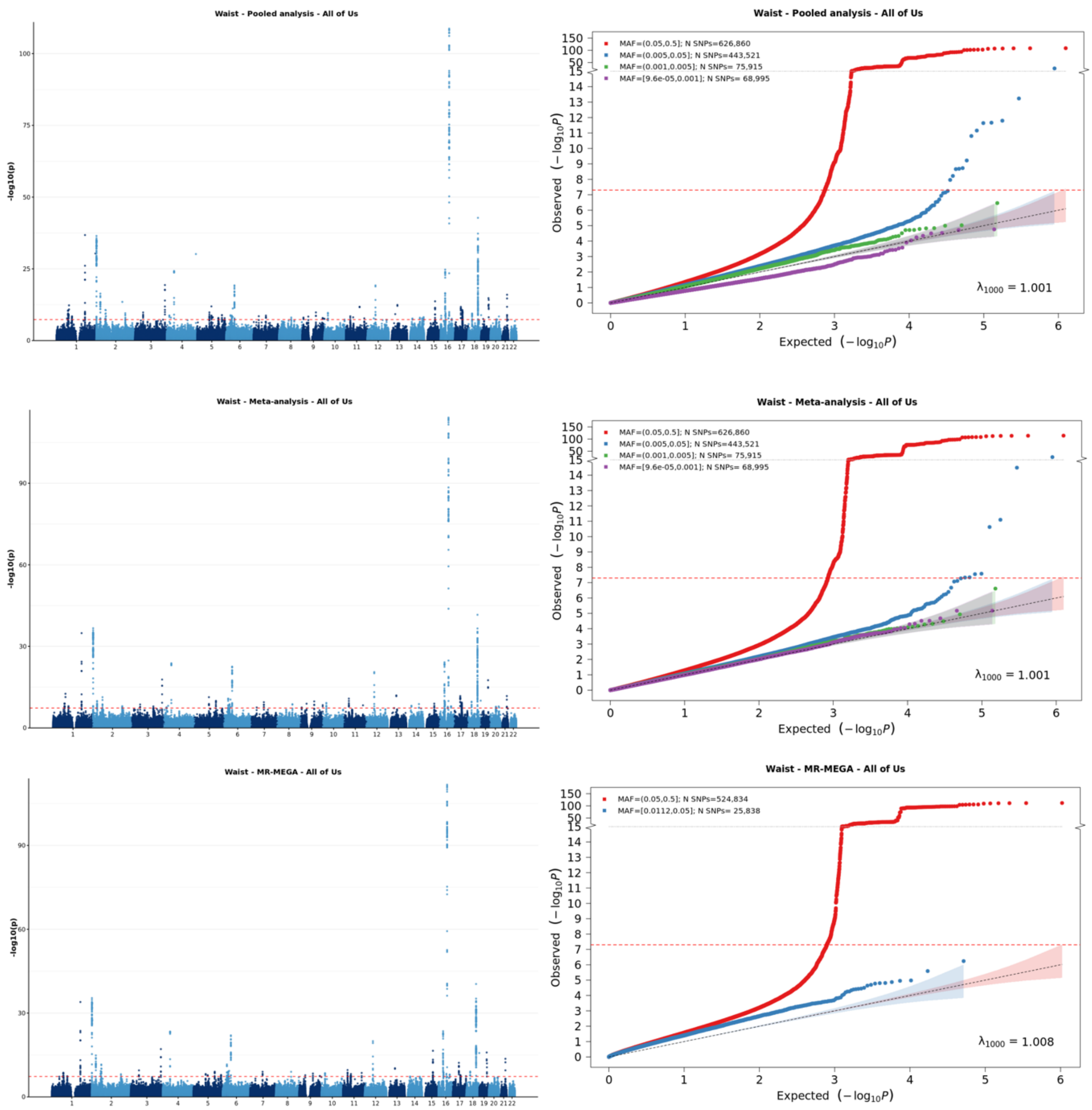


[1] For continuous traits, $\lambda_{1000}$ scales the genomic inflation factor $\lambda$ to a study with 1000 subjects using $\lambda_{1000} = 1 + 1000 * (\lambda - 1)/N$, where N is the total sample size. For binary traits, $\lambda_{1000}$ scales $\lambda$ to a study with 1000 cases and 1000 controls using $\lambda_{1000} = 1 + 1000 * (\lambda - 1) * (\frac{1}{N_{case}} + \frac{1}{N_{control}})$

**Supplementary Figure 8-c: Manhattan plot and QQ plot[1] based on the All of Us multi-ancestry GWAS summary statistics for LDL in six populations (European, African American, Latino, East Asian, South Asian, Middle Eastern), obtained for pooled analysis, meta-analysis and MR-MEGA using fixed-effect modelling (from top panel to bottom panel).** The red, blue, green and purple shaded regions around the diagonal line in the QQ plots indicate the 95% confidence intervals expected under the null hypothesis of no association between genetic markers and the trait of interest, for minor allele frequencies (MAF) within the ranges (0.05, 0.5], (0.01, 0.05], (0.001, 0.005] and (0,0.001] respectively. Under the null hypothesis, the p-value follows a uniform (0,1) distribution. The jth order statistic follows a Beta (j, N-j+1) distribution, where N is the total number of variants given a specific MAF cutoff.

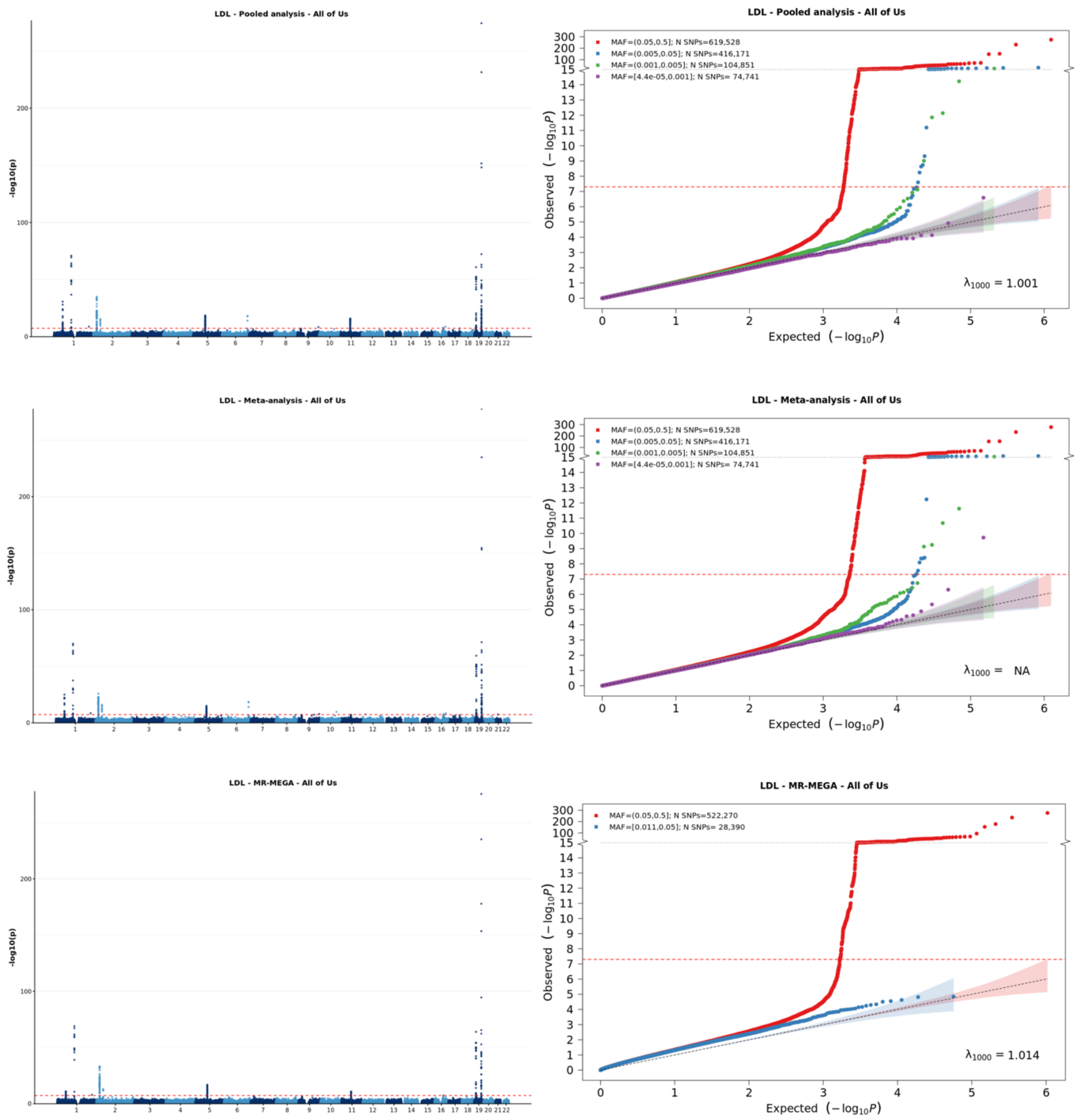


[1] For continuous traits, $\lambda_{1000}$ scales the genomic inflation factor $\lambda$ to a study with 1000 subjects using $\lambda_{1000} = 1 + 1000 * (\lambda - 1)/N$, where N is the total sample size. For binary traits, $\lambda_{1000}$ scales $\lambda$ to a study with 1000 cases and 1000 controls using $\lambda_{1000} = 1 + 1000 * (\lambda - 1) * (\frac{1}{N_{case}} + \frac{1}{N_{control}})$

**Supplementary Figure 8-d: Manhattan plot and QQ plot[1] based on the All of Us multi-ancestry GWAS summary statistics for HDL in six populations (European, African American, Latino, East Asian, South Asian, Middle Eastern), obtained for pooled analysis, meta-analysis and MR-MEGA using fixed-effect modelling** (from top panel to bottom panel). The red, blue, green and purple shaded regions around the diagonal line in the QQ plots indicate the 95% confidence intervals expected under the null hypothesis of no association between genetic markers and the trait of interest, for minor allele frequencies (MAF) within the ranges (0.05, 0.5], (0.01, 0.05], (0.001, 0.005] and (0,0.001] respectively. Under the null hypothesis, the p-value follows a uniform (0,1) distribution. The jth order statistic follows a Beta (j, N-j+1) distribution, where N is the total number of variants given a specific MAF cutoff.

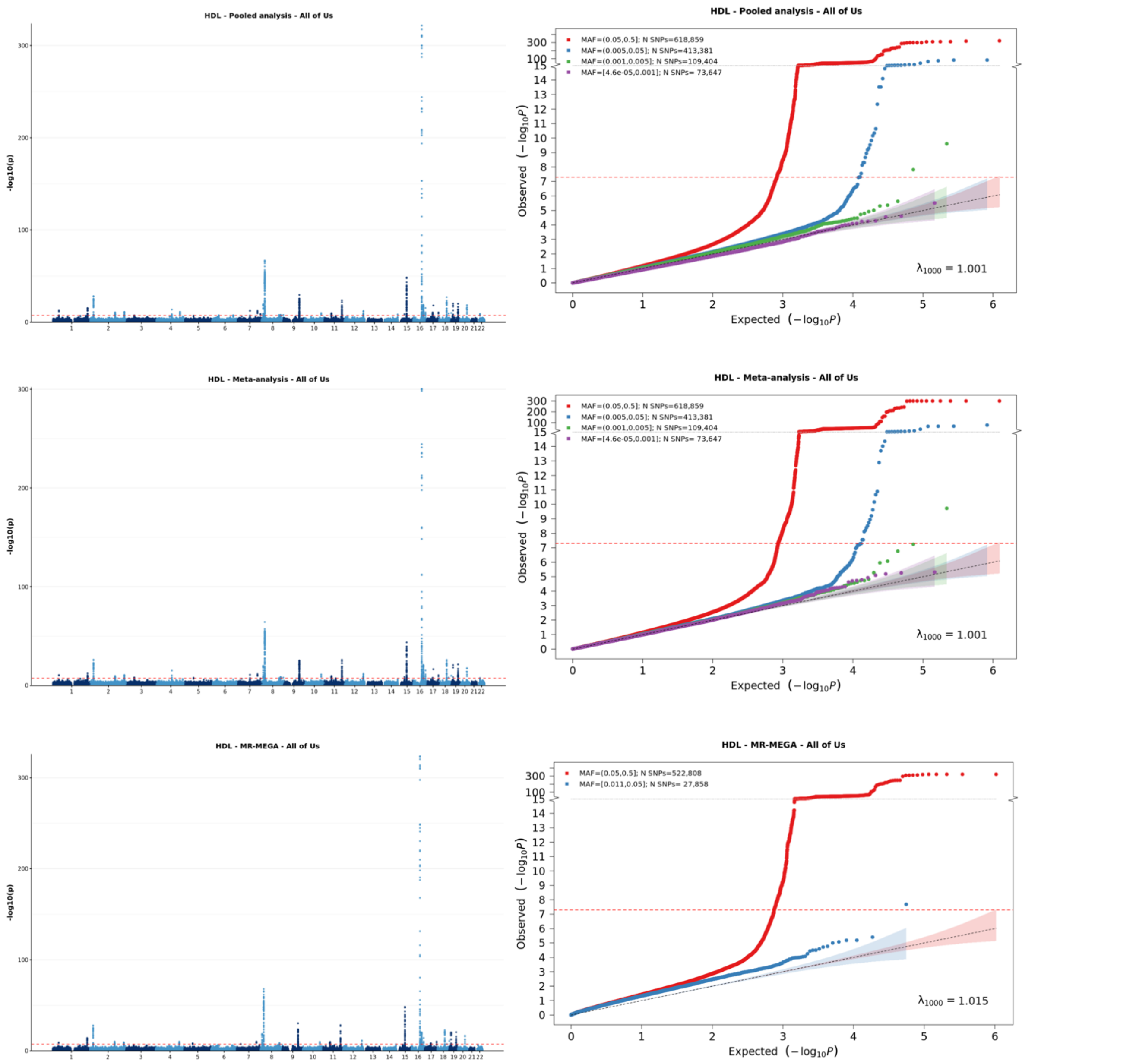


[1] For continuous traits, $\lambda_{1000}$ scales the genomic inflation factor $\lambda$ to a study with 1000 subjects using $\lambda_{1000} = 1 + 1000 * (\lambda - 1)/N$, where N is the total sample size. For binary traits, $\lambda_{1000}$ scales $\lambda$ to a study with 1000 cases and 1000 controls using $\lambda_{1000} = 1 + 1000 * (\lambda - 1) * (\frac{1}{N_{case}} + \frac{1}{N_{control}})$

**Supplementary Figure 8-e: Manhattan plot and QQ plot[1] based on the All of Us multi-ancestry GWAS summary statistics for total cholesterol (TC) in six populations (European, African American, Latino, East Asian, South Asian, Middle Eastern), obtained for pooled analysis, meta-analysis and MR-MEGA using fixed-effect modelling** (from top panel to bottom panel). The red, blue, green and purple shaded regions around the diagonal line in the QQ plots indicate the 95% confidence intervals expected under the null hypothesis of no association between genetic markers and the trait of interest, for minor allele frequencies (MAF) within the ranges (0.05, 0.5], (0.01, 0.05], (0.001, 0.005] and (0,0.001] respectively. Under the null hypothesis, the p-value follows a uniform (0,1) distribution. The jth order statistic follows a Beta (j, N-j+1) distribution, where N is the total number of variants given a specific MAF cutoff.

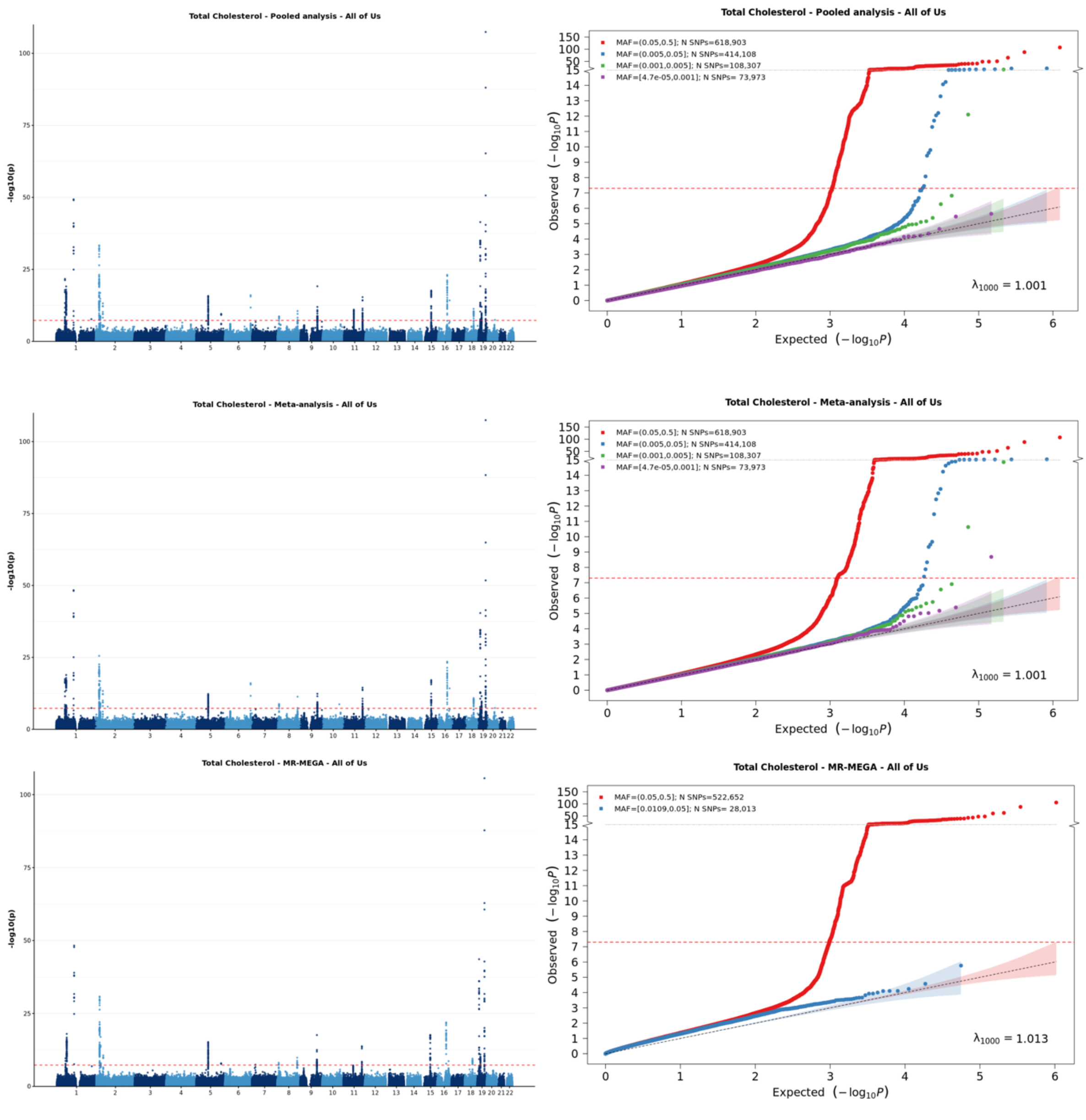


[1] For continuous traits, $\lambda_{1000}$ scales the genomic inflation factor $\lambda$ to a study with 1000 subjects using $\lambda_{1000} = 1 + 1000 * (\lambda - 1)/N$, where N is the total sample size. For binary traits, $\lambda_{1000}$ scales $\lambda$ to a study with 1000 cases and 1000 controls using $\lambda_{1000} = 1 + 1000 * (\lambda - 1) * (\frac{1}{N_{case}} + \frac{1}{N_{control}})$

**Supplementary Figure 8-f: Manhattan plot and QQ plot[1] based on the All of Us multi-ancestry GWAS summary statistics for calcium in six populations (European, African American, Latino, East Asian, South Asian, Middle Eastern), obtained for pooled analysis, meta-analysis and MR-MEGA using fixed-effect modelling** (from top panel to bottom panel). The red, blue, green and purple shaded regions around the diagonal line in the QQ plots indicate the 95% confidence intervals expected under the null hypothesis of no association between genetic markers and the trait of interest, for minor allele frequencies (MAF) within the ranges (0.05, 0.5], (0.01, 0.05], (0.001, 0.005] and (0,0.001] respectively. Under the null hypothesis, the p-value follows a uniform (0,1) distribution. The jth order statistic follows a Beta (j, N-j+1) distribution, where N is the total number of variants given a specific MAF cutoff.

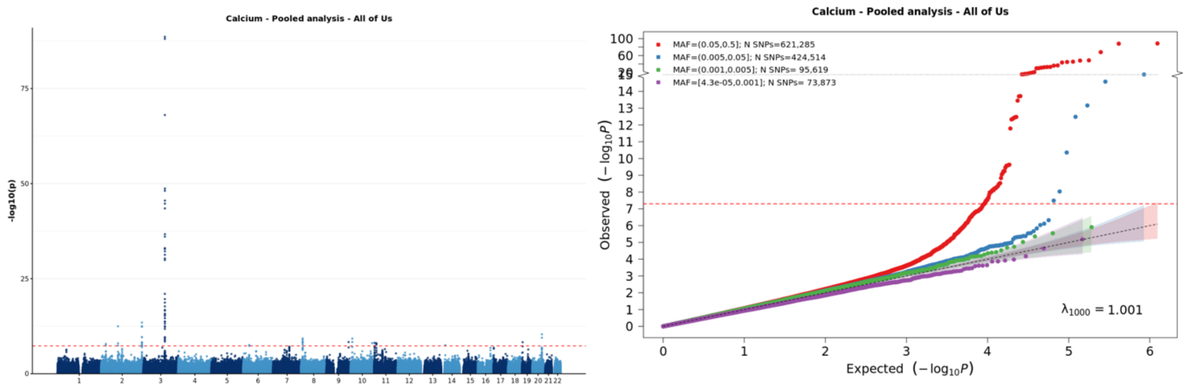


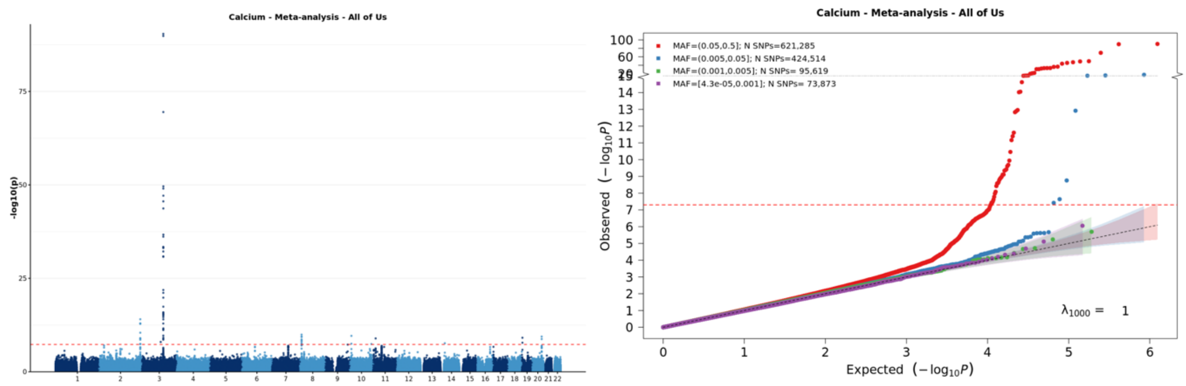


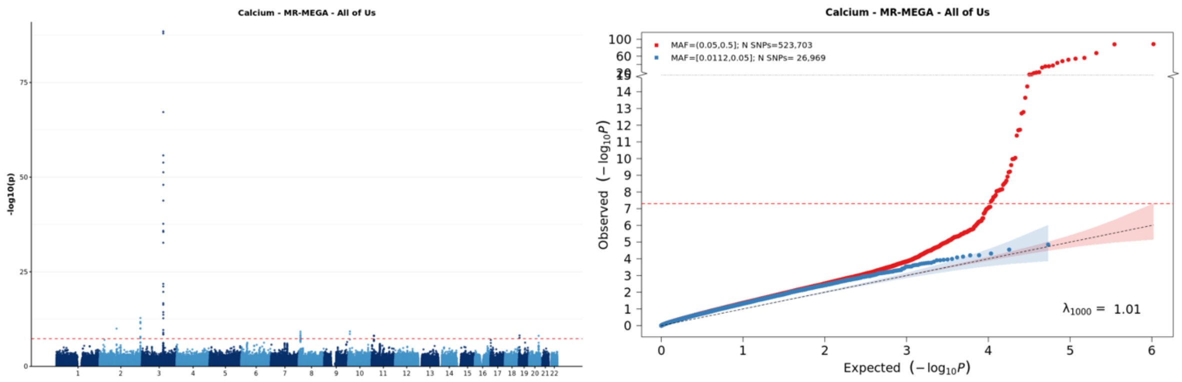


[1] For continuous traits, $\lambda_{1000}$ scales the genomic inflation factor $\lambda$ to a study with 1000 subjects using $\lambda_{1000} = 1 + 1000 * (\lambda - 1)/N$, where N is the total sample size. For binary traits, $\lambda_{1000}$ scales $\lambda$ to a study with 1000 cases and 1000 controls using $\lambda_{1000} = 1 + 1000 * (\lambda - 1) * (\frac{1}{N_{case}} + \frac{1}{N_{control}})$

**Supplementary Figure 8-g: Manhattan plot and QQ plot[1] based on the All of Us multi-ancestry GWAS summary statistics for creatinine in six populations (European, African American, Latino, East Asian, South Asian, Middle Eastern), obtained for pooled analysis, meta-analysis and MR-MEGA using fixed-effect modelling** (from top panel to bottom panel). The red, blue, green and purple shaded regions around the diagonal line in the QQ plots indicate the 95% confidence intervals expected under the null hypothesis of no association between genetic markers and the trait of interest, for minor allele frequencies (MAF) within the ranges (0.05, 0.5], (0.01, 0.05], (0.001, 0.005] and (0,0.001] respectively. Under the null hypothesis, the p-value follows a uniform (0,1) distribution. The jth order statistic follows a Beta (j, N-j+1) distribution, where N is the total number of variants given a specific MAF cutoff.

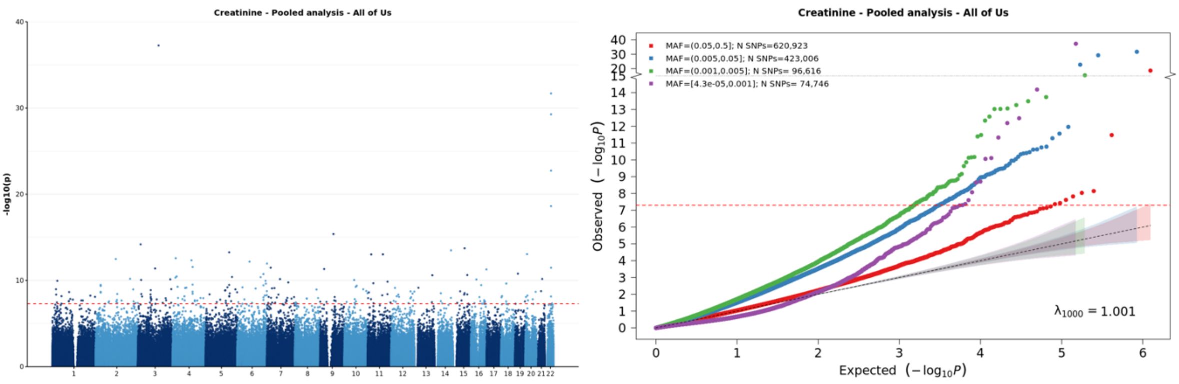


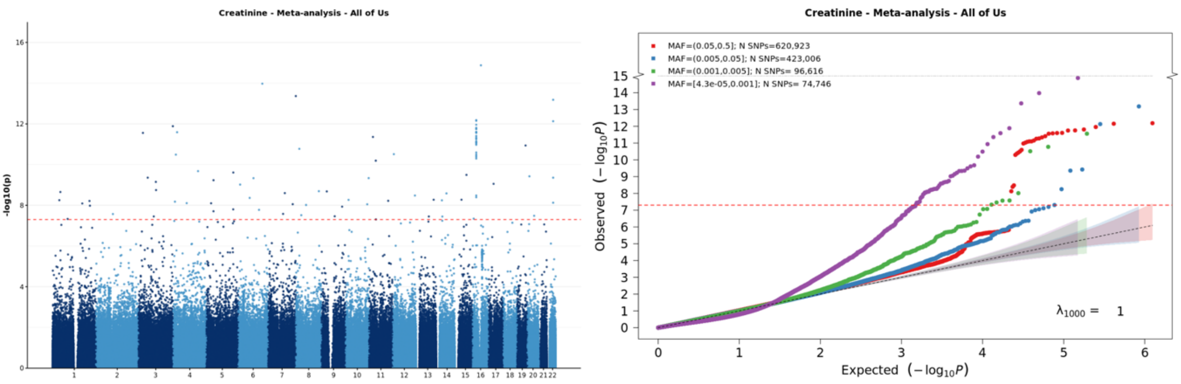


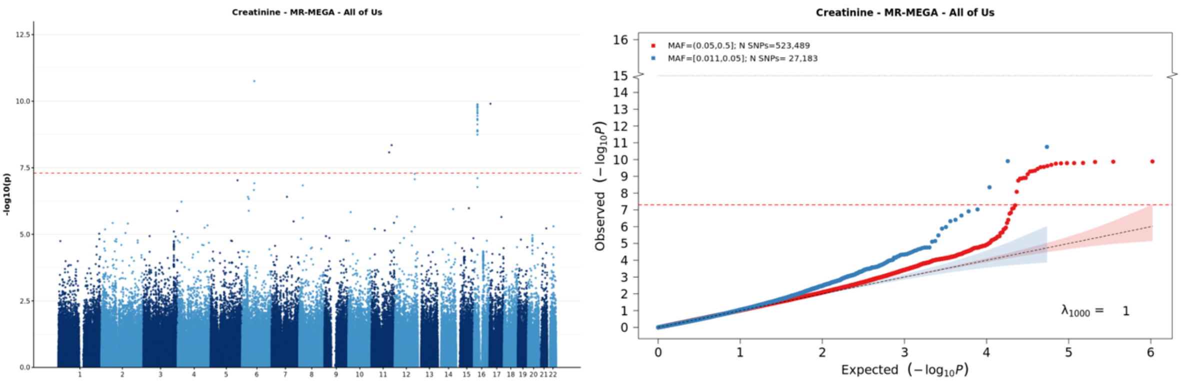


[1] For continuous traits, $\lambda_{1000}$ scales the genomic inflation factor $\lambda$ to a study with 1000 subjects using $\lambda_{1000} = 1 + 1000 * (\lambda - 1)/N$, where N is the total sample size. For binary traits, $\lambda_{1000}$ scales $\lambda$ to a study with 1000 cases and 1000 controls using $\lambda_{1000} = 1 + 1000 * (\lambda - 1) * (\frac{1}{N_{case}} + \frac{1}{N_{control}})$

**Supplementary Figure 8-h: Manhattan plot and QQ plot[1] based on the All of Us multi-ancestry GWAS summary statistics for EGFR in six populations (European, African American, Latino, East Asian, South Asian, Middle Eastern), obtained for pooled analysis, meta-analysis and MR-MEGA using fixed-effect modelling** (from top panel to bottom panel). The red, blue, green and purple shaded regions around the diagonal line in the QQ plots indicate the 95% confidence intervals expected under the null hypothesis of no association between genetic markers and the trait of interest, for minor allele frequencies (MAF) within the ranges (0.05, 0.5], (0.01, 0.05], (0.001, 0.005] and (0,0.001] respectively. Under the null hypothesis, the p-value follows a uniform (0,1) distribution. The jth order statistic follows a Beta (j, N-j+1) distribution, where N is the total number of variants given a specific MAF cutoff.

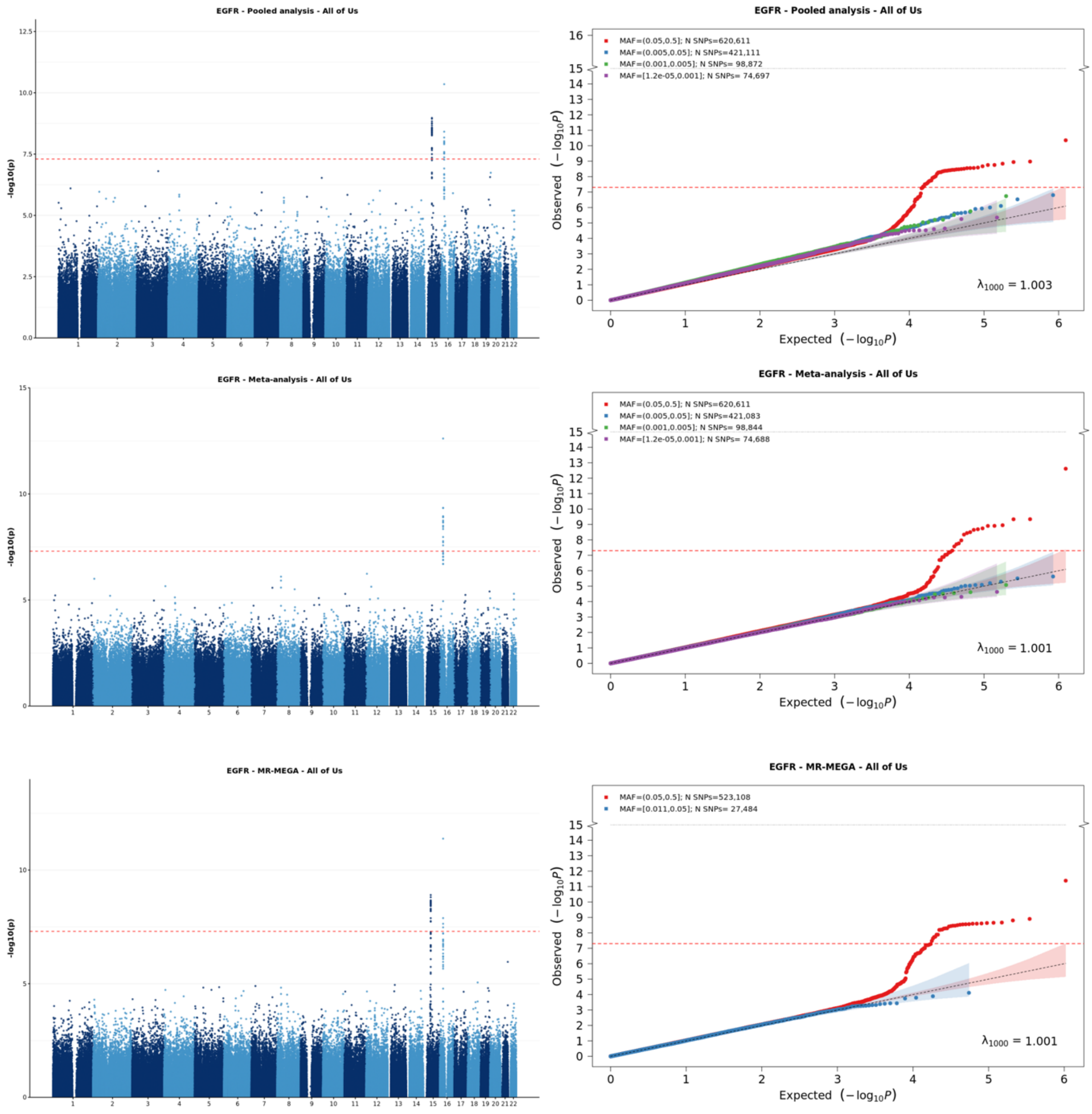


[1] For continuous traits, $\lambda_{1000}$ scales the genomic inflation factor $\lambda$ to a study with 1000 subjects using $\lambda_{1000} = 1 + 1000 * (\lambda - 1)/N$, where N is the total sample size. For binary traits, $\lambda_{1000}$ scales $\lambda$ to a study with 1000 cases and 1000 controls using $\lambda_{1000} = 1 + 1000 * (\lambda - 1) * (\frac{1}{N_{case}} + \frac{1}{N_{control}})$

**Supplementary Figure 9-a: Manhattan plot and QQ plot[1] based on the UK Biobank multi-ancestry GWAS summary statistics for height in five populations (European, African American, Latino, East Asian, South Asian) obtained for pooled analysis, meta-analysis and MR-MEGA using fixed-effect modelling**(from top panel to bottom panel). The red, blue, green and purple shaded regions around the diagonal line in the QQ plots indicate the 95% confidence intervals expected under the null hypothesis of no association between genetic markers and the trait of interest, for minor allele frequencies (MAF) within the ranges (0.05, 0.5], (0.01, 0.05], (0.001, 0.005] and (0,0.001] respectively. Under the null hypothesis, the p-value follows a uniform (0,1) distribution. The jth order statistic follows a Beta (j, N-j+1) distribution, where N is the total number of variants given a specific MAF cutoff.

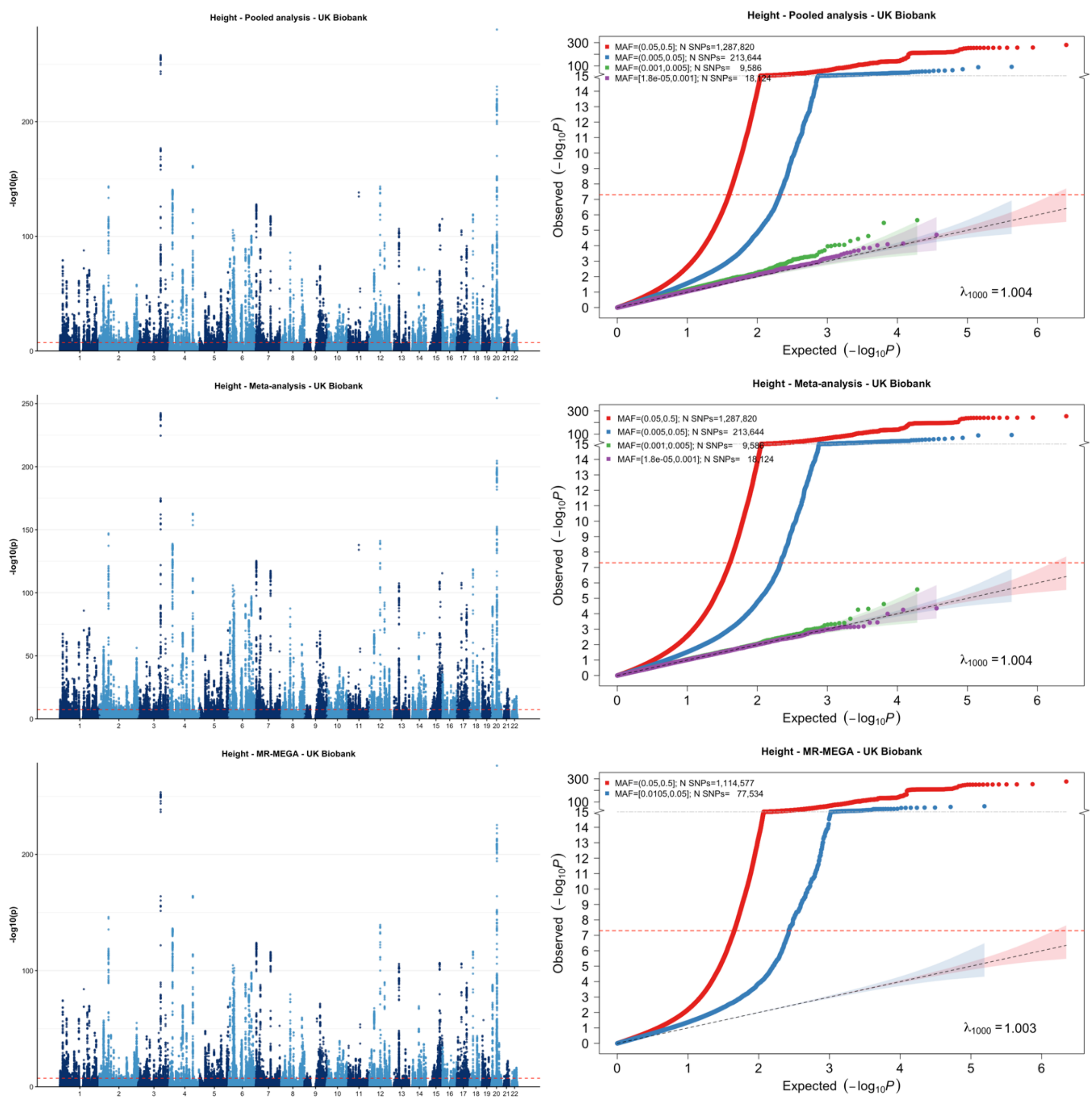


[1] For continuous traits, $\lambda_{1000}$ scales the genomic inflation factor $\lambda$ to a study with 1000 subjects using $\lambda_{1000} = 1 + 1000 * (\lambda - 1)/N$, where N is the total sample size. For binary traits, $\lambda_{1000}$ scales $\lambda$ to a study with 1000 cases and 1000 controls using $\lambda_{1000} = 1 + 1000 * (\lambda - 1) * (\frac{1}{N_{case}} + \frac{1}{N_{control}})$

**Supplementary Figure 9-b: Manhattan plot and QQ plot[1] based on the UK Biobank multi-ancestry GWAS summary statistics for waist circumference in five populations (European, African American, Latino, East Asian, South Asian) obtained for pooled analysis, meta-analysis and MR-MEGA using fixed-effect modelling** (from top panel to bottom panel). The red, blue, green and purple shaded regions around the diagonal line in the QQ plots indicate the 95% confidence intervals expected under the null hypothesis of no association between genetic markers and the trait of interest, for minor allele frequencies (MAF) within the ranges (0.05, 0.5], (0.01, 0.05], (0.001, 0.005] and (0,0.001] respectively. Under the null hypothesis, the p-value follows a uniform (0,1) distribution. The jth order statistic follows a Beta (j, N-j+1) distribution, where N is the total number of variants given a specific MAF cutoff.

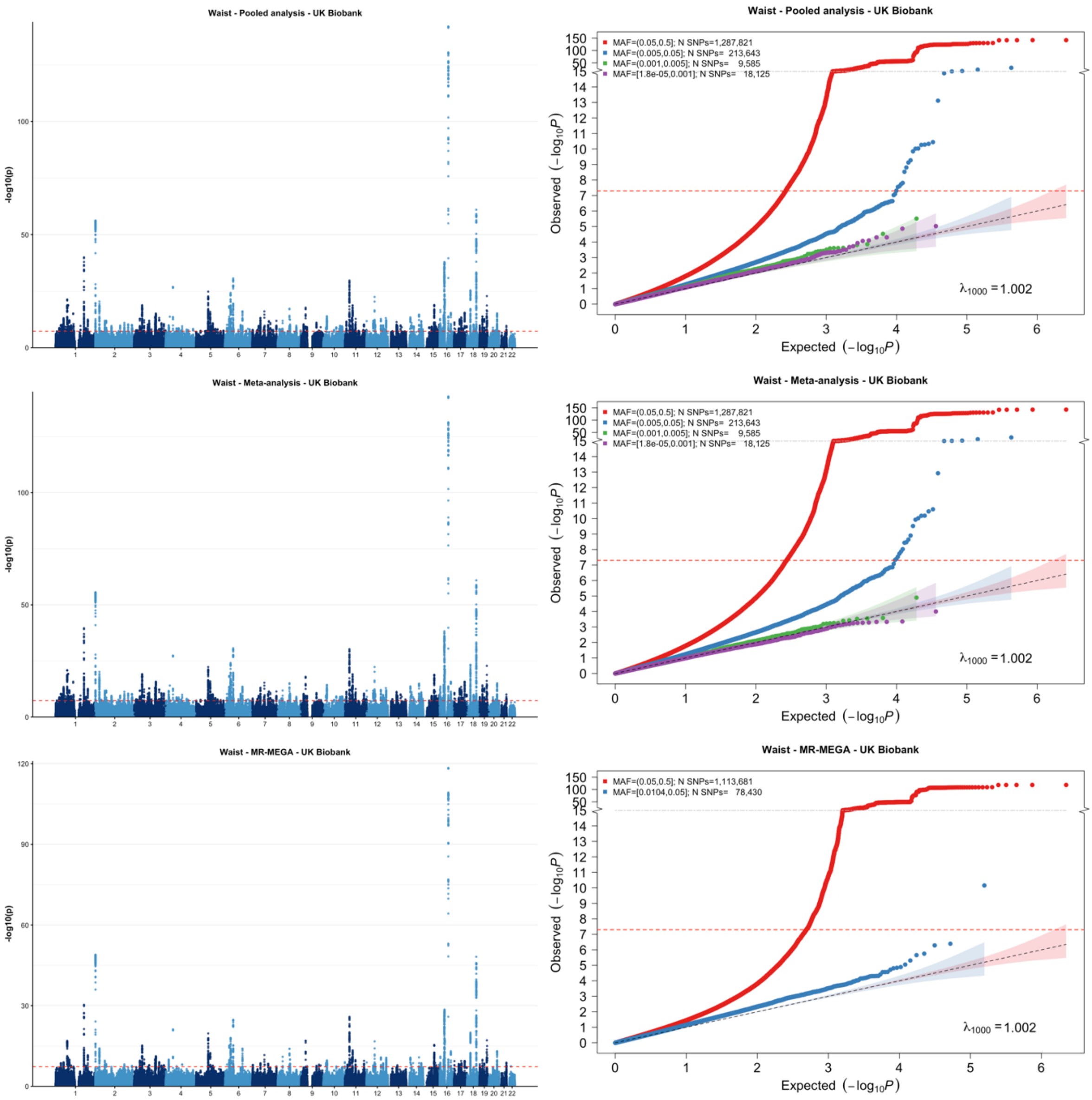


[1] For continuous traits, $\lambda_{1000}$ scales the genomic inflation factor $\lambda$ to a study with 1000 subjects using $\lambda_{1000} = 1 + 1000 * (\lambda - 1)/N$, where N is the total sample size. For binary traits, $\lambda_{1000}$ scales $\lambda$ to a study with 1000 cases and 1000 controls using $\lambda_{1000} = 1 + 1000 * (\lambda - 1) * (\frac{1}{N_{case}} + \frac{1}{N_{control}})$

**Supplementary Figure 9-c: Manhattan plot and QQ plot[1] based on the UK Biobank multi-ancestry GWAS summary statistics for LDL in five populations (European, African American, Latino, East Asian, South Asian) obtained for pooled analysis, meta-analysis and MR-MEGA using fixed-effect modelling** (from top panel to bottom panel). The red, blue, green and purple shaded regions around the diagonal line in the QQ plots indicate the 95% confidence intervals expected under the null hypothesis of no association between genetic markers and the trait of interest, for minor allele frequencies (MAF) within the ranges (0.05, 0.5], (0.01, 0.05], (0.001, 0.005] and (0,0.001] respectively. Under the null hypothesis, the p-value follows a uniform (0,1) distribution. The jth order statistic follows a Beta (j, N-j+1) distribution, where N is the total number of variants given a specific MAF cutoff.

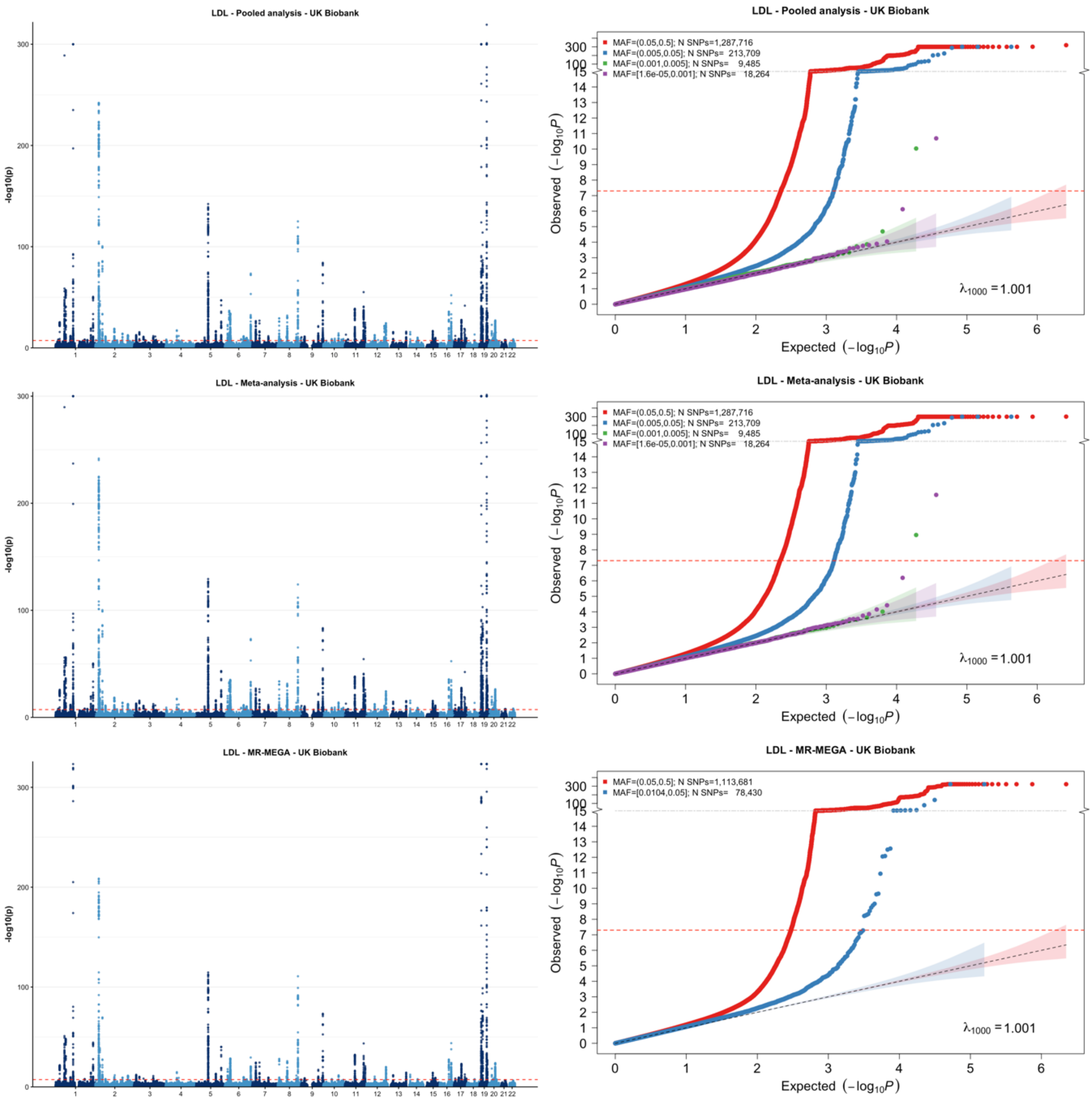


[1] For continuous traits, $\lambda_{1000}$ scales the genomic inflation factor $\lambda$ to a study with 1000 subjects using $\lambda_{1000} = 1 + 1000 * (\lambda - 1)/N$, where N is the total sample size. For binary traits, $\lambda_{1000}$ scales $\lambda$ to a study with 1000 cases and 1000 controls using $\lambda_{1000} = 1 + 1000 * (\lambda - 1) * (\frac{1}{N_{case}} + \frac{1}{N_{control}})$

**Supplementary Figure 9-d: Manhattan plot and QQ plot[1] based on the UK Biobank multi-ancestry GWAS summary statistics for HDL in five populations (European, African American, Latino, East Asian, South Asian) obtained for pooled analysis, meta-analysis and MR-MEGA using fixed-effect modelling** (from top panel to bottom panel). The red, blue, green and purple shaded regions around the diagonal line in the QQ plots indicate the 95% confidence intervals expected under the null hypothesis of no association between genetic markers and the trait of interest, for minor allele frequencies (MAF) within the ranges (0.05, 0.5], (0.01, 0.05], (0.001, 0.005] and (0,0.001] respectively. Under the null hypothesis, the p-value follows a uniform (0,1) distribution. The jth order statistic follows a Beta (j, N-j+1) distribution, where N is the total number of variants given a specific MAF cutoff.

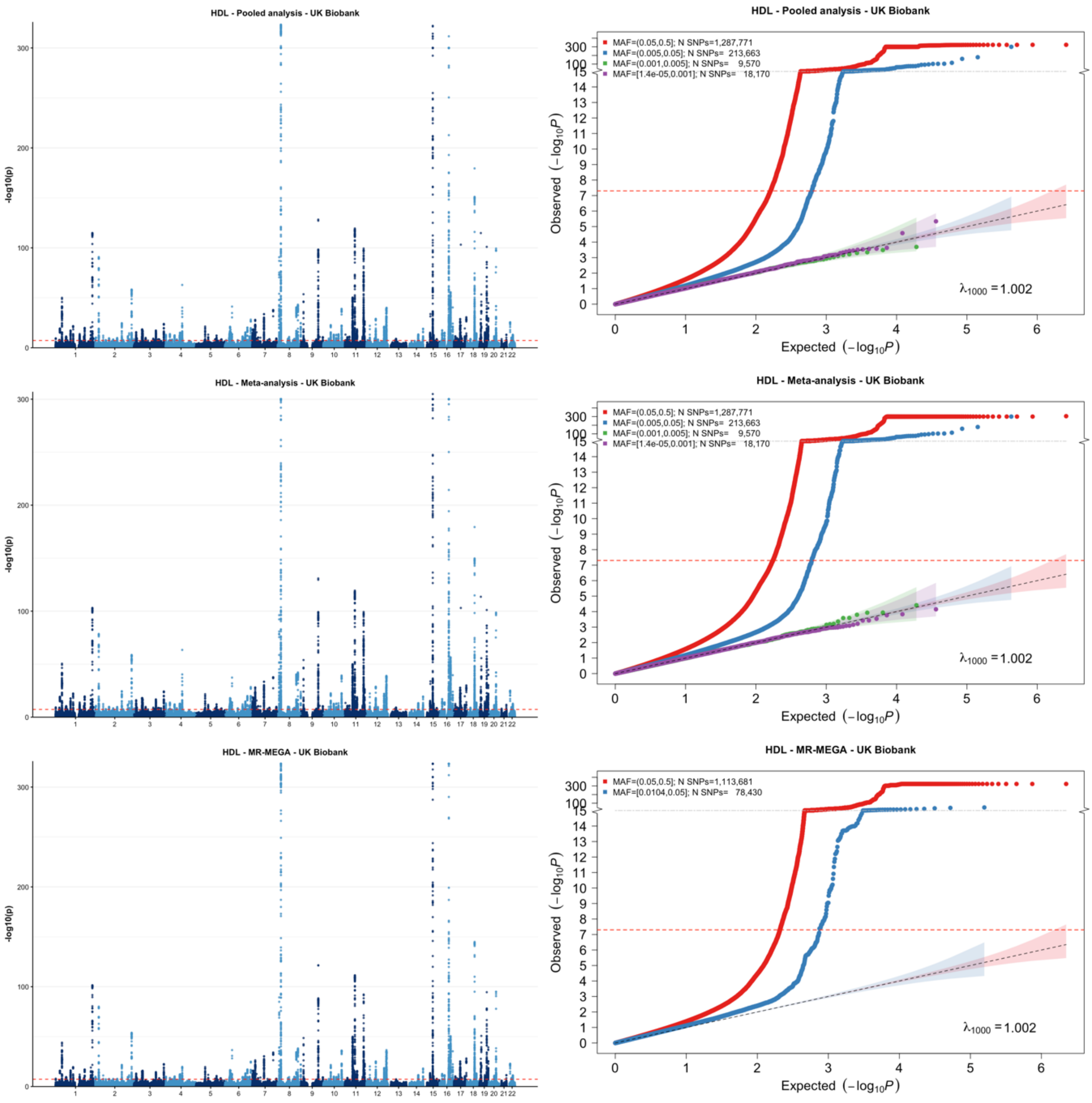


[1] For continuous traits, $\lambda_{1000}$ scales the genomic inflation factor $\lambda$ to a study with 1000 subjects using $\lambda_{1000} = 1 + 1000 * (\lambda - 1)/N$, where N is the total sample size. For binary traits, $\lambda_{1000}$ scales $\lambda$ to a study with 1000 cases and 1000 controls using $\lambda_{1000} = 1 + 1000 * (\lambda - 1) * (\frac{1}{N_{case}} + \frac{1}{N_{control}})$

**Supplementary Figure 9-e: Manhattan plot and QQ plot[1] based on the UK Biobank multi-ancestry GWAS summary statistics for total cholesterol (TC) in five populations (European, African American, Latino, East Asian, South Asian) obtained for pooled analysis, meta-analysis and MR-MEGA using fixed-effect modelling** (from top panel to bottom panel). The red, blue, green and purple shaded regions around the diagonal line in the QQ plots indicate the 95% confidence intervals expected under the null hypothesis of no association between genetic markers and the trait of interest, for minor allele frequencies (MAF) within the ranges (0.05, 0.5], (0.01, 0.05], (0.001, 0.005] and (0,0.001] respectively. Under the null hypothesis, the p-value follows a uniform (0,1) distribution. The jth order statistic follows a Beta (j, N-j+1) distribution, where N is the total number of variants given a specific MAF cutoff.

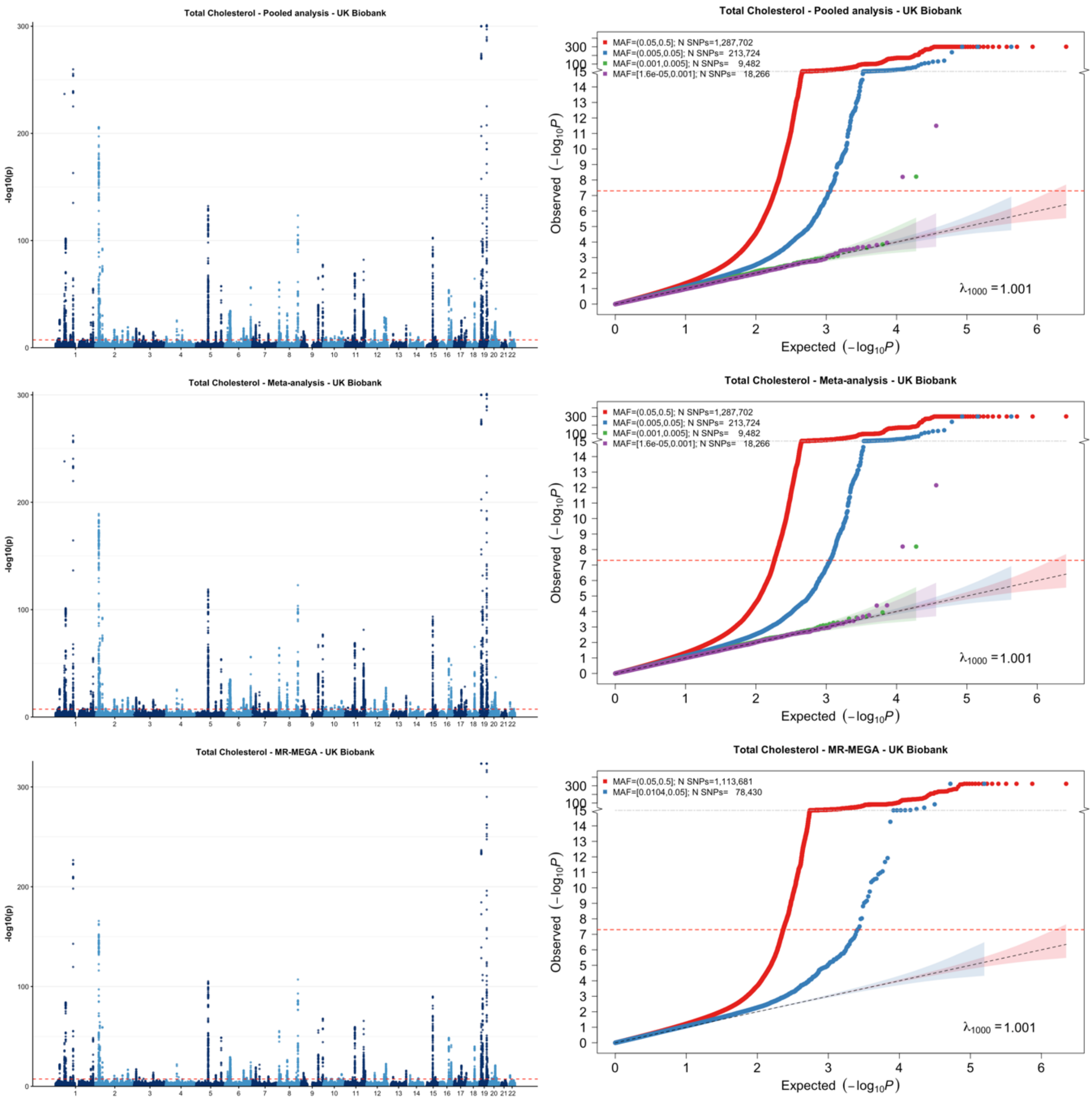


[1] For continuous traits, $\lambda_{1000}$ scales the genomic inflation factor $\lambda$ to a study with 1000 subjects using $\lambda_{1000} = 1 + 1000 * (\lambda - 1)/N$, where N is the total sample size. For binary traits, $\lambda_{1000}$ scales $\lambda$ to a study with 1000 cases and 1000 controls using $\lambda_{1000} = 1 + 1000 * (\lambda - 1) * (\frac{1}{N_{case}} + \frac{1}{N_{control}})$

**Supplementary Figure 9-f: Manhattan plot and QQ plot[1] based on the UK Biobank multi-ancestry GWAS summary statistics for calcium in five populations (European, African American, Latino, East Asian, South Asian) obtained for pooled analysis, meta-analysis and MR-MEGA using fixed-effect modelling** (from top panel to bottom panel). The red, blue, green and purple shaded regions around the diagonal line in the QQ plots indicate the 95% confidence intervals expected under the null hypothesis of no association between genetic markers and the trait of interest, for minor allele frequencies (MAF) within the ranges (0.05, 0.5], (0.01, 0.05], (0.001, 0.005] and (0,0.001] respectively. Under the null hypothesis, the p-value follows a uniform (0,1) distribution. The jth order statistic follows a Beta (j, N-j+1) distribution, where N is the total number of variants given a specific MAF cutoff.

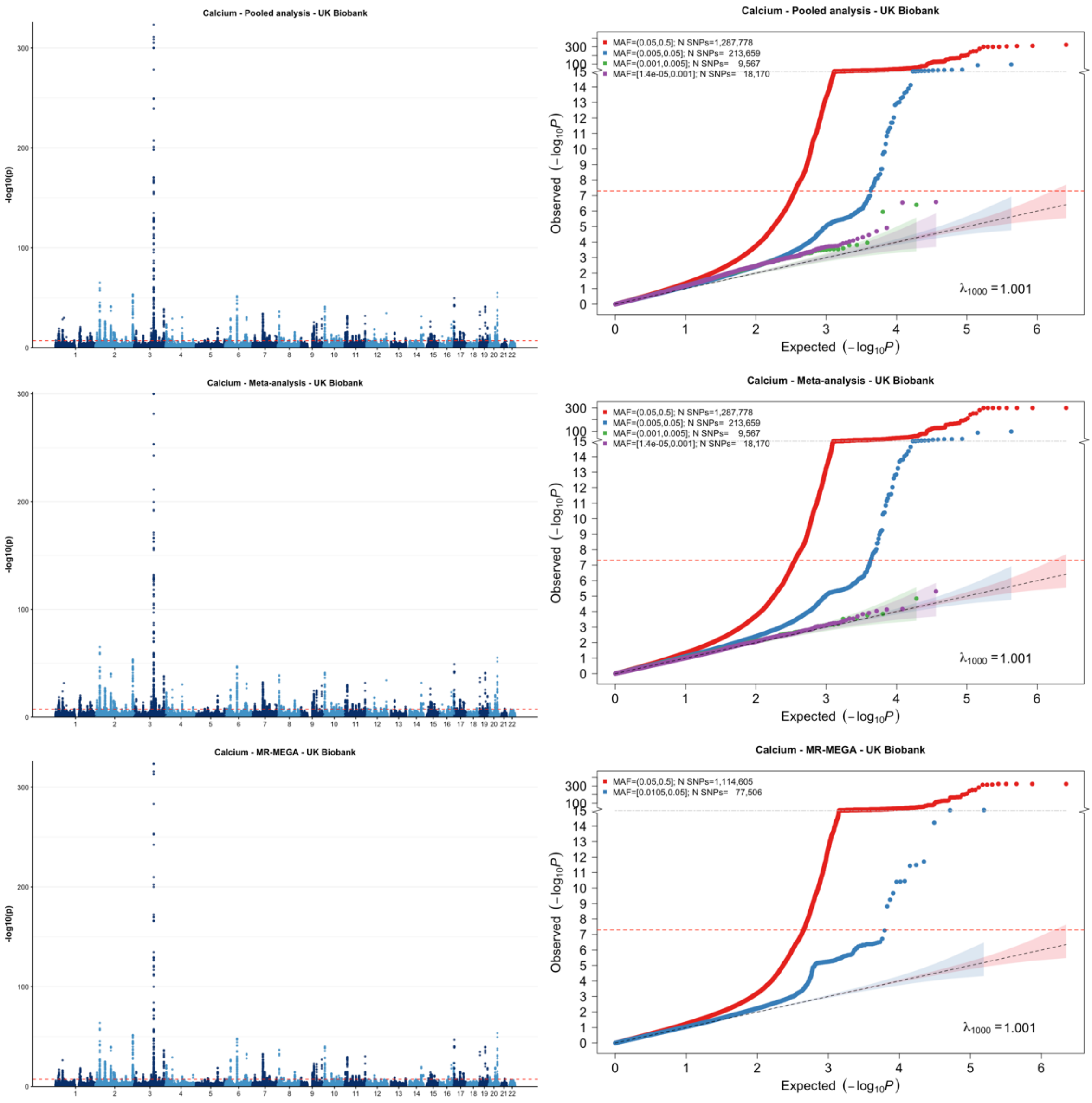


[1] For continuous traits, $\lambda_{1000}$ scales the genomic inflation factor $\lambda$ to a study with 1000 subjects using $\lambda_{1000} = 1 + 1000 * (\lambda - 1)/N$, where N is the total sample size. For binary traits, $\lambda_{1000}$ scales $\lambda$ to a study with 1000 cases and 1000 controls using $\lambda_{1000} = 1 + 1000 * (\lambda - 1) * (\frac{1}{N_{case}} + \frac{1}{N_{control}})$

**Supplementary Figure 9-g: Manhattan plot and QQ plot[1] based on the UK Biobank multi-ancestry GWAS summary statistics for creatinine in five populations (European, African American, Latino, East Asian, South Asian) obtained for pooled analysis, meta-analysis and MR-MEGA using fixed-effect modelling** (from top panel to bottom panel). The red, blue, green and purple shaded regions around the diagonal line in the QQ plots indicate the 95% confidence intervals expected under the null hypothesis of no association between genetic markers and the trait of interest, for minor allele frequencies (MAF) within the ranges (0.05, 0.5], (0.01, 0.05], (0.001, 0.005] and (0,0.001] respectively. Under the null hypothesis, the p-value follows a uniform (0,1) distribution. The jth order statistic follows a Beta (j, N-j+1) distribution, where N is the total number of variants given a specific MAF cutoff.

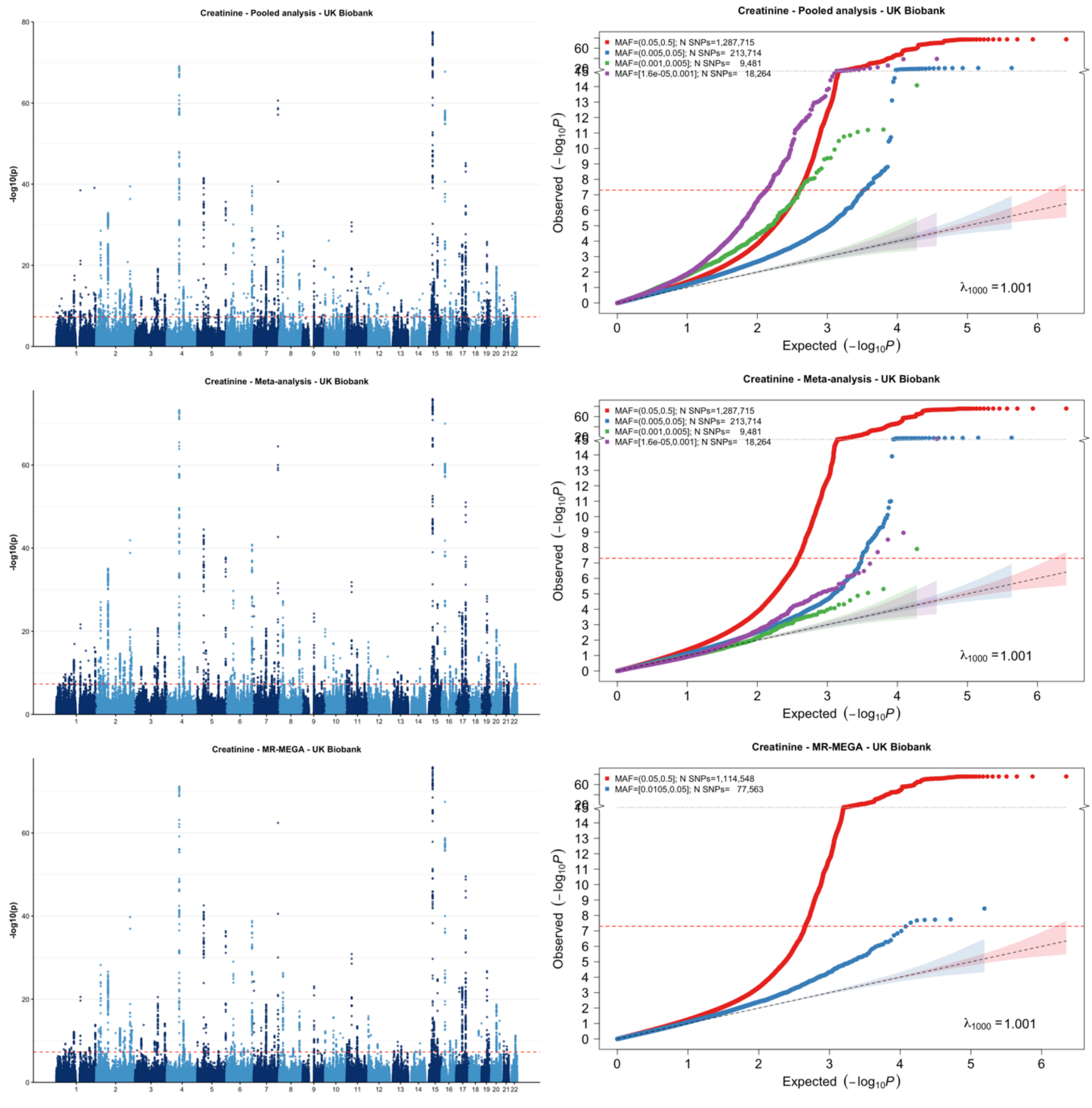


[1] For continuous traits, $\lambda_{1000}$ scales the genomic inflation factor $\lambda$ to a study with 1000 subjects using $\lambda_{1000} = 1 + 1000 * (\lambda - 1)/N$, where N is the total sample size. For binary traits, $\lambda_{1000}$ scales $\lambda$ to a study with 1000 cases and 1000 controls using $\lambda_{1000} = 1 + 1000 * (\lambda - 1) * (\frac{1}{N_{case}} + \frac{1}{N_{control}})$

**Supplementary Figure 9-h: Manhattan plot and QQ plot[1] based on the UK Biobank multi-ancestry GWAS summary statistics for EGFR in five populations (European, African American, Latino, East Asian, South Asian) obtained for pooled analysis, meta-analysis and MR-MEGA using fixed-effect modelling** (from top panel to bottom panel). The red, blue, green and purple shaded regions around the diagonal line in the QQ plots indicate the 95% confidence intervals expected under the null hypothesis of no association between genetic markers and the trait of interest, for minor allele frequencies (MAF) within the ranges (0.05, 0.5], (0.01, 0.05], (0.001, 0.005] and (0,0.001] respectively. Under the null hypothesis, the p-value follows a uniform (0,1) distribution. The jth order statistic follows a Beta (j, N-j+1) distribution, where N is the total number of variants given a specific MAF cutoff.

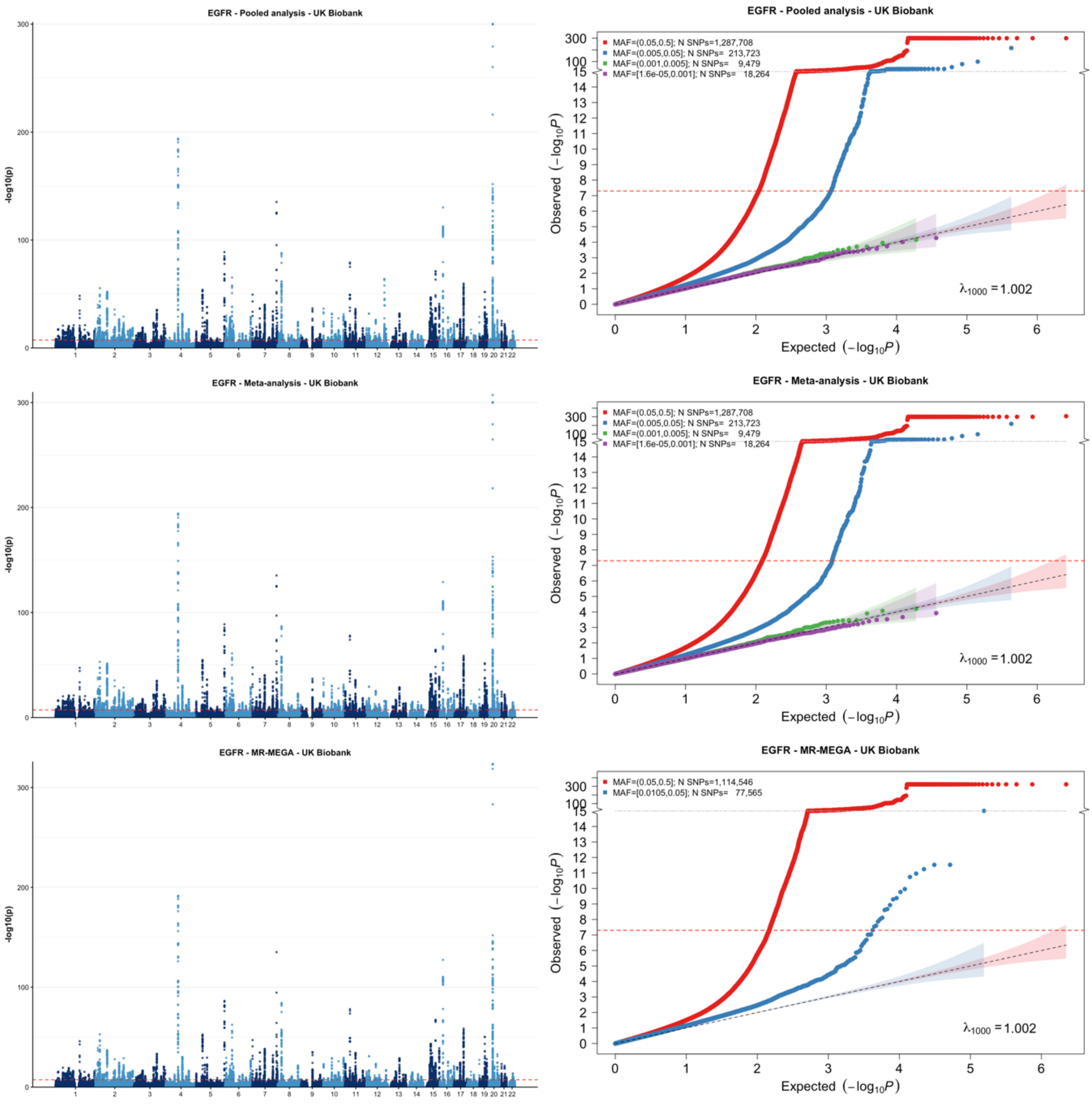


[1] For continuous traits, $\lambda_{1000}$ scales the genomic inflation factor $\lambda$ to a study with 1000 subjects using $\lambda_{1000} = 1 + 1000 * (\lambda - 1)/N$, where N is the total sample size. For binary traits, $\lambda_{1000}$ scales $\lambda$ to a study with 1000 cases and 1000 controls using $\lambda_{1000} = 1 + 1000 * (\lambda - 1) * (\frac{1}{N_{case}} + \frac{1}{N_{control}})$
